# Parental body mass index and offspring childhood body size and eating behaviour: causal inference via parental comparisons and extended children of twins structural equation modelling

**DOI:** 10.1101/2023.02.06.23284912

**Authors:** Tom A Bond, Tom A McAdams, Nicole M Warrington, Laurie J Hannigan, Espen Moen Eilertsen, Ziada Ayorech, Fartein A Torvik, George Davey Smith, Deborah A Lawlor, Eivind Ystrøm, Alexandra Havdahl, David M Evans

## Abstract

**Background:** The intergenerational transmission of obesity-related traits could propagate an accelerating cycle of obesity, if parental adiposity causally influences offspring adiposity via intrauterine or periconceptional mechanisms. We aimed to establish whether associations between parental peri-pregnancy body mass index (BMI) and offspring birth weight (BW), BMI until 8 years and 8-year eating behaviour are due to genetic confounding.

**Methods:** We used data from the Norwegian Mother, Father and Child Cohort Study and the Medical Birth Registry of Norway. We compared the strength of the associations of maternal versus paternal BMI with offspring outcomes, and used an extended children of twins structural equation model (SEM) to quantify the extent to which associations were due to genetic confounding (n = 17001 to 85866 children).

**Findings:** Maternal BMI was more strongly associated than paternal BMI with offspring BW, but the maternal-paternal difference decreased for offspring BMI after birth. Greater parental BMI was associated with obesity-related offspring eating behaviours. SEM results indicated that genetic confounding did not explain the association between parental BMI and offspring BW, but explained the majority of the association with offspring BMI from 6 months onwards. For 8-year BMI, genetic confounding explained 79% (95% CI: 62%, 95%) of the covariance with maternal BMI and 94% (95% CI: 72%, 113%) of the covariance with paternal BMI.

**Interpretation:** We found strong evidence that parent-child BMI associations are primarily due to genetic confounding, arguing against a strong causal effect of maternal or paternal adiposity on childhood adiposity via intrauterine or periconceptional mechanisms.

## Introduction

The positive observational association between parental body mass index (BMI) and offspring adiposity in childhood is well replicated (1), but the mechanisms driving this association remain unknown. If greater maternal or paternal BMI causes greater offspring BMI via prenatal or intrauterine developmental mechanisms, a vicious cycle could amplify BMI through successive generations and be a major driver of the obesity epidemic (2). It is therefore crucial to establish why parental BMI is associated with offspring childhood BMI.

Several mechanisms are plausible (**Figure 1a**). Higher parental BMI could cause higher offspring adiposity through pre-conception and/or intrauterine developmental mechanisms (the developmental overnutrition hypothesis) (3-6), with some authors advocating that interventions to maintain women ‘s preconception BMI at a healthy level be used as a means to reduce offspring adiposity (1, 3, 7). Because adiposity is highly heritable across the life course (8), genetic confounding (via the inheritance of parental genetic alleles by the offspring) could result in intergenerational BMI associations. Non-genetic (environmental) confounding, for example via shared familial socioeconomic position or parental influences on offspring postnatal food intake and physical activity behaviours, could also contribute to these associations.

**Figure 1:**
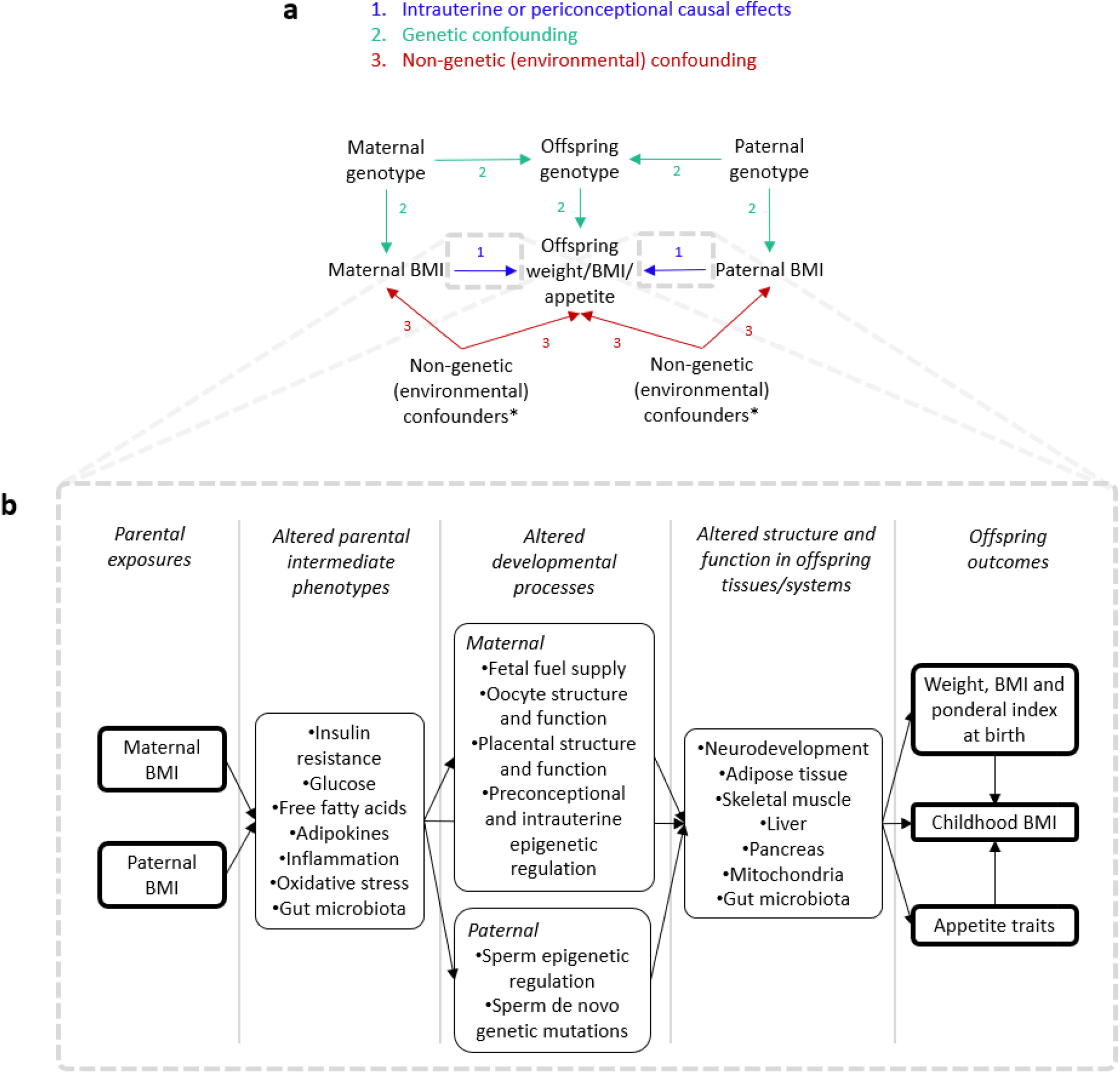
**(a)** Directed acyclic graph (DAG) showing three plausible mechanisms for associations between parental BMI and offspring weight, BMI and appetite traits. *We define non-genetic (environmental) confounding as confounding that does not involve the offspring ‘s own genotype (non-genetic confounding could therefore still involve parental genetic effects, i.e. effects of parental genotype on offspring outcomes independently of offspring genotype, via the offspring ‘s environment). Examples of non-genetic confounding include shared familial socioeconomic position or parental influences on offspring postnatal food intake and physical activity behaviours. To the extent that postnatal parental BMI *per se* causally influences offspring outcomes after birth (e.g. by influencing feeding- and exercise-related parenting practices), this would constitute a postnatal causal effect rather than non-genetic confounding. **(b)** Conceptual diagram showing putative biological mechanisms by which parental BMI could have intrauterine or periconceptional causal effects on offspring outcomes. Arrows denote potential causal effects, bold outlined boxes denote variables analysed in the present study. The intent is to non-exhaustively show some key variables and relationships that have been hypothesized in the literature

Numerous animal studies purport to provide evidence in favour of the developmental overnutrition hypothesis (9). Potential biological mechanisms have been elucidated (**Figure 1b**) (5, 9), including a putatively key role for the programming of offspring appetite via energy homeostasis brain networks (10). In humans, child appetite traits are associated with the child ‘s own BMI (11), and with maternal overweight/obesity (12). However, whether developmental programming of adiposity and appetite occurs in humans remains unclear. Mendelian randomization (13), sibling studies (14, 15) and paternal negative exposure control studies (16, 17) suggest that familial confounding (either genetic or non-genetic) may be an important cause of parent-child BMI associations. However, such associations are generally unchanged on adjustment for measured variables (18), leaving the specific confounders unidentified.

We aimed to establish whether associations between peri-pregnancy parental BMI and offspring birth weight, childhood BMI and appetite-related eating behaviours are due to genetic confounding. We first compared the strength of the associations of maternal versus paternal BMI with offspring outcomes, which are likely to be similarly strong if they are primarily due to genetic confounding. We then applied a genetically informed structural equation model (SEM) to a population-based sample of twins, siblings and half siblings, and their children, to quantify the relative importance of genetic confounding versus other mechanisms in underpinning intergenerational associations. Based on prior evidence (13-19) we hypothesized that genetic confounding would not explain the associations of parental BMI with offspring birth weight, but would be a major driver of associations with offspring childhood BMI.

## Methods

### Study design and participants

We analysed data from the Norwegian Mother, Father and Child Cohort Study (MoBa; described in detail elsewhere (20)), a prospective population-based birth cohort conducted by the Norwegian Institute of Public Health, and used data from the Medical Birth Registry of Norway (MBRN), a national health registry containing information about all births in Norway (21). Pregnant women were recruited at 50 out of 52 hospital maternity units in Norway, on attendance of a routine antenatal ultrasound scan offered to all Norwegian women at around 17 weeks of gestation. 41% of invitees participated, resulting in a total sample of around 114,500 children born between 1999 and 2009, along with around 95,200 mothers and 75,200 fathers. We used version 11 of the quality assured data files released for research in 2018, and analysed only live-born offspring. Flowcharts detailing sample selection are presented in **Supplementary information S1**. The establishment of MoBa and initial data collection was based on a license from the Norwegian Data Protection Agency and approval from The Regional Committees for Medical and Health Research Ethics. The MoBa cohort is currently regulated by the Norwegian Health Registry Act. The current study was approved by The Regional Committees for Medical and Health Research Ethics (REK 2013/863).

### Exposures and outcomes

The exposures were maternal pre-pregnancy BMI and paternal BMI during pregnancy, calculated from weight and height reported by the parents at the first study questionnaire (around 17 weeks gestation). Maternal height and pre-pregnancy weight were reported by the mothers, and paternal weight and height were reported by the fathers (for 35% of measurements), or by the mothers when paternal report was unavailable (Pearson ‘s r = 0.98 between maternally and paternally reported paternal weight and height).

Offspring outcomes included birth weight and BMI assessed between age 6 months and 8 years, and appetite-related eating behaviour traits assessed at age 8 years via the Child Eating Behaviour Questionnaire (CEBQ) (22). Birth weight and length were from the MBRN. Mothers completed regular questionnaires when their children were aged between 6 months and 8 years, from which the child ‘s weight and height at age 6 months and 1, 2, 3, 5, and 8 years were obtained. Measurements at ages up 3 years were predominantly from the child ‘s health card, whereas measurements from 5 years onwards were carried out by the parents. In order to maximise statistical efficiency we also used all available offspring BMI measurements to fit a growth curve, from which we predicted offspring BMI at 1 year intervals between age 1 and 8 years for children with at least three BMI measurements. These fitted BMI values, which we refer to as “predicted BMI”, were used as a supplement to the mother-reported BMI measures described above, enabling comparison of results in an identical (and larger) sample across different ages. Full details of the cleaning of anthropometric data and growth curve fitting are given in **Supplementary information S2**. As pre-planned secondary outcomes we analysed ponderal index (weight [kg]/length [m]^3^) and BMI at birth, and weight at ages up to 8 years.

The CEBQ is a widely used and validated psychometric instrument for child obesogenic eating behaviours (22). At the 8-year questionnaire, mothers completed 5-point Likert scales for 18 CEBQ items related to their child ‘s satiety responsiveness, slowness in eating, enjoyment of food, fussiness, emotional overeating and emotional undereating. We calculated the mean item score for each of the six scales for participants with available data for least two out of three items per scale. Covariate data were obtained from the MBRN or study questionnaires and are described in **Supplementary information S3**.

### Linear regression analyses

We fitted linear regression models to explore associations between exposures and outcomes, adjusting when relevant for offspring sex and age at outcome measurement, the other parent ‘s BMI, and potential non-genetic confounders including maternal parity, parental and grandparental language group (as a proxy for ethnicity) and maternal and paternal characteristics (age, smoking during pregnancy, educational attainment and income). Participants with non-missing values for all relevant variables were included in analyses. To account for non-independence between siblings we used a linear mixed model with a random intercept at the family level (**Supplementary information S4**), and a z-test was used to test whether associations with maternal and paternal BMI differed in magnitude (**Supplementary information S5**). For ease of interpretation, exposure and outcome variables were standardized, therefore regression coefficients are interpreted as the average change in the outcome in standard deviation (SD) units per 1 SD increase in the exposure. Because the standard deviation may differ for maternal versus paternal BMI we also tested for maternal-paternal differences using unstandardized exposures. Offspring BMI from age 5 years onwards was positively skewed (**Supplementary information S6**) so was natural log transformed, and several CEBQ eating behaviour scores were strongly skewed so were regressed on offspring age and sex followed by rank-based inverse normal transformation of the residuals. We carried out sensitivity analyses including 1) additionally adjusting birth weight models for gestational age at birth, 2) testing for non-linear associations (**Supplementary information S7**), 3) testing for interaction by offspring sex, and 4) testing for maternal BMI-paternal BMI interaction. Analyses were carried out in R version 4.0.3 (23).

### Genetically informed structural equation modelling

To quantify the extent to which exposure-outcome associations were due to genetic confounding, we fit an extended children of twins SEM (the Multiple Children of Twins and Siblings [MCoTS] model, described in **Supplementary information S8** and elsewhere (24)) in a subset of the MoBa sample. An extended pedigree including twins, siblings and half siblings in both the parental and offspring generations was identified within MoBa using data from the study questionnaires, genotyping, and linkage to the Norwegian Population Registry, the Norwegian Twin Registry and the MBRN (24). Our MCoTS model partitions the phenotypic covariance between exposures and outcomes into a part due to genetic confounding and a residual part (due to any causal effects and/or non-genetic confounding). Skewed exposure and outcome variables were transformed as for linear regression analyses, with the exception that parental BMI was also natural log transformed given the multivariate normality assumptions of SEM fit via maximum likelihood. Exposure variables were standardized to give unit variance and zero mean. Outcome variables were standardized (or inverse normalized for eating behaviour variables) within sex strata (or within age and sex strata for child BMI outcomes). Because the variance of exposures and outcome variables was close to one, covariances are approximately equal to Pearson ‘s correlation coefficients. Classic and extended twin studies suggest the presence of dominance genetic effects and absence of common environmental effects for adult BMI (8), but provide support for common environmental effects on birth weight and child BMI (25, 26). We therefore chose a priori to fit an MCoTS model that partitioned parental BMI variance into additive, dominance and non-shared environmental components (an ADE model) and partitioned offspring outcome variance into additive, common environmental and non-shared environmental components (an ACE model). In sensitivity analyses we fit ACE and AE models for parental BMI as well as stratifying analyses by offspring sex, fitting a liability threshold model for untransformed eating behaviour outcomes (**Supplementary information S9–11**), and refitting BW models having dropped offspring-generation twins (because monozygotic [MZ] twins may share a placenta and twins have lower BW than singletons, which could generate biases). Standard errors and 95% confidence intervals were calculated via bias corrected bootstrapping of the MCoTS model with 10,000 resamples. SEM were fit in R version 4.0.3 (OpenMx package version 2.18.1) (23, 27).

## Results

Statistics are for the sample used for linear regression analyses of birth weight (n = 85,866), aside from the other outcome variables, for which statistics are from the corresponding linear regression samples. Equivalent data for the 8-year BMI sample (n = 46,620) are presented in Supplementary information S12. SD: standard deviations, WHO: World Health Organization The number of offspring included in linear regression analyses varied by outcome, from 85,866 (74.9% of recruited sample) to 30,904 (27.0% of recruited sample) for analyses of birth weight and 2-year BMI respectively. **Supplementary information S1** shows the proportion of the sample with non-missing data for each analysis, and **Table 1** shows the characteristics of the study participants. There was statistical evidence for selective attrition, such that the sample used for analyses of 8-year BMI (n = 46,620) was more highly educated and had lower obesity prevalence and greater maternal age versus the baseline sample, but the magnitude of such differences was relatively small (**Supplementary information S12**).

**Table 1:** Characteristics of the parental and offspring generation of study participants (*n* = 85,866)

| Variable | Mean | SD | *n* | % |
| --- | --- | --- | --- | --- |
| Parental characteristics |  |  |  |  |
| Maternal BMI ($\text{kg}/\text{m}^2$) | 24.1 | 4.3 | | |
| Paternal BMI ($\text{kg}/\text{m}^2$) | 25.9 | 3.3 | | |
| Maternal WHO BMI category ($\text{kg}/\text{m}^2$) | | | | |
| <18.5 |  |  | 2560 | 3.0 |
| 18.5–24.9 |  |  | 56367 | 65.6 |
| 25–29.9 |  |  | 18727 | 21.8 |
| $\geq 30$ | | | 8212 | 9.6 |
| Paternal WHO BMI category ($\text{kg}/\text{m}^2$) | | | | |
| <18.5 |  |  | 197 | 0.2 |
| 18.5–24.9 |  |  | 37822 | 44.0 |
| 25–29.9 |  |  | 39120 | 45.6 |
| $\geq 30$ | | | 8727 | 10.2 |
| Parity (number of previous births) |  |  |  |  |
| 0 |  |  | 38736 | 45.1 |
| 1 |  |  | 30807 | 35.9 |
| 2 |  |  | 12795 | 14.9 |
| 3 |  |  | 2740 | 3.2 |
| 4+ |  |  | 788 | 0.9 |
| Maternal age at birth of child (years) |  |  |  |  |
| $\leq 19$ | | | 624 | 0.7 |
| 20–24 |  |  | 8029 | 9.4 |
| 25–29 |  |  | 28245 | 32.9 |
| 30–34 |  |  | 33814 | 39.4 |
| 35–39 |  |  | 13465 | 15.7 |
| $\geq 40$ | | | 1689 | 1.9 |
| Paternal age at birth of child (years) |  |  |  |  |
| $\leq 19$ | | | 180 | 0.2 |
| 20–24 |  |  | 3497 | 4.1 |
| 25–29 |  |  | 19167 | 22.3 |
| 30–34 |  |  | 33725 | 39.3 |
| 35–39 |  |  | 20734 | 24.1 |
| $\geq 40$ | | | 8563 | 10.1 |
| Maternal smoking during pregnancy |  |  |  |  |
| No |  |  | 79345 | 92.4 |
| Yes |  |  | 6521 | 7.6 |
| Paternal smoking during pregnancy |  |  |  |  |
| No |  |  | 65570 | 76.4 |
| Yes |  |  | 20296 | 23.6 |
| Maternal educational attainment |  |  |  |  |
| Incomplete upper 2° school |  |  | 199 | 0.2 |
| Upper 2° school |  |  | 1527 | 1.8 |
| High school/junior college |  |  | 23896 | 27.9 |
| University/college, 4 years |  |  | 36309 | 42.3 |
| University/college, >4 years |  |  | 22558 | 26.3 |
| Other |  |  | 1377 | 1.6 |
| Parental language |  |  |  |  |
| Norwegian |  |  | 76521 | 89.1 |
| Other |  |  | 9345 | 10.9 |
| Offspring characteristics |  |  |  |  |
| Gestational age (weeks) | 39.8 | 2.0 |  |  |
| Birth weight (g) | 3563 | 596 |  |  |
| 6 month BMI ($\text{kg}/\text{m}^2$) | 17.2 | 1.5 | | |
| Age at 6 month BMI measurement (months) | 5.8 | 0.5 |  |  |
| 1 year BMI ($\text{kg}/\text{m}^2$) | 17.0 | 1.4 | | |
| Age at 1 year BMI measurement (years) | 1.0 | 0.1 |  |  |
| 2 year BMI ($\text{kg}/\text{m}^2$) | 16.5 | 1.4 | | |
| Age at 2 year BMI measurement (years) | 2.0 | 0.2 |  |  |
| 3 year BMI ($\text{kg}/\text{m}^2$) | 16.1 | 1.5 | | |
| Age at 3 year BMI measurement (years) | 3.0 | 0.1 |  |  |
| 5 year BMI ($\text{kg}/\text{m}^2$) | 15.6 | 1.6 | | |
| Age at 5 year BMI measurement (years) | 5.2 | 0.3 |  |  |
| 8 year BMI ($\text{kg}/\text{m}^2$) | 16.1 | 2.0 | | |
| Age at 8 year BMI measurement (years) | 7.8 | 0.5 |  |  |

Linear regression analyses provided strong statistical evidence that the association of maternal BMI with offspring birth weight is stronger than that of paternal BMI with offspring birth weight (**Figure 2a**). However, after birth the associations with offspring BMI converged, and the associations of maternal and paternal BMI with offspring 2–5- year BMI were similar. Although for 8-year BMI there was statistical evidence that the paternal association was slightly weaker, the difference was not large, and when we used unstandardized parental variables the paternal association was actually slightly stronger than the maternal association (**Supplementary table S1**). These results were not markedly different when using offspring BMI predicted from a modelled growth curve (**Supplementary information S13**), when substituting birth weight for ponderal index/BMI at birth and substituting child BMI for weight, when stratifying by offspring sex, or when additionally adjusting for gestational age (**Supplementary table S1**). With respect to eating behaviour outcomes, both maternal and paternal BMI were positively associated with offspring food responsiveness and emotional overeating, and negatively associated with emotional undereating. Only paternal BMI was associated (negatively) with offspring satiety responsiveness and slow eating. Offspring 8- year BMI was associated with all eating behaviour outcomes except for emotional undereating, in the directions that would be expected from the behavioral susceptibility theory of obesity (22) (**Supplementary information S14**). We did not observe large departures from log-linear relationships (**Supplementary information S7**), and statistical interaction between maternal and paternal BMI was at most minor (**Supplementary information S15**).

**Figure 2:**
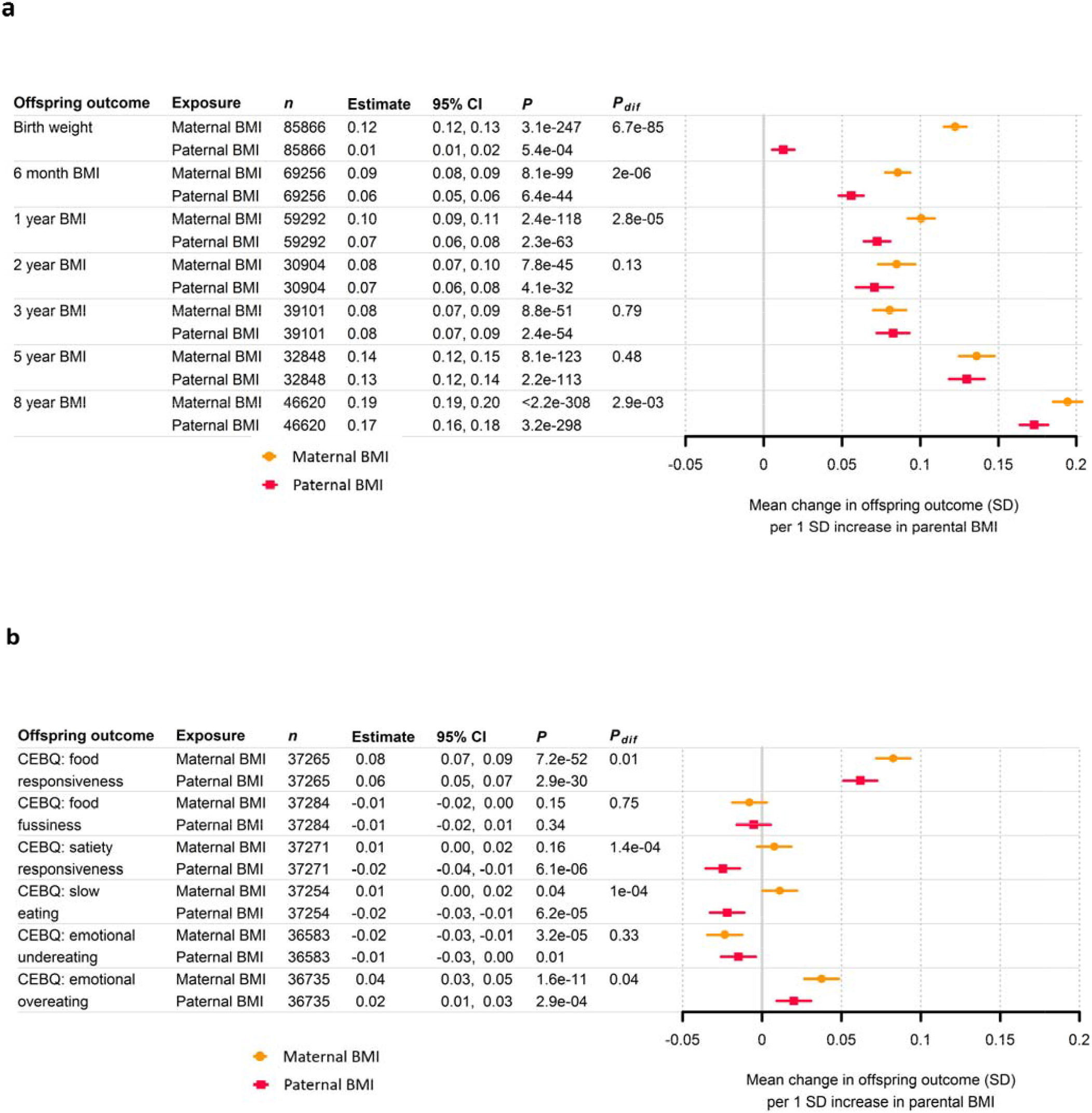
Linear associations of maternal and paternal BMI with offspring birth weight, child BMI and 8-year eating behaviours Linear associations of maternal and paternal BMI with offspring outcomes. a: offspring birth weight and child BMI, **b**: offspr*ing 8-* year old CEBQ eating behaviour traits, ***P***: *P*-value testing the null hypothesis that regression coefficient is zero, ***P***_**dif**_: *P*-*value* testing the null hypothesis that maternal and paternal regression coefficients are equal

**Table 2** shows the sample size available for MCoTS analyses, stratified by maternal and offspring relationship. The MCoTS results indicated that the positive phenotypic covariance between maternal BMI and offspring birth weight was not explained by genetic confounding, with genetic covariance estimates that were statistically indistinguishable from zero (**Figure 3**). The weak positive phenotypic covariance between paternal BMI and offspring birth weight was also not explained by genetic confounding. Surprisingly, there was statistical evidence for a small negative genetic covariance between paternal BMI and offspring birth weight, but this attenuated and became statistically indistinguishable from zero when, in exploratory analyses, we adjusted exposures and outcomes for potential confounders (including maternal BMI, paternal age and paternal income), suggesting that bias due to uncontrolled confounding may be the explanation (**Supplementary information S16**).

**Table 2:** Number of families available for MCoTS analyses, stratified by parental and offspring relationship

| Relationship type | Relatedness coefficient | n |  |  |  |
| --- | --- | --- | --- | --- | --- |
|  |  | Total | Parental exposure data available<sup>a</sup> | Offspring BW data available<sup>a</sup> | Offspring 8yr BMI data available<sup>a</sup> |
| Maternal BMI analyses |  |  |  |  |  |
| Extended families (n = 16292) |  |  |  |  |  |
| n stratified by mothers' relatedness in each extended family |  |  |  |  |  |
| MZ twin mother pair | 1.00 | 61 | 60 | 61 | 48 |
| DZ twin mother pair | 0.50 | 34<sup>b</sup> | 34 | 34 | 24 |
| Full sibling mother pair | 0.50 | 4916 | 4745 | 4910 | 3472 |
| Maternal half sibling mother pair | 0.25 | 376 | 355 | 375 | 218 |
| Unrelated sister-in-law mother pair | 0.00 | 10905 | 10495 | 10887 | 7633 |
| n offspring pairs linked to each mother<sup>c</sup> |  |  |  |  |  |
| Full sibling offspring pair | 0.50 | 4538 | 4477 | 4535 | 3154 |
| Maternal half-sibling offspring pair | 0.25 | 38 | 38 | 38 | 20 |
| Unpaired (single) offspring | ... | 28008 | 20205 | 22288 | 11689 |
| Unpaired nuclear families (n = 7339) |  |  |  |  |  |
| n offspring pairs linked to each mother |  |  |  |  |  |
| MZ twin offspring pair | 1.00 | 287 | 269 | 278 | 157 |
| DZ twin offspring pair | 0.50 | 738 | 698 | 735 | 379 |
| Full sibling offspring pair | 0.50 | 6229 | 6142 | 6227 | 4216 |
| Maternal half sibling offspring pair | 0.25 | 85 | 81 | 85 | 37 |
| Paternal BMI analyses |  |  |  |  |  |
| Extended families (n = 17036) |  |  |  |  |  |
| n stratified by fathers' relatedness in each extended family |  |  |  |  |  |
| MZ twin father pair | 1.00 | 27 | 27 | 27 | 24 |
| DZ twin father pair | 0.50 | 16<sup>b</sup> | 16 | 16 | 13 |
| Full sibling father pair | 0.50 | 3154 | 3127 | 3154 | 2462 |
| Paternal half sibling father pair | 0.25 | 171 | 168 | 171 | 128 |
| Unrelated brother-in-law father pair | 0.00 | 13668 | 13164 | 13603 | 9621 |
| n offspring pairs linked to each father<sup>c</sup> |  |  |  |  |  |
| Full sibling offspring pair | 0.50 | 5710 | 5570 | 5706 | 3861 |
| Paternal half sibling offspring pair | 0.25 | 28 | 28 | 28 | 22 |
| Unpaired (single) offspring | ... | 28334 | 22207 | 25046 | 12605 |
| Unpaired nuclear families (n = 7537) |  |  |  |  |  |
| n offspring pairs linked to each father |  |  |  |  |  |
| MZ twin offspring pair | 1.00 | 287 | 265 | 278 | 157 |
| DZ twin offspring pair | 0.50 | 951 | 854 | 924 | 453 |
| Full sibling offspring pair | 0.50 | 6229 | 6152 | 6227 | 4216 |
| Paternal half sibling offspring pair | 0.25 | 70 | 67 | 69 | 39 |

**Figure 3:**
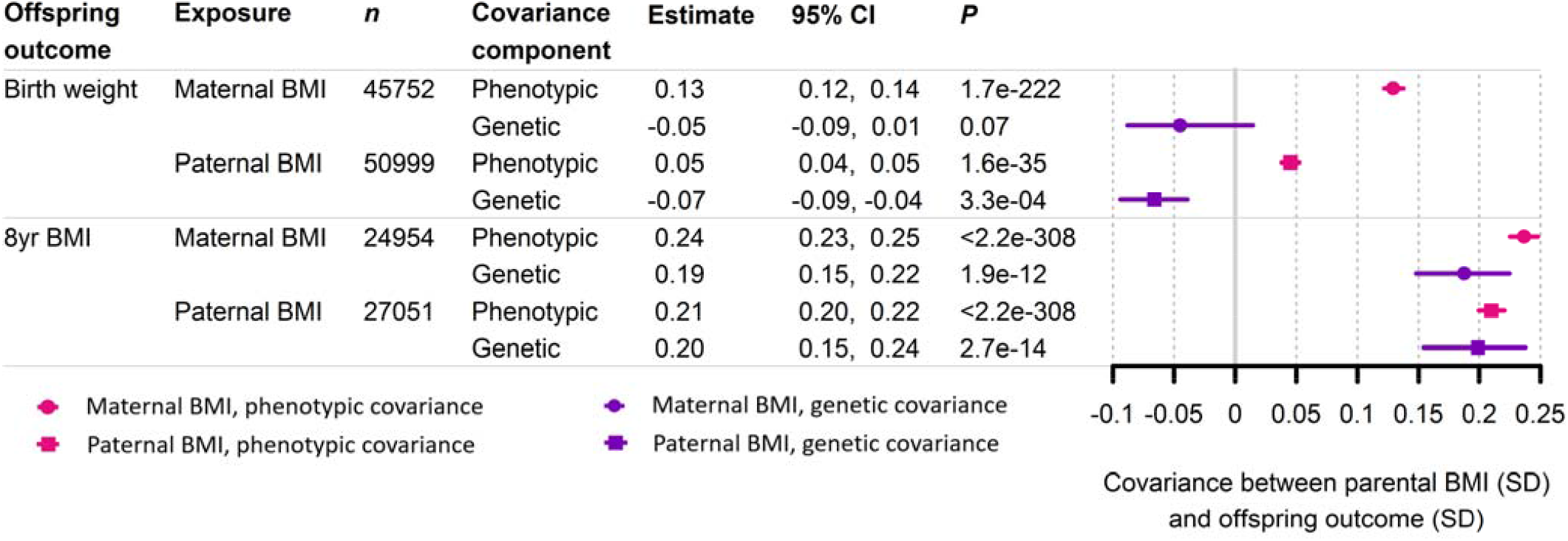
MCoTS SEM estimates of phenotypic and genetic covariance of parental BMI with offspring birth weight and 8yr B*MI* **Phenotypic covariance** denotes the overall covariance between the exposure and outcome, **Genetic covariance** denote*s the* part of the phenotypic covariance that is due to genetic confounding, ***P***: *P*-value for phenotypic covariance calculated via a z*-test* using the standard error from bootstrapping the MCoTS model, *P*-value for genetic covariance calculated via a chi square*d test* for deterioration of model fit on deletion of the a1’ path coefficient, ***n***: number of offspring with outcome data available

From age 6 months onwards, genetic covariance estimates became positive and increased in magnitude, such that for offspring 8-year BMI, genetic confounding explained 79% (0.19 / 0.24 * 100) (95% CI: 62%, 95%) of the covariance with maternal BMI and 94% (0.20 / 0.21 * 100) (95% CI: 72%, 113%) of the covariance with paternal BMI (**Figure 3, Supplementary information S17**). Genetic confounding explained a high and stable proportion of the phenotypic covariance with predicted BMI from age 1 to 8 years (**Figure 4, Supplementary information S18**). Results did not appreciably differ when we fitted ACE or AE models instead of the primary ADE model in the parent generation (**Supplementary Table 2**), when birth weight was substituted for ponderal index/BMI at birth and child BMI was substituted for weight (**Supplementary Table 2, Supplementary information S19**), or when BW models were refit without offspring-generation twins (results available from the authors on request). Although sex stratified models were underpowered, they provided no evidence for large sex differences in estimates (**Supplementary Table 2**). MCoTS models for eating behaviour outcomes were underpowered and uninformative (**Supplementary Table 2, Supplementary information S20**). Full MCoTS results including model fit statistics and estimated variance components for parental and offspring phenotypes are presented in **Supplementary Table 2**.

**Figure 4:**
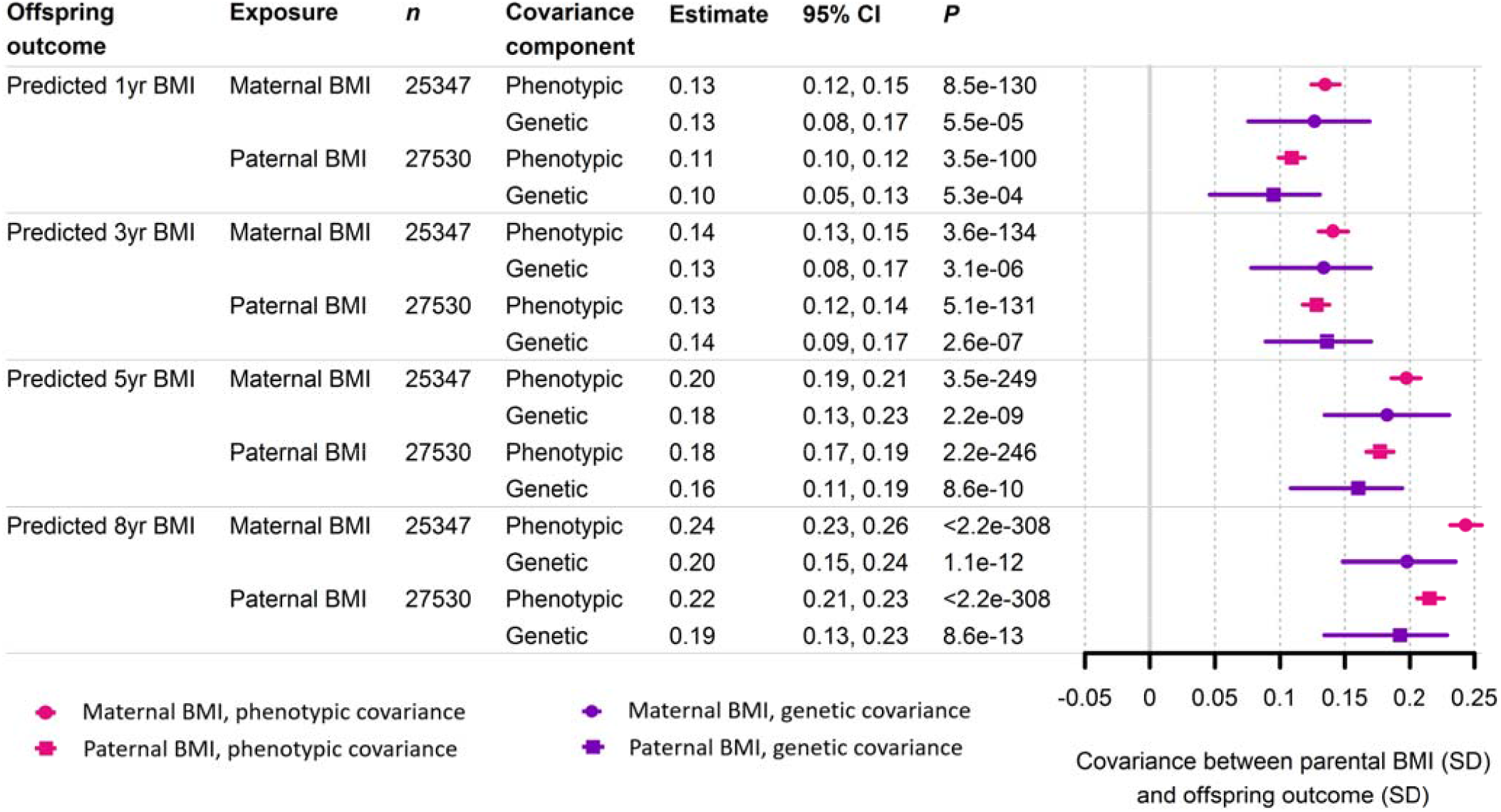
MCoTS SEM estimates of phenotypic and genetic covariance of parental BMI with offspring predicted BMI**Phenotypic covariance** denotes the overall covariance between the exposure and outcome, **Genetic covariance** denote*s the* part of the phenotypic covariance that is due to genetic confounding, ***P***: *P*-value for phenotypic covariance calculated via a z*-test* using the standard error from bootstrapping the MCoTS model, *P*-value for genetic covariance calculated via a chi square*d test* for deterioration of model fit on deletion of the a1’ path coefficient, ***n***: number of offspring with outcome data available

## Discussion

We triangulated evidence from two analytic approaches applied to a large European birth cohort, to infer the mechanisms underlying associations between parental BMI and offspring BW, BMI until 8 years and 8-year eating behaviour. There were not large differences in the magnitude of the associations of maternal and paternal BMI with offspring BMI beyond early childhood, suggesting confounding within families as the most parsimonious explanation for such associations. This was confirmed by our MCoTS analyses, which indicated that the covariance between parental BMI and offspring BMI from age 6 months to 8 years is primarily due to genetic confounding.

We have previously used genomic restricted maximum likelihood (GCTA-GREML) to explore whether intergenerational BMI associations could be due to genetic confounding (18). Those analyses investigated whether mother-offspring BMI covariance was explained by a set of ∼8 million imputed common genetic variants, in nominally unrelated individuals from 3 European birth cohorts. In contrast, the present MCoTS analyses investigated confounding involving all genetic variants, using expected relatedness inferred via quantitative genetic theory in a pedigree of close relatives. GCTA-GREML indicated that for maternal BMI, imputed variants explained 43% (95% CI: 15%, 72%) of the covariance with offspring 15-year BMI, with a similar estimate for 10-year BMI. This is highly consistent with the present MCoTS estimates: 79% (95% CI: 62%, 95%) of the covariance between maternal BMI and offspring 8-year BMI was explained by genetic confounding. MCoTS estimates were somewhat higher than those from GCTA-GREML, which is expected given that GCTA-GREML uses a set of measured genetic variants whereas MCoTS infers the effects of all variants. Furthermore, both GCTA-GREML and MCoTS indicated that the covariance between maternal BMI and offspring birth weight was not due to genetic confounding. The high concordance of results between the two methods, which make different assumptions and were applied to different cohorts, provides strong evidence that genetic confounding is a major driver of associations between parental BMI and offspring adiposity in late childhood.

Our results are consistent with previous maternal-paternal comparison, MR and sibling studies (several of which share authors with the present study), which have not supported a large causal effect of maternal BMI on offspring childhood adiposity (13, 14, 16, 17). However, sibling studies do provide a degree of support for potential causal effects of more extreme maternal metabolic dysregulation (for example maternal diabetes and severe obesity) on offspring adiposity beyond birth (2, 14). Taken together, this suggests that putative causal effects observed in animal models of developmental overnutrition (5, 9) should be interpreted cautiously and do not necessarily occur in humans. The association between maternal BMI and offspring size/adiposity at birth has been invoked in the literature to argue that maternal adiposity has a causal effect on offspring adiposity beyond birth (28). Indeed, MR and sibling studies (13, 19, 29, 30) support a causal effect of maternal BMI on offspring birth size. However, the present results, alongside previous studies (13, 17-19, 29), strongly suggest that the causes of weight at birth are somewhat different from the causes of BMI in later childhood.

We observed some associations between parental BMI and offspring obesity-related eating behaviours assessed via the CEBQ questionnaire. Previous studies in smaller samples have found similar associations, albeit somewhat inconsistently (12, 31). In particular, we found that greater maternal and paternal BMI were associated with increased scores on the CEBQ food responsiveness and emotional overeating scales, and a reduced score on the emotional undereating scale. Offspring satiety responsiveness and slow eating were only associated with paternal BMI; however, we cannot exclude measurement error as an explanation for the absent maternal associations because the CEBQ questionnaire was completed by mothers and is inherently subjective. Given our maternal-paternal comparison results it is plausible that child appetite-related eating behaviours mediate the genetically driven association between parent and child BMI. However, it was not possible to confirm this because our MCoTS analyses were underpowered for eating behaviours.

Our results have important public health implications, when considered alongside prior evidence. Maternal BMI is unlikely to have a large causal effect on child BMI beyond birth, although a small causal effect remains plausible, potentially mediated via maternal glycaemia during pregnancy. Any causal effect of paternal BMI on offspring childhood BMI is likely to be similar to or smaller than that of maternal BMI. Consequently, reductions in the BMI of either parent before pregnancy are unlikely to cause large reductions in childhood adiposity. It is possible that interventions targeting parental BMI reduction could influence childhood adiposity via parental lifestyle changes that persist after birth and affect the offspring ‘s environment. However, whether preconceptional interventions are the optimal approach for preventing childhood obesity requires further evaluation in light of evidence from the present study. Importantly, women considering pregnancy should still be advised and supported to maintain a healthy weight, because there is good evidence that maternal obesity in pregnancy causes adverse perinatal outcomes in the mother and offspring (32).

Our study has important strengths. We have analysed data from a large prospective birth cohort, enabling precise estimation of the associations between parental BMI assessed during pregnancy and offspring outcomes in mid childhood. We leveraged a pedigree involving twin and sibling relationships in both the parental and offspring generations, to partition parent-offspring phenotypic covariance via an MCoTS SEM that is rooted in quantitative genetics theory. We gained increased power over a conventional children of twins model by including non-twin siblings in the parent and offspring generations, and up to two children for each parent. We also acknowledge potential limitations of this study. First, because the MCoTS model cannot simultaneously estimate dominance genetic effects and common environmental effects, we fitted an ADE model in the parent generation, which assumes that common environmental effects are absent. This assumption is supported by classic and extended twin studies of adult BMI (8) and the similarity of our findings when we used ACE or AE models in the parent generation (**Supplementary table 2**). Second, our MCoTS analyses assume that phenotypic associations between log parent BMI and offspring outcomes are linear. In our data there were only mild deviations from log-linearity which would be unlikely to meaningfully alter our conclusions (**Supplementary information S7**). Third, our MCoTS analyses did not account for assortative mating. However, we do not expect this to have had a large impact on our results because spousal phenotypic correlations for BMI are relatively weak (13, 33), and spousal correlations at BMI associated loci are weaker still (13). Fourth, the MCoTS model does not account for gene by environment interaction, and if interactions exist between the additive genetic and common environmental variance components our results could overestimate genetic confounding. We believe any such bias is likely to be small though because extended twin family design data suggest common environmental effects are negligible for adult BMI (8). Fifth, the residual covariance estimated by the MCoTS model will not be indicative of the true causal effect of parental BMI, to the extent that residual confounding affects associations between parental BMI and offspring outcomes. It is likely that the negative residual covariance estimate for paternal BMI and offspring birth weight reflects residual confounding, particularly as this estimate attenuated on adjustment for potential confounders (**Supplementary information S15**). Sixth, our maternal-paternal comparisons did not account for non-paternity, which could weaken paternal associations. However, a previous simulation study showed that for a maternal-paternal comparison analysis using MoBa data with follow up to age three years, results would have changed little with non-paternity rates of up to 10% (17). Seventh, BMI is an imprecise proxy measure for adiposity. Despite this, BMI is highly correlated with more direct adiposity measures in childhood (34). Eighth, MoBa has a participation rate of 41% and there has been attrition over follow up (20). Although we cannot exclude an effect of selection bias on our results, we believe it unlikely that this would be of sufficient magnitude to alter our conclusions. Lastly, we have studied a Norwegian population which has relatively high obesity prevalence and income per capita in international terms and it would be beneficial to replicate our analyses in other settings.

In summary, we have shown that in a Norwegian population the linear association between parental BMI around the time of pregnancy and offspring BMI from age 6 months to 8 years is primarily due to genetic confounding. Our results suggest that neither the mothers ‘nor fathers’ pre-pregnancy BMI has a large causal effect on childhood BMI. This implies that any hypothetical intervention that successfully reduced parental BMI before pregnancy, without altering the offspring ‘s postnatal environment, would be insufficient to achieve large reductions in the offspring ‘s childhood obesity risk. Our results suggest that in the studied population, maintaining healthy parental BMI before conception is unlikely to be a promising target for childhood obesity prevention interventions.

## Contributors

Tom A Bond: conceptualisation, data analysis, software, data interpretation, literature search, writing- original draft Tom A McAdams: funding acquisition, data curation, methodology, software, data interpretation, writing- review and editing

Nicole M Warrington: methodology, software, data interpretation, writing- review and editing

Laurie J Hannigan: methodology, data curation, software, data interpretation, writing- review and editing Espen Moen Eilertsen: methodology, data curation, software, data interpretation, writing- review and editing Fartein A Torvik: methodology, data curation, software, data interpretation, writing- review and editing

Ziada Ayorech: data curation, software, data interpretation, writing- review and editing George Davey Smith: data interpretation, writing- review and editing

Deborah A Lawlor: data interpretation, writing- review and editing

Eivind Ystrom: funding acquisition, methodology, software, data interpretation, writing- review and editing Alexandra Havdahl: funding acquisition, conceptualisation, supervision, data interpretation, writing- review and editing

David M Evans: funding acquisition, conceptualisation, supervision, data interpretation, writing- review and editing

## Data Availability

Data from the Norwegian Mother, Father and Child Cohort Study and the Medical Birth Registry of Norway used in this study are managed by the national health register holders in Norway (Norwegian Institute of Public Health) and can be made available to researchers, subject to approval from the Regional Committees for Medical and Health Research Ethics (REC), compliance with the EU General Data Protection Regulation (GDPR) and approval from the data owners. The consent given by the participants does not cover storage of data on an individual level in repositories. Researchers who want access to data sets for replication should apply through helsedata.no. Access to data sets requires approval from The Regional Committee for Medical and Health Research Ethics in Norway and an agreement with MoBa.

https://helsedata.no

## Acknowledgements

The Norwegian Mother, Father and Child Cohort Study is supported by the Norwegian Ministry of Health and Care Services and the Ministry of Education and Research. We are grateful to all the participating families in Norway who take part in this on-going cohort study. TAB, GDS, DAL, NMW and DME work in or are affiliated a unit that receives support from the University of Bristol and UK Medical Research Council (MC_UU_00011/1 and MC_UU_00011/6). This study has received support from the British Heart Foundation Accelerator Award at the University of Bristol (AA/18/1/34219) and a University of Queensland-University of Exeter Accelerator grant. LJH was supported by the South-Eastern Norway Regional Health Authority (2018058, 2019097) during the completion of this work. NMW is funded by a NHMRC Investigator grant (APP2008723). DME is funded by an Australian National Health and Medical Research Council Senior Research Fellowship (APP1137714) and this work was funded by National Health and Medical Research Council (Australia) (NHMRC) project grants (GNT1157714, GNT1183074). FAT was partly supported by the Research Council of Norway through its Centers of Excellence funding scheme (262700). ZA is funded by a Marie Skłodowska-Curie Fellowship from the European Research Council (894675). TAM is supported by a Wellcome Trust Senior Research Fellowship (220382/Z/20/Z). AH was supported by grants from the Norwegian Research Council (274611, 3006668) and the South-Eastern Norway Regional Health Authority (2018059, 2020022). EY was supported by the Research Council of Norway (288083). This paper is the work of the authors and does not necessarily represent the views of individuals or organizations acknowledged here.

