## Supplementary information for "Parental body mass index and offspring childhood body size and eating behaviour: causal inference via parental comparisons and extended children of twins structural equation modelling"

Tom A Bond[1,2,3,4,*], Tom A McAdams[5,6], Nicole M Warrington[1,2,7,8], Laurie J Hannigan[2,9,10], Espen Moen Eilertsen[6,11,12], Ziada Ayorech[6], Fartein A Torvik[6,12], George Davey Smith[2,3], Deborah A Lawlor[2,3], Eivind Ystrom[6,10], Alexandra Havdahl[2,6,9,10,§], David M Evans[1,2,7,§]

[1]The University of Queensland Diamantina Institute, The University of Queensland, Brisbane, Australia.
[2]MRC Integrative Epidemiology Unit at the University of Bristol, Bristol, UK.
[3]Population Health Sciences, Bristol Medical School, University of Bristol, Bristol, UK.
[4]Department of Epidemiology and Biostatistics, Imperial College London, London, UK.
[5]Social, Genetic and Developmental Psychiatry Centre, Institute of Psychiatry, Psychology and Neuroscience, King's College, London, UK.
[6]Department of Psychology, PROMENTA Research Center, University of Oslo, Oslo, Norway.
[7]Institute for Molecular Bioscience, University of Queensland, Brisbane, Australia.
[8]K.G. Jebsen Center for Genetic Epidemiology, Department of Public Health and Nursing, NTNU, Norwegian University of Science and Technology, Norway.
[9]Nic Waals Institute, Lovisenberg Diakonale Hospital, Oslo, Norway.
[10]Department of Mental Disorders, Norwegian Institute of Public Health, Oslo, Norway.
[11]Department of Psychology, University of Oslo, Forskningsveien 3A, 0373, Oslo, Norway.
[12]Centre for Fertility and Health, Norwegian Institute of Public Health, Oslo, Norway.

*Corresponding author
§These authors contributed equally to this work

**Contents**

**Supplementary information S1: Sample selection flowchart** 3
**Supplementary information S2: Anthropometric data cleaning and growth curve fitting** 3
**Supplementary information S3: Covariate data** 5
**Supplementary information S4: Linear mixed model to account for non-independence between siblings** 6
**Supplementary information S5: *z*-test for the difference in maternal and paternal associations** 6
**Supplementary information S6: Summary statistics for transformed and untransformed exposure and outcome variables** 7
**Supplementary information S7: Tests for non-linear associations between exposures and outcomes** 8
**Supplementary information S8: Multiple Children of Twins and Siblings (MCoTS) model** 30
**Supplementary information S9: MCoTS model with ACE partition of parental exposure** 32
**Supplementary information S10: MCoTS model with AE partition of parental exposure** 33
**Supplementary information S11: Liability threshold model for untransformed CEBQ outcomes** 34
**Supplementary information S12: Differences in participant characteristics between baseline and 8 year old sample** 35
**Supplementary information S13: Linear associations between parental BMI and offspring predicted BMI** 36
**Supplementary information S14: Linear associations between offspring 8 year BMI and CEBQ outcomes** 36
**Supplementary information S15: Statistical interaction between maternal and paternal BMI** 37
**Supplementary information S16: MCoTS results for the association of parental BMI with offspring birth weight, adjusted for potential confounders** 38
**Supplementary information S17: MCoTS results for the association of parental BMI with offspring birth weight and BMI** 39
**Supplementary information S18: MCoTS results for the association of parental BMI with offspring predicted BMI** 40
**Supplementary information S19: MCoTS results for the association of parental BMI with offspring weight, BMI and ponderal index at birth** 41
**Supplementary information S20: MCoTS results for the association of parental BMI with offspring eating behaviour (CEBQ) traits** 42
**References** 42

**Supplementary information S1: Sample selection flowchart**

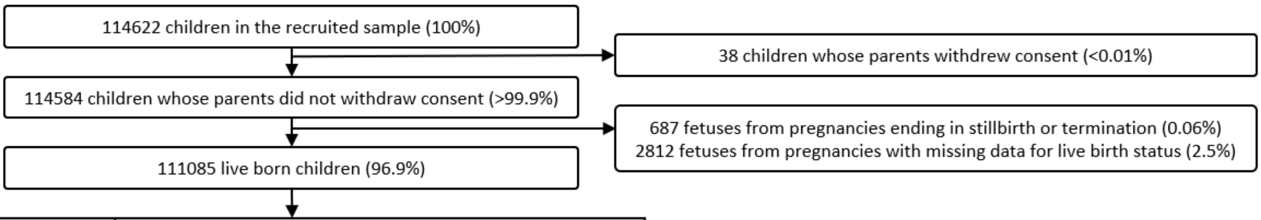


| Outcome | *n* children with available data for all necessary variables | | |
|---|---|---|---|
| | Linear regression analyses | MCoTS analyses (maternal BMI) | MCoTS analyses (paternal BMI) |
| Birth weight | 85866 (74.9%) | 45752 (39.9%) | 50999 (44.5%) |
| 6 month BMI | 69256 (60.4%) | 35913 (31.3%) | 39529 (34.5%) |
| 1 year BMI | 59292 (51.7%) | 31100 (27.1%) | 34210 (29.8%) |
| 2 year BMI | 30904 (27.0%) | 17001 (14.8%) | 18653 (16.3%) |
| 3 year BMI | 39101 (34.1%) | 21512 (18.8%) | 23319 (20.3%) |
| 5 year BMI | 32848 (28.7%) | 18194 (15.9%) | 19393 (16.9%) |
| 8 year BMI | 46620 (40.7%) | 24954 (21.8%) | 27051 (23.6%) |
| Predicted BMI (all ages) | 47524 (41.5%) | 25347 (22.1%) | 27530 (24.0%) |
| CEBQ: food responsiveness | 37265 (32.5%) | 20074 (17.5%) | 21573 (18.8%) |
| CEBQ: fussiness | 37284 (32.5%) | 20089 (17.5%) | 21587 (18.8%) |
| CEBQ: satiety responsiveness | 37371 (32.6%) | 20077 (17.5%) | 21576 (18.8%) |
| CEBQ: slow eating | 37254 (32.5%) | 20071 (17.5%) | 21570 (18.8%) |
| CEBQ: emotional undereating | 36583 (31.9%) | 19713 (17.2%) | 21175 (18.5%) |
| CEBQ: emotional overeating | 36735 (32.0%) | 19807 (17.3%) | 21269 (18.6%) |

**Supplementary information S2: Anthropometric data cleaning and growth curve fitting**

For all anthropometric variables, we set values that were biologically implausible to missing. Biological implausibility was determined by manual examination of the distribution of weight, height, BMI and ponderal index at each age and, where relevant, by comparison of values to those at earlier and later timepoints. Implausible weight and height values were excluded prior to calculation of BMI and ponderal index.

We analysed the children's mother-reported BMI at age 6 months and 1, 2, 3, 5, and 8 years. Analyses involving mother-reported BMI measurements had different sample sizes for each age due to missing data (either for whole questionnaires or for individual questions within questionnaires). Furthermore, analyses involving mother-reported BMI measurements made inefficient use of the available data, because each mother-reported measurement was considered in isolation without accounting for the child's other measurements, and sometimes children had multiple measurements available at similar ages, of which only one was used. In order to maximise statistical efficiency we therefore used all available offspring BMI measurements to fit a growth curve, from which we predicted offspring BMI at 1 year intervals between age 1 and 8 years. These fitted BMI values, which we refer to as "predicted BMI", were used as outcomes as a supplement to the mother-reported BMI measures described above, enabling comparison of linear regression and MCoTS results from an identical (and larger) sample across different ages.

Children with at least one BMI measurement taken between 1 and 8 years of age were included in the sample used to fit the growth curve. BMI across childhood has a complex shape; there is a steep increase from birth to around 9 months of age (which is referred to as the adiposity peak), then a decline until around 6 years of age (the adiposity rebound) before increasing through puberty. Due to this complex shape, measurements taken before age one were not used as several repeated measures in the first year of life are required to accurately model the adiposity peak

(1). In total, 323,372 measurements from 80,805 individual children (48.8% female) were used, and the mean number of measurements per child was 4.00. We did not natural log transform BMI measurements prior to fitting the growth curve because BMI was only slightly skewed from age five onwards. We fit a linear mixed model, separately in males and females with linear, quadratic and cubic terms for age in both the fixed and random effects to account for nonlinearity of BMI change over time, using the R package nlme (2, 3). The random effects allow the intercept and slope to vary around the population mean for each individual, and appropriately account for clustering of BMI measurements within individual children. Prior to model fitting, age was centered at four years (the mean age in the data). To ensure we had the best fitting model, we varied both the fixed and random effects and the correlation structure for the residuals then compared the models using the Akaike Information Criterion (AIC), Bayesian Information Criterion (BIC) and likelihood ratio tests. We established that (i) linear, quadratic and cubic fixed and random effects were required to adequately describe the growth curve, and (ii) a first order continuous autoregressive (AR1) correlation structure for the residuals gave better fit than a compound symmetry structure or unstructured residuals. Therefore, the final model for the BMI of the $i^{th}$ individual at the $t^{th}$ time point (age) was:

$$BMI_{it} = \beta 0 + u0_i + \beta 1 age_{it} + u1_i age_{it} + \beta 2 age_{it}^2 + u2_i age_{it}^2 + \beta 3 age_{it}^3 + u3_i age_{it}^3 + e_{it}$$

where $\beta 0$, $\beta 1$, $\beta 2$ and $\beta 3$ are the fixed coefficients representing the average intercept and linear, quadratic and cubic age effects respectively. $u0_i$, $u1_i$, $u2_i$ and $u3_i$ are the random coefficients representing the deviation for individual $i$ from the average intercept and linear, quadratic and cubic age effects respectively, with $u_i \sim N(\mathbf{0}, \boldsymbol{G})$ where $\boldsymbol{G}$ is the variance-covariance matrix of the random effects. $e_{it}$ is a normally distributed residual capturing the deviation of the $i^{th}$ individual at the $t^{th}$ time point from the individual growth curve, $e_{it} \sim N(\mathbf{0}, \mathbf{\Sigma})$ where $\mathbf{\Sigma}$ is the residual correlation matrix and a first order continuous autoregressive (AR1) correlation structure was assumed. The model accounts for missing data on the assumption that data are missing at random.

We used the growth curve to calculate predicted BMI (i.e. the fitted values from the linear mixed model) at one year age intervals between one and eight years, separately for male and female offspring, for children with at least three BMI measurements available. From age five onwards, predicted BMI variables were slightly skewed so were natural log transformed prior to subsequent analysis.

**Supplementary information S3: Covariate data**

MoBa mothers and fathers each completed a questionnaire at around 17 weeks of gestation (which are referred to here as “Questionnaire 1”, and “Father Questionnaire”). MoBa questionnaire data were linked to the Medical Birth Registry of Norway (MBRN) version 2017Q4 (4), which contained data from the birth notification forms (99.9% of pregnancies), or notification forms related to termination of pregnancy (0.1% of pregnancies). The questionnaire- and MBRN-derived variables used in the present study are detailed in the table below.

| Variable | Data source | Categories | Notes |
|---|---|---|---|
| Maternal parity | MBRN | “zero”, “one”, “two”, “three”, “four or more” | The highest value of the separate variables “parity registered by MBRN” and “parity reported by the mother” was used |
| Maternal smoking during pregnancy | Questionnaire 1 | “yes, “no” | |
| Paternal smoking during pregnancy | Paternal questionnaire, Questionnaire 1 | “yes, “no” | Maternal report was substituted for missing paternal report data for 23.9% of fathers |
| Maternal educational attainment | Questionnaire 1 | “Incomplete 9-year secondary school”, “9-year secondary school”, “1-2 year high school”, “vocational high school”, “3-year high school general studies/junior college”, regional technical college/4-year university degree”, “university/technical college, more than 4 years”, “other” | |
| Paternal educational attainment | Paternal questionnaire, Questionnaire 1 | “Incomplete 9-year secondary school”, “9-year secondary school”, “1-2 year high school”, “vocational high school”, “3-year high school general studies/junior college”, regional technical college/4-year university degree”, “university/technical college, more than 4 years”, “other” | Maternal report was substituted for missing paternal report data for 23.6% of fathers |
| Parental language | Questionnaire 1 | “Norwegian”, “other” | |
| Grandparental language | Questionnaire 1 | “Norwegian”, “other” | |
| Maternal income | Questionnaire 1 | “no income”, <150,000 NOK”, “150,000-199,999 NOK”, “200,000-299,999 NOK”, “300,000-399,999 NOK”, “400,000-499,999 NOK”, >500,000 NOK " | |
| Paternal income | Paternal questionnaire, Questionnaire 1 | “no income”, <150,000 NOK”, “150,000-199,999 NOK”, “200,000-299,999 NOK”, “300,000-399,999 NOK”, “400,000-499,999 NOK”, >500,000 NOK " | Maternal report was substituted for missing paternal report data for 64.3% of fathers |
| Maternal age at childbirth | MBRN | “≤17 years”, 18-19 years”, “20-24 years”, “20-25 years”, “25-29 years”, “30-34 years”, “35-39 years”, “40-44 years”, 45+ years” | |
| Paternal age at childbirth | MBRN | “≤19 years”, 20-24 years”, “25-29 years”, “30-34 years”, “35-39 years”, “40-44 years”, “45-49 years”, 50+ years” | |
| Offspring gestational age at birth | MBRN | NA | Based on ultrasound estimation. If ultrasound was not available, the gestational age was calculated from the last menstrual period |
| Liveborn status | MBRN | “liveborn”, “stillborn/termination” | |
| Offspring sex | MBRN | “female”, “male” | |

**NOK**: Norwegian Krone

### Supplementary information S4: Linear mixed model to account for non-independence between siblings

In MoBa there are a large number of siblings in the offspring generation (if siblings were excluded, the sample would be around 17% smaller). In order to maximise power we included siblings in the sample used for linear regression analyses, and used a linear mixed model with a random intercept at the family level to appropriately account for non-independence between siblings:

$$Y_{ij} = \beta 0 + u0_j + \beta 1 \times maternal\ BMI_{ij} + \beta 2 \times paternal\ BMI_{ij} + \sum_l \beta_l \times covariate_{lij} + e_{ij} \quad (1)$$

where $Y_{ij}$ is the phenotype for the $i^{th}$ individual in the $j^{th}$ family, $\beta 0$, $\beta 1$, $\beta 2$ and $\beta_l$ are the fixed coefficients for the intercept, maternal BMI, paternal BMI and the $l^{th}$ covariate respectively, $u0_j$ is a random intercept for the $j^{th}$ family, $u_j \sim N(0, \sigma_u^2)$, and $e_{ij}$ is a residual, $e_{ij} \sim N(\mathbf{0}, \mathbf{\Sigma})$. $\mathbf{\Sigma} = \boldsymbol{I}\sigma_e^2$ is the residual variance-covariance matrix, where $\boldsymbol{I}$ is an identity matrix and $\sigma_e^2$ is the residual variance. Linear mixed models were fitted using the R package nlme (1, 2).

### Supplementary information S5: *z*-test for the difference in maternal and paternal associations

For each offspring outcome we used a z-test to test whether associations with maternal and paternal BMI differed in magnitude. We calculated the *z* statistic as $z = \delta / \sqrt{Var(\beta 1) + Var(\beta 2) - 2Cov(\beta 1, \beta 2)}$, where $\delta$ is the difference between the coefficients for maternal BMI and paternal BMI ($\beta 1$ and $\beta 2$ respectively in equation 1), and $Cov(\beta 1, \beta 2)$ is from the estimated covariance matrix for the fixed effects in equation 1.

**Supplementary information S6: Summary statistics for transformed and untransformed exposure and outcome variables**

| Variable | Mean | SD | Minimum | Maximum | Skew | Kurtosis |
|---|---|---|---|---|---|---|
| Maternal BMI (z-score) | 0.00 | 1.00 | -2.68 | 7.84 | 1.42 | 3.02 |
| Log maternal BMI (z-score)[a] | 0.00 | 1.00 | -3.86 | 5.38 | 0.81 | 0.85 |
| Paternal BMI (z-score) | 0.00 | 1.00 | -3.82 | 10.14 | 1.02 | 3.10 |
| Log paternal BMI (z-score) [a] | 0.00 | 1.00 | -5.38 | 6.79 | 0.44 | 0.97 |
| Birth weight (sex standardised z-score)[a] | 0.01 | 0.98 | -5.30 | 4.61 | -0.74 | 2.34 |
| Birth BMI (sex standardised z-score)[a] | 0.01 | 0.98 | -5.96 | 5.24 | -0.50 | 1.96 |
| Birth PI (sex standardised z-score)[a] | 0.01 | 0.99 | -4.85 | 11.91 | 0.32 | 2.81 |
| 6 month weight (age/sex standardised z-score)[a] | 0.00 | 1.00 | -5.57 | 6.21 | 0.26 | 0.70 |
| 6 month BMI (age/sex standardised z-score)[a] | 0.00 | 1.00 | -4.28 | 6.25 | 0.45 | 0.58 |
| 1 year weight (age/sex standardised z-score)[a] | 0.00 | 1.00 | -5.56 | 5.84 | 0.35 | 0.52 |
| 1 year BMI (age/sex standardised z-score)[a] | 0.00 | 1.00 | -5.54 | 5.54 | 0.35 | 0.47 |
| 2 year weight (age/sex standardised z-score)[a] | 0.00 | 1.00 | -4.19 | 5.64 | 0.30 | 0.51 |
| 2 year BMI (age/sex standardised z-score)[a] | 0.00 | 1.00 | -4.29 | 5.52 | 0.31 | 0.70 |
| 3 year weight (age/sex standardised z-score)[a] | 0.00 | 1.00 | -3.79 | 8.59 | 0.48 | 0.99 |
| 3 year BMI (age/sex standardised z-score)[a] | 0.00 | 1.00 | -4.38 | 5.94 | 0.45 | 1.29 |
| 5 year weight (age/sex standardised z-score) | 0.00 | 1.00 | -3.78 | 6.77 | 0.74 | 1.76 |
| Log 5 year weight (age/sex standardised z-score)[a] | 0.00 | 1.00 | -5.42 | 4.93 | 0.17 | 0.74 |
| 5 year BMI (age/sex standardised z-score) | 0.00 | 1.00 | -4.08 | 6.52 | 0.69 | 2.12 |
| Log 5 year BMI (age/sex standardised z-score)[a] | 0.00 | 1.00 | -5.33 | 5.13 | 0.17 | 1.33 |
| 8 year weight (age/sex standardised z-score) | 0.00 | 1.00 | -3.24 | 8.82 | 1.02 | 2.21 |
| Log 8 year weight (age/sex standardised z-score)[a] | 0.00 | 1.00 | -4.97 | 5.78 | 0.38 | 0.57 |
| 8 year BMI (age/sex standardised z-score) | 0.00 | 1.00 | -3.53 | 7.31 | 0.97 | 2.29 |
| Log 8 year BMI (age/sex standardised z-score)[a] | 0.00 | 1.00 | -4.67 | 5.50 | 0.43 | 1.00 |
| 1 year predicted BMI (sex standardised z-score)[a] | -0.01 | 1.04 | -5.03 | 5.73 | 0.34 | 0.52 |
| 2 year predicted BMI (sex standardised z-score)[a] | 0.00 | 1.04 | -4.17 | 5.62 | 0.32 | 0.46 |
| 3 year predicted BMI (sex standardised z-score)[a] | 0.00 | 1.04 | -4.27 | 6.23 | 0.36 | 0.58 |
| 4 year predicted BMI (sex standardised z-score)[a] | 0.00 | 1.04 | -4.12 | 6.84 | 0.45 | 0.82 |
| 5 year predicted BMI (sex standardised z-score) | 0.00 | 1.05 | -4.26 | 7.01 | 0.62 | 1.35 |
| Log 5 year predicted BMI (sex standardised z-score)[a] | -0.01 | 1.05 | -5.23 | 5.80 | 0.31 | 0.73 |
| 6 year predicted BMI (sex standardised z-score) | 0.00 | 1.05 | -4.12 | 6.93 | 0.81 | 1.97 |
| Log 6 year predicted BMI (sex standardised z-score)[a] | -0.01 | 1.05 | -5.02 | 5.70 | 0.43 | 1.04 |
| 7 year predicted BMI (sex standardised z-score) | 0.00 | 1.06 | -3.99 | 7.63 | 0.94 | 2.38 |
| Log 7 year predicted BMI (sex standardised z-score)[a] | -0.01 | 1.06 | -4.92 | 6.10 | 0.51 | 1.21 |
| 8 year predicted BMI (sex standardised z-score) | 0.00 | 1.07 | -3.91 | 7.70 | 0.98 | 2.60 |
| Log 8 year predicted BMI (sex standardised z-score)[a] | -0.01 | 1.07 | -4.90 | 6.07 | 0.51 | 1.26 |
| CEBQ satiety responsiveness (age/sex standardised z-score) | 0.00 | 1.00 | -3.42 | 3.27 | 0.06 | -0.06 |
| CEBQ satiety responsiveness (inverse normalised z-score)[a, b] | 0.00 | 1.00 | -3.91 | 4.08 | 0.00 | 0.00 |
| CEBQ slow eating (age/sex standardised z-score) | 0.00 | 1.00 | -2.02 | 4.11 | 0.57 | 0.14 |
| CEBQ slow eating (inverse normalised z-score)[a, b] | 0.00 | 1.00 | -4.08 | 4.08 | 0.01 | -0.02 |
| CEBQ food responsiveness (age/sex standardised z-score) | 0.00 | 1.00 | -1.23 | 5.84 | 1.88 | 4.01 |
| CEBQ food responsiveness (inverse normalised z-score)[a, b] | 0.00 | 0.99 | -3.81 | 4.07 | 0.02 | 0.01 |
| CEBQ fussiness (age/sex standardised z-score) | 0.00 | 1.00 | -2.17 | 2.80 | 0.16 | -0.44 |
| CEBQ fussiness (inverse normalised z-score)[a, b] | 0.00 | 1.00 | -3.91 | 4.07 | 0.01 | -0.04 |
| CEBQ emotional overeating (age/sex standardised z-score) | 0.00 | 1.00 | -1.73 | 7.70 | 1.36 | 1.63 |
| CEBQ emotional overeating (inverse normalised z-score)[a, b] | 0.00 | 0.99 | -3.74 | 4.06 | 0.01 | 0.02 |
| CEBQ emotional undereating (age/sex standardised z-score) | 0.00 | 1.00 | -1.97 | 3.25 | 0.36 | -0.44 |
| CEBQ emotional undereating (inverse normalised z-score)[a, b] | 0.00 | 1.00 | -3.91 | 4.06 | 0.02 | -0.04 |

**a**: variable was used in genetically informed structural equation modelling (MCoTS) analyses (variables used in linear regression analyses were identical, with the exception that unlogged parental BMI was used as the exposure, and for models involving weight/BMI/PI, age and sex were included as covariates rather than using age/sex standardized z-scores), **b**: CEBQ variables were regressed on age and sex prior to inverse normalization of the residuals within sex strata

**Supplementary information S7: Tests for non-linear associations between exposures and outcomes**

Our MCoTS structural equation models partitioned the covariance, which is a measure of linear association, between the natural logarithm of the exposure, and the outcome (which was logged when necessary). To assess whether a linear association adequately described the relationship between each logged exposure and outcome we (i) explored whether there was evidence for a quadratic association, (ii) fitted a LOWESS smoother, and (iii) regressed the outcome on categories of log parental BMI equivalent to the World Health Organization obesity classification (<18.5 kg/m$^2$, 18.5–24.9 kg/m$^2$, 25–29.9 kg/m$^2$, 30–34.9 kg/m$^2$, 35–39.9 kg/m$^2$, ≥40 kg/m$^2$). Results are shown in the plots below, with vertical grey lines indicating the log BMI categories used for the categorical fit. Variables were treated as per the MCoTS analyses, i.e. exposures were logged and outcomes were logged when necessary. **INT:** inverse normal transformation.

Although there was some evidence for a non-linear association between log maternal BMI and offspring outcomes at younger ages (birth to three years), such that the association plateaued at higher maternal BMI, these departures from linearity were relatively mild, as demonstrated by the LOWESS and categorical fits. For log offspring BMI from age five onwards the associations with log maternal BMI were approximately linear. For log paternal BMI there was also evidence for a slight plateauing of the association with birth weight at higher paternal BMI, but associations with offspring BMI at ages beyond birth were approximately linear.

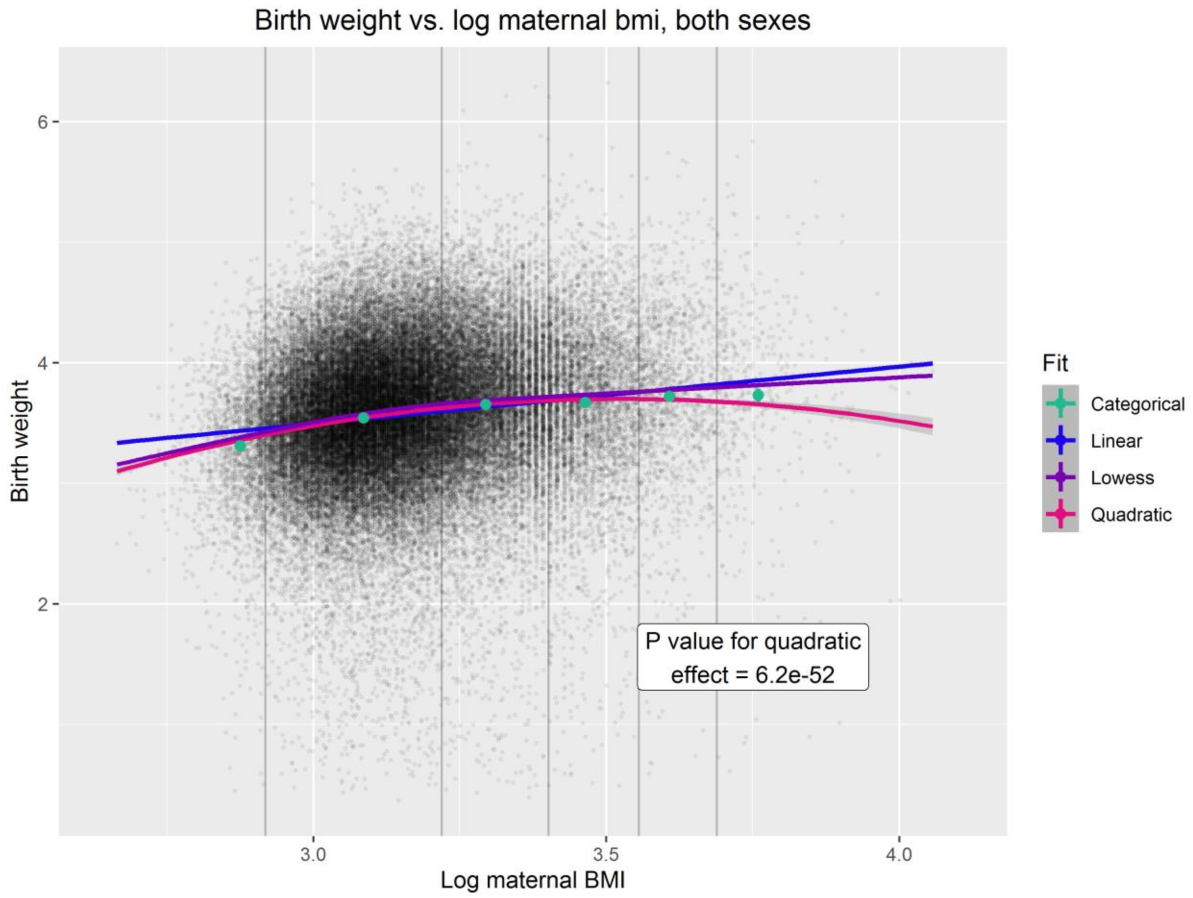

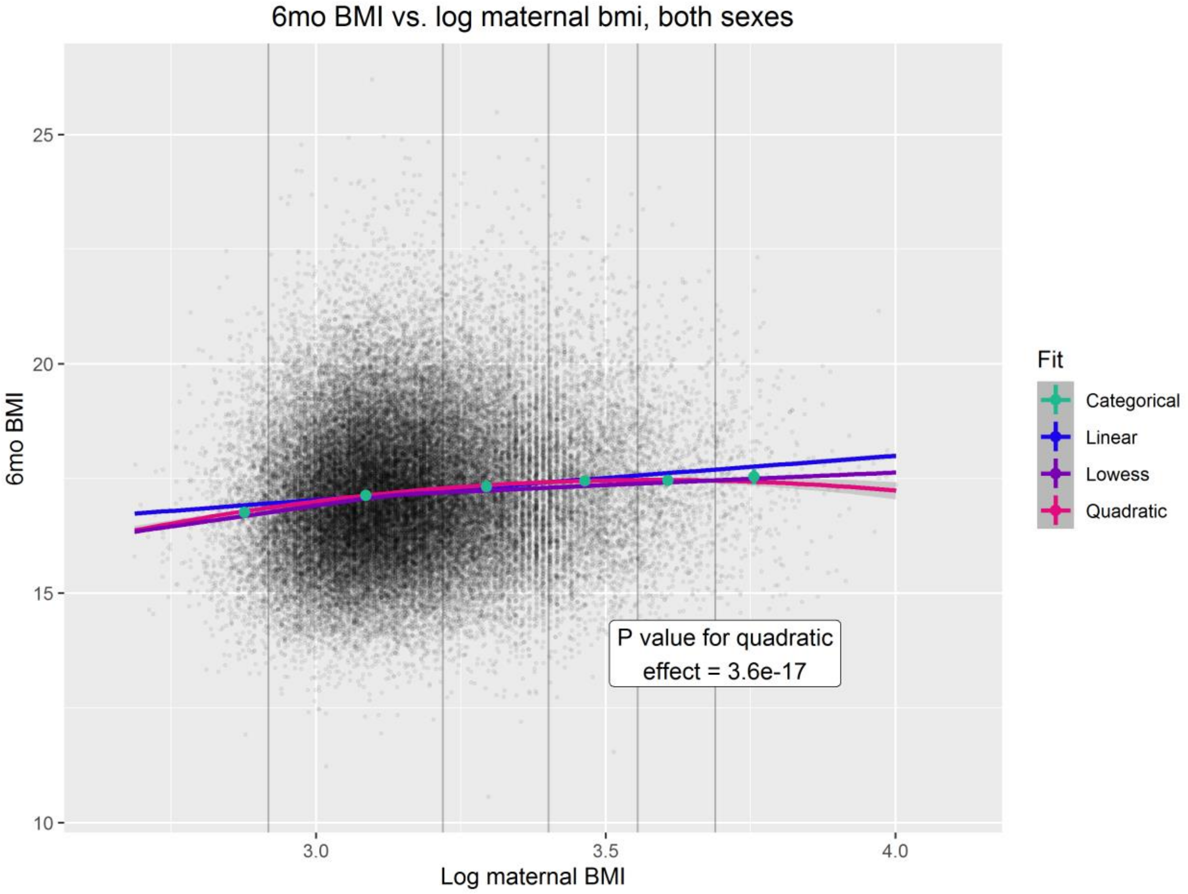
6mo BMI vs. log maternal bmi, both sexes
6mo BMI
Log maternal BMI
25
20
15
10
3.0
3.5
4.0
Fit
Categorical
Linear
Lowess
Quadratic
P value for quadratic effect = 3.6e-17

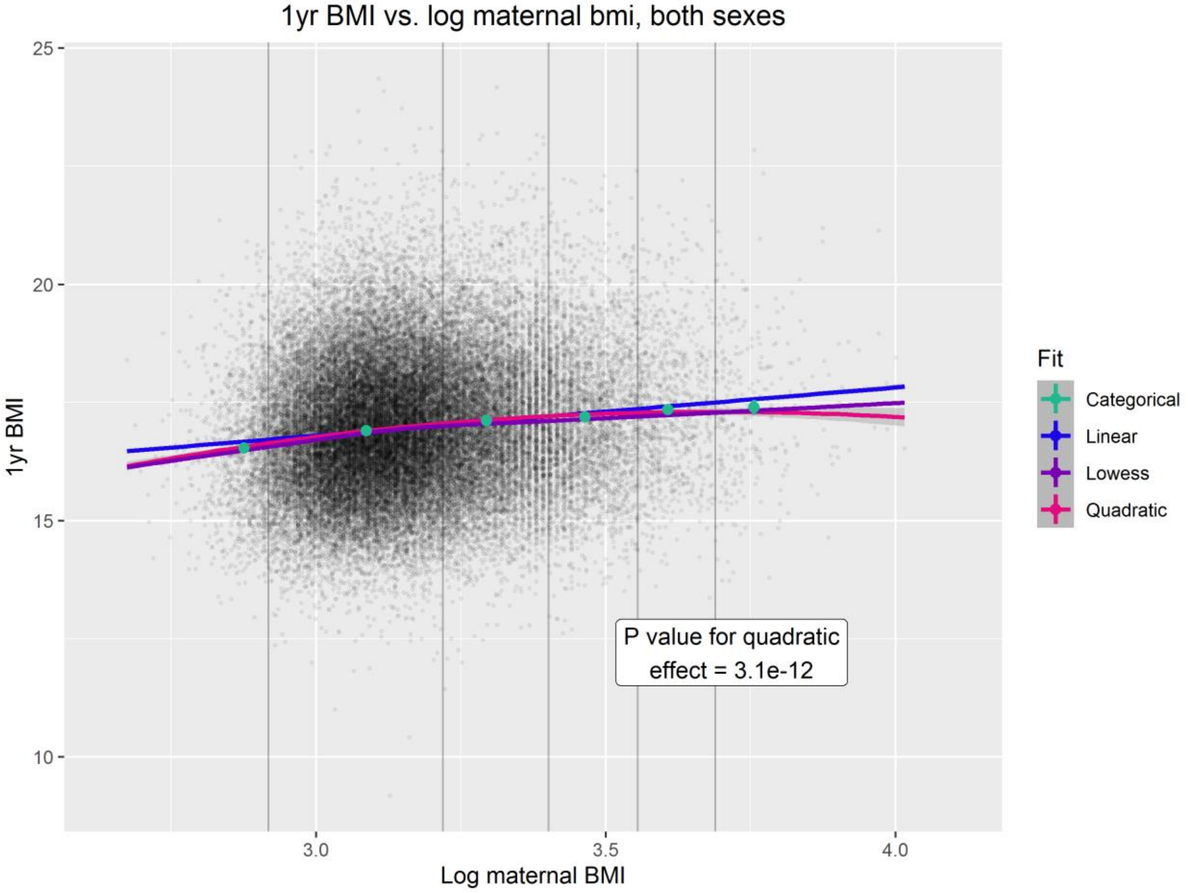
1yr BMI vs. log maternal bmi, both sexes
1yr BMI
Log maternal BMI
25
20
15
10
3.0
3.5
4.0
Fit
Categorical
Linear
Lowess
Quadratic
P value for quadratic effect = 3.1e-12

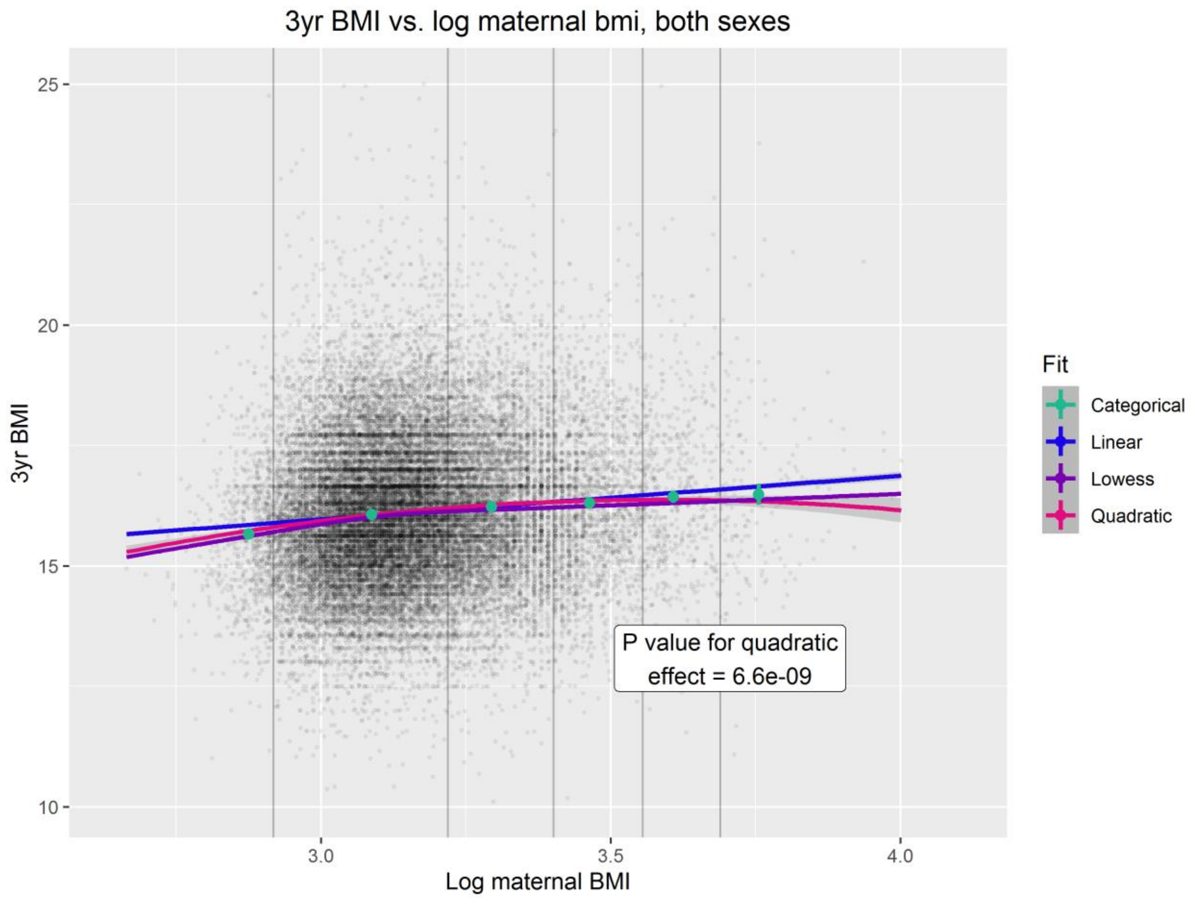
3yr BMI vs. log maternal bmi, both sexes
3yr BMI
Log maternal BMI
25
20
15
10
3.0
3.5
4.0
Fit
Categorical
Linear
Lowess
Quadratic
P value for quadratic
effect = 6.6e-09

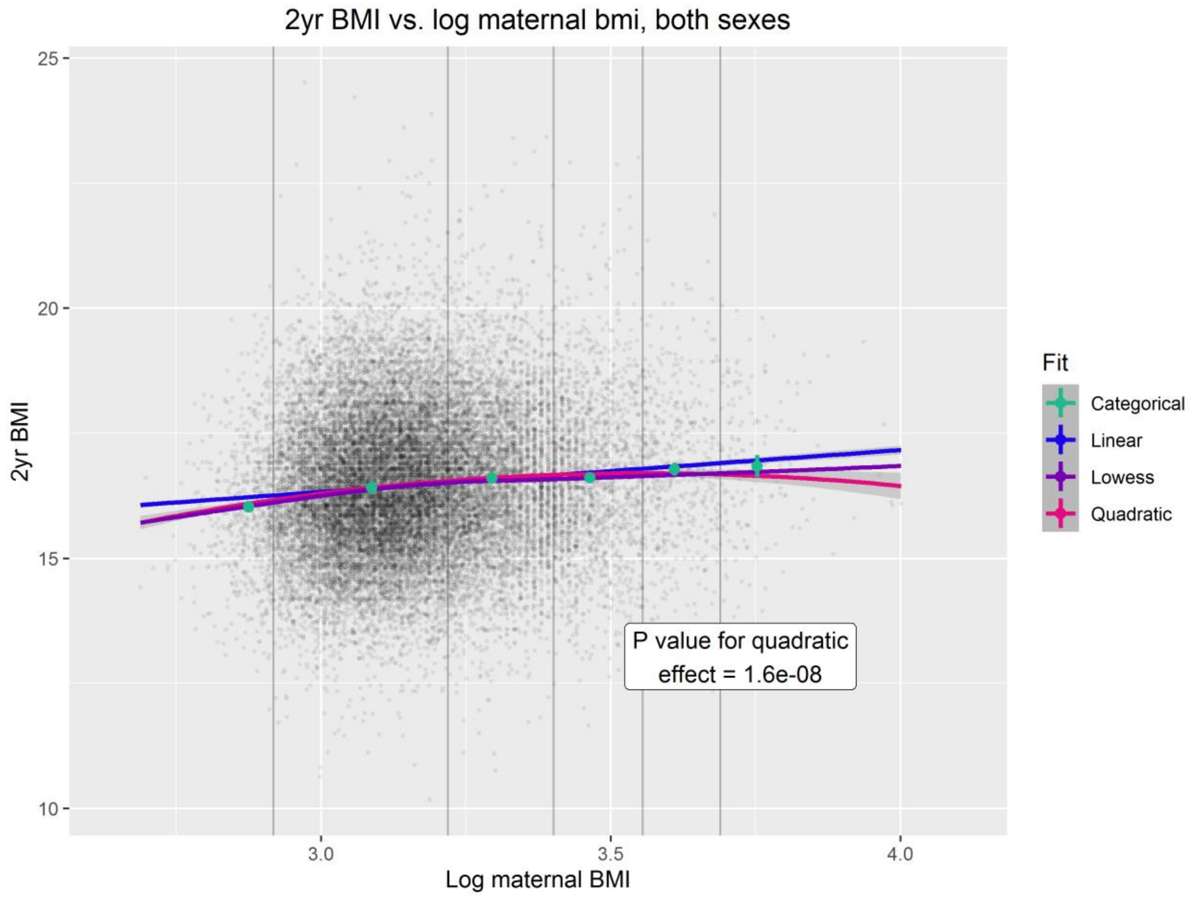
2yr BMI vs. log maternal bmi, both sexes
2yr BMI
Log maternal BMI
25
20
15
10
3.0
3.5
4.0
Fit
Categorical
Linear
Lowess
Quadratic
P value for quadratic
effect = 1.6e-08

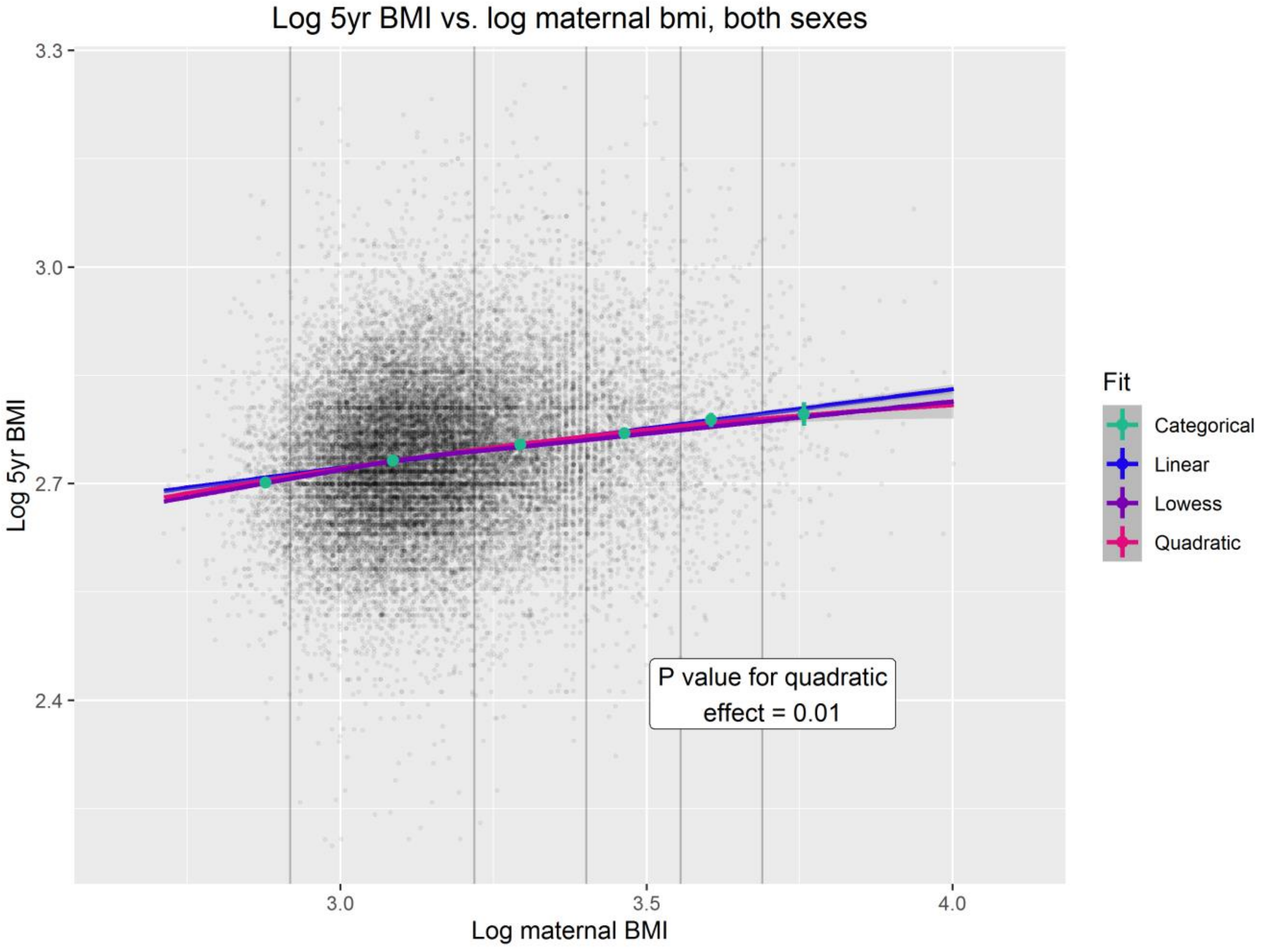
Log 5yr BMI vs. log maternal bmi, both sexes
3.3
3.0
2.7
2.4
Log 5yr BMI
3.0
3.5
4.0
Log maternal BMI
Fit
Categorical
Linear
Lowess
Quadratic
P value for quadratic effect = 0.01

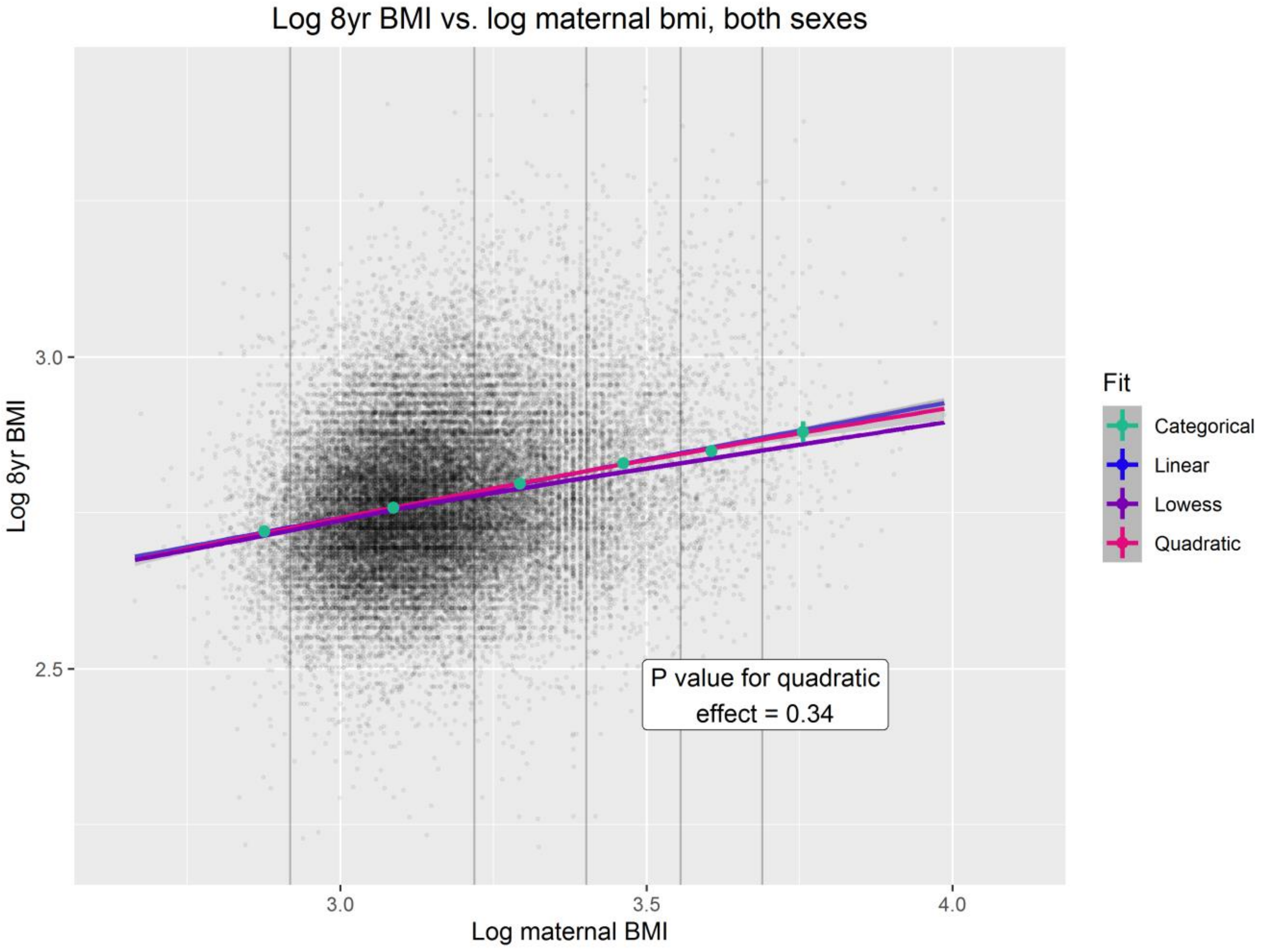
Log 8yr BMI vs. log maternal bmi, both sexes
3.0
2.5
Log 8yr BMI
3.0
3.5
4.0
Log maternal BMI
Fit
Categorical
Linear
Lowess
Quadratic
P value for quadratic effect = 0.34

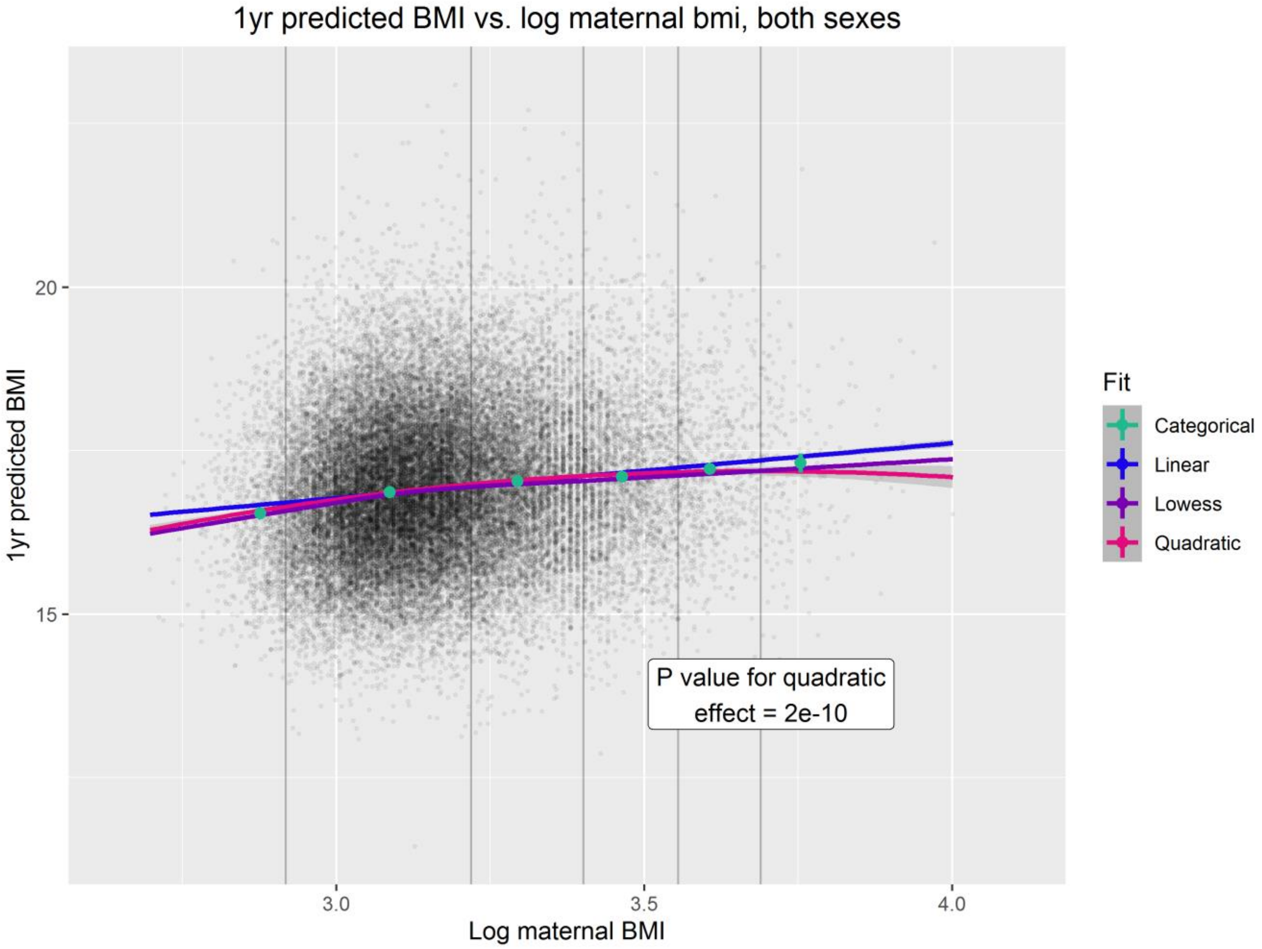
1yr predicted BMI vs. log maternal bmi, both sexes
1yr predicted BMI
20
15
3.0
3.5
4.0
Log maternal BMI
P value for quadratic
effect = 2e-10
Fit
Categorical
Linear
Lowess
Quadratic

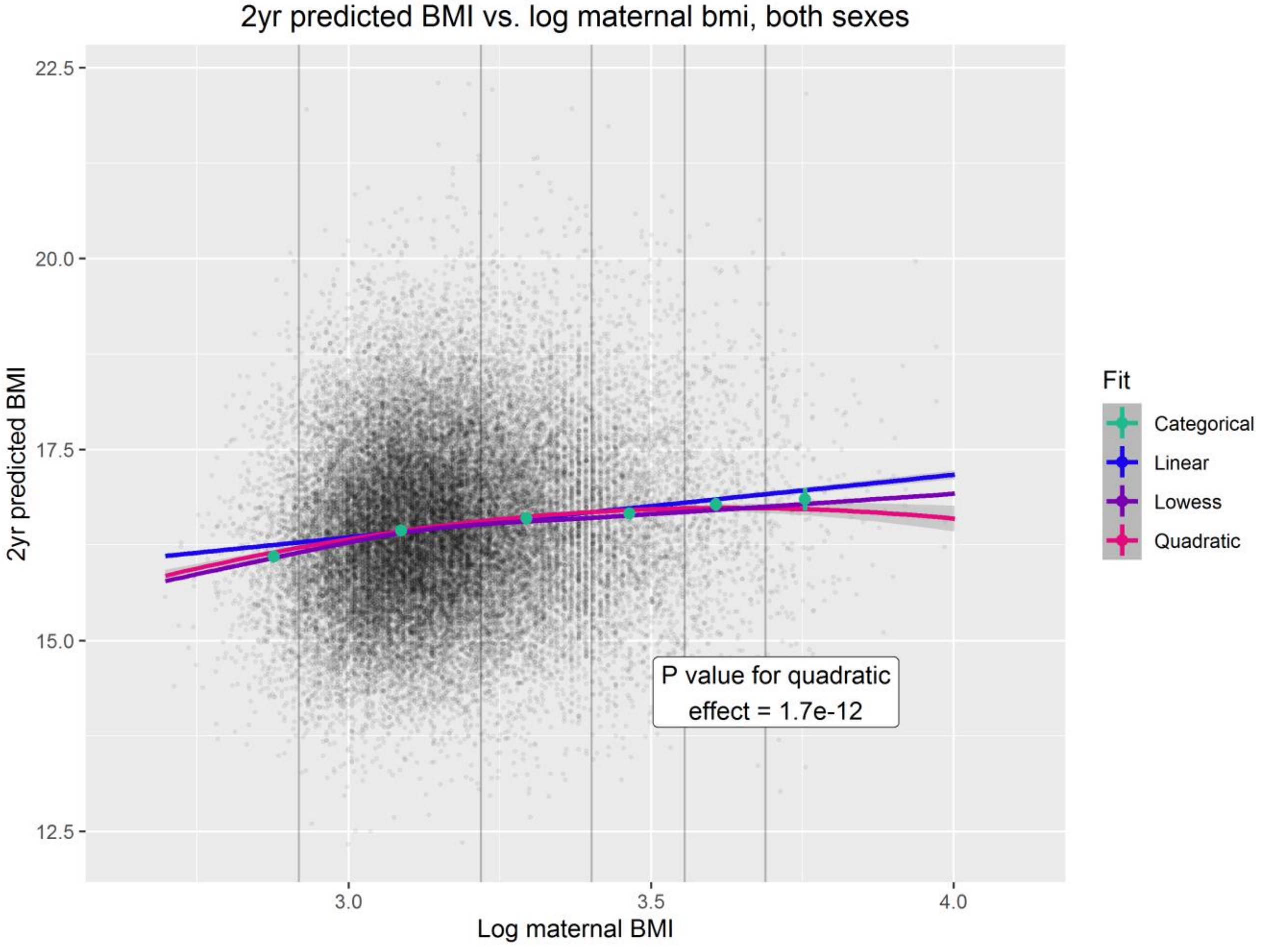
2yr predicted BMI vs. log maternal bmi, both sexes
2yr predicted BMI
22.5
20.0
17.5
15.0
12.5
3.0
3.5
4.0
Log maternal BMI
P value for quadratic
effect = 1.7e-12
Fit
Categorical
Linear
Lowess
Quadratic

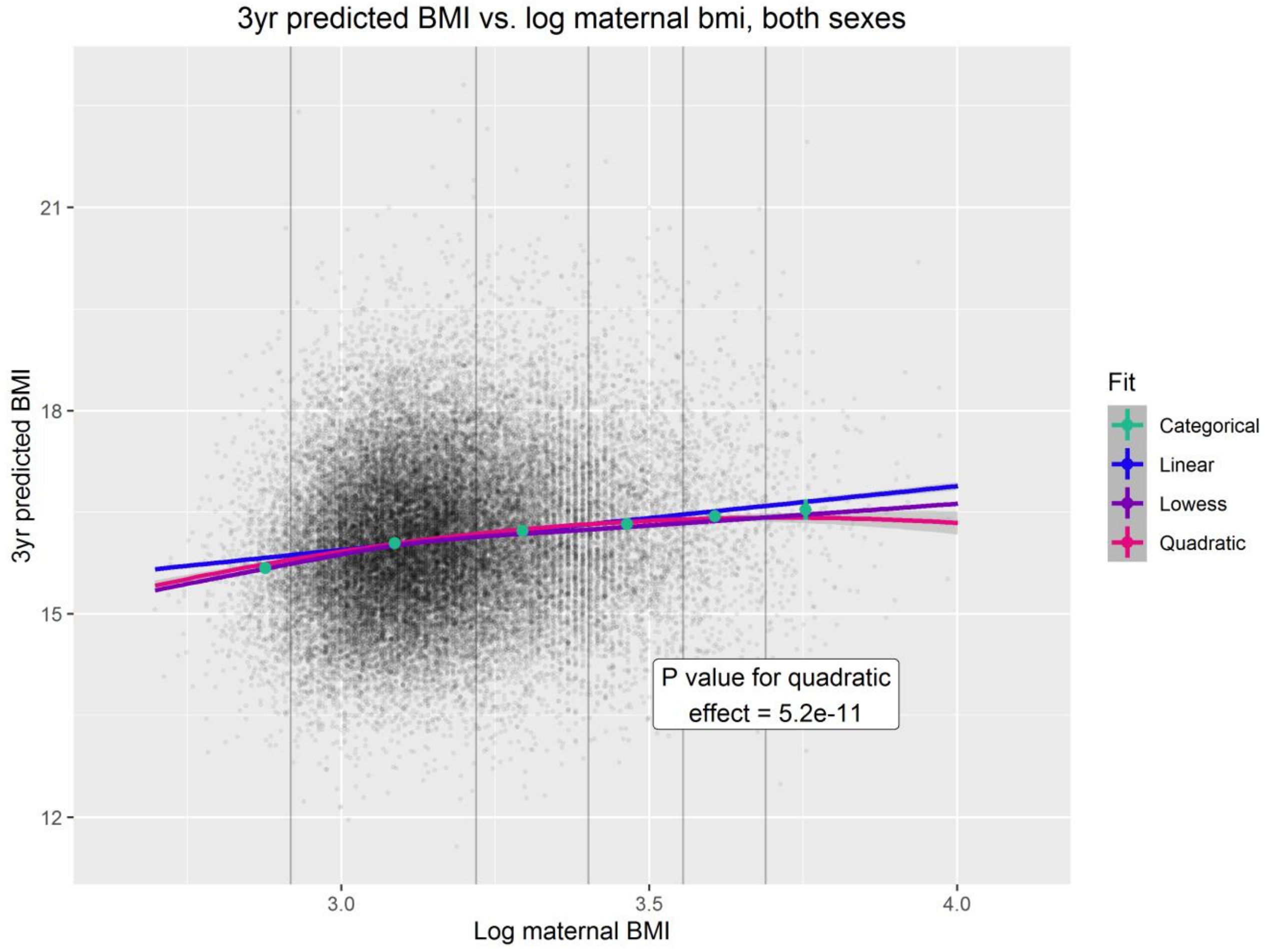
3yr predicted BMI vs. log maternal bmi, both sexes
3yr predicted BMI
21
18
15
12
3.0
3.5
4.0
Log maternal BMI
Fit
Categorical
Linear
Lowess
Quadratic
P value for quadratic effect = 5.2e-11

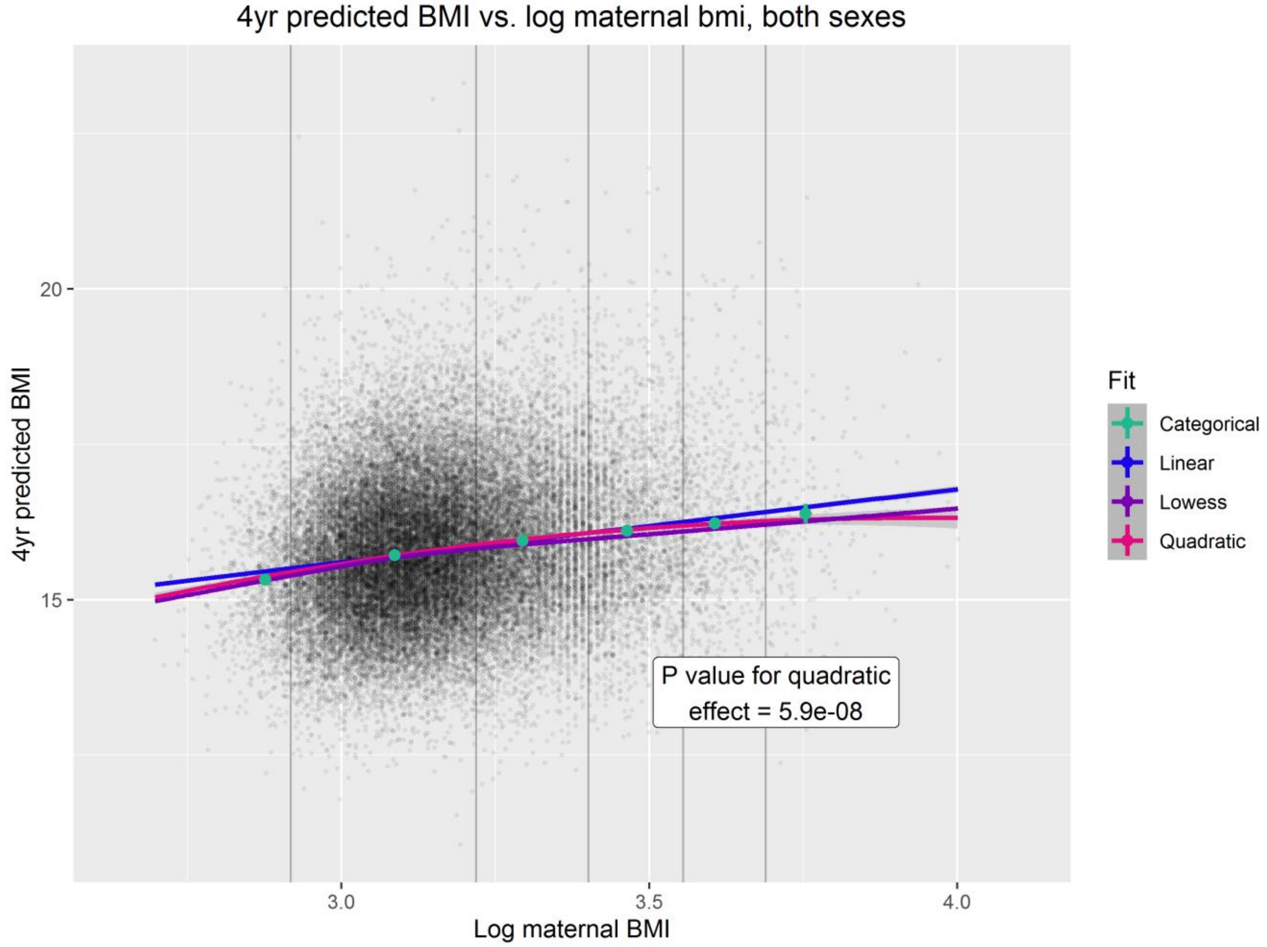
4yr predicted BMI vs. log maternal bmi, both sexes
4yr predicted BMI
20
15
3.0
3.5
4.0
Log maternal BMI
Fit
Categorical
Linear
Lowess
Quadratic
P value for quadratic effect = 5.9e-08

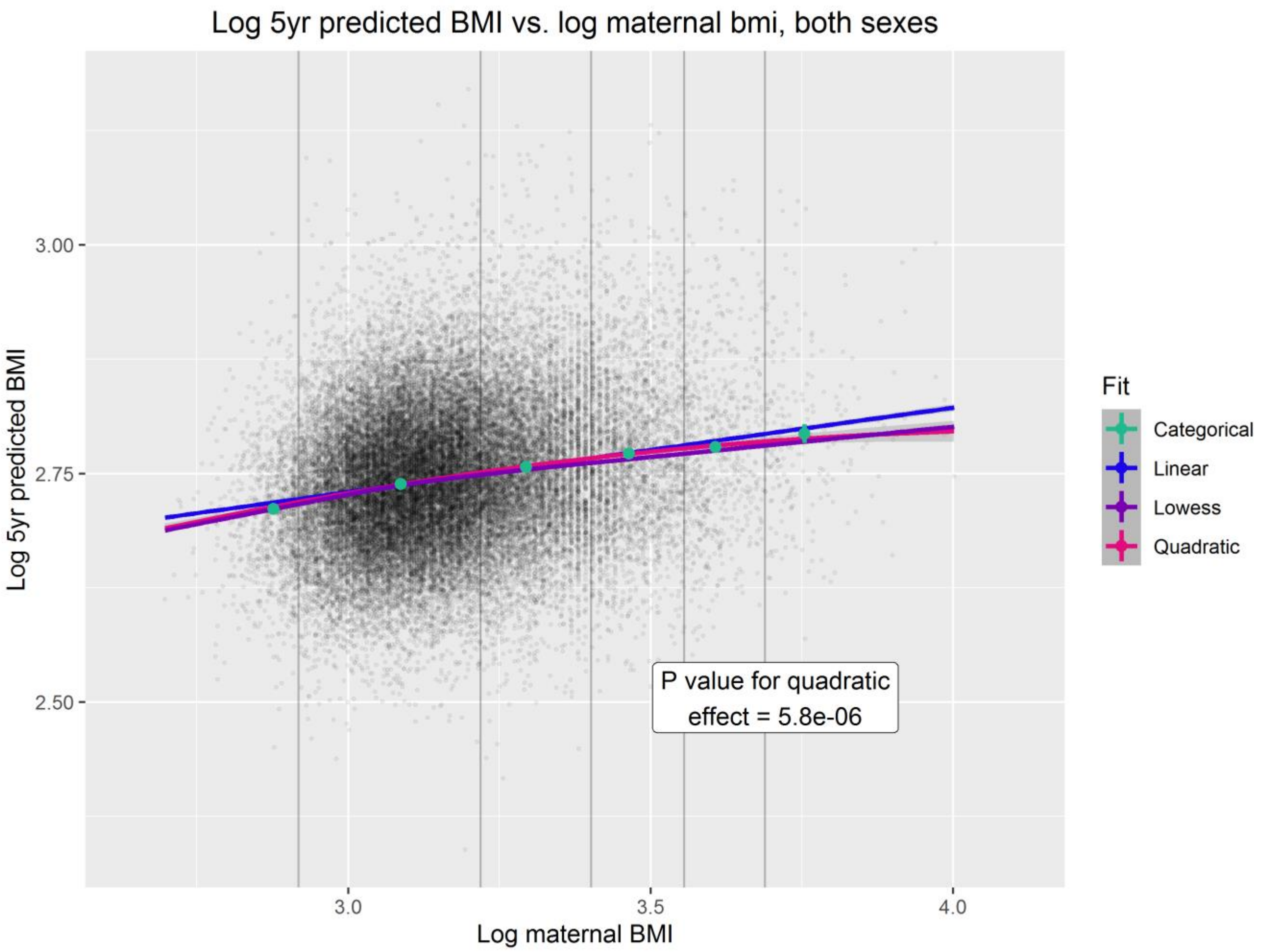
Log 5yr predicted BMI vs. log maternal bmi, both sexes
Log 5yr predicted BMI
3.00
2.75
2.50
3.0
3.5
4.0
Log maternal BMI
Fit
Categorical
Linear
Lowess
Quadratic
P value for quadratic effect = 5.8e-06

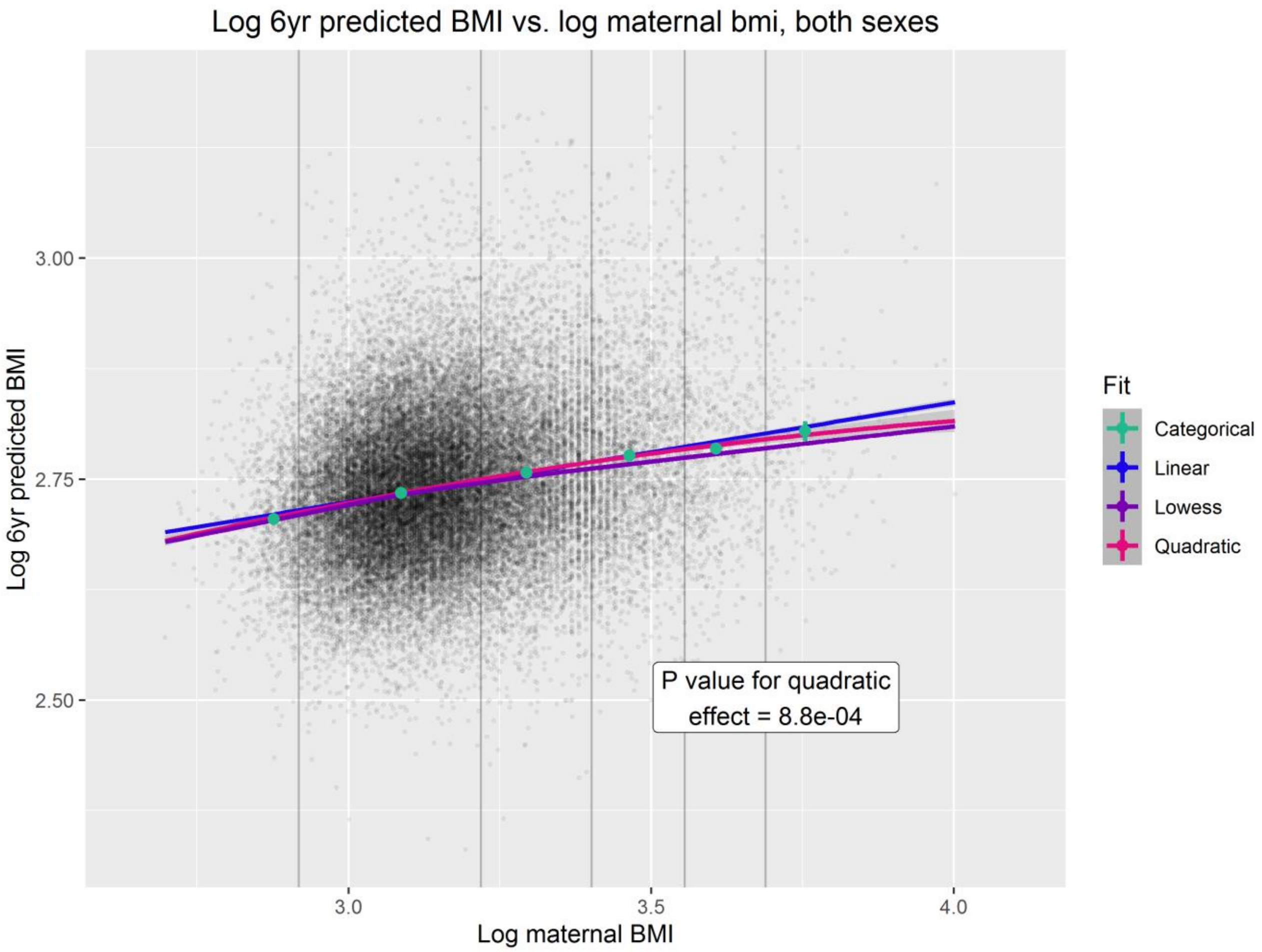
Log 6yr predicted BMI vs. log maternal bmi, both sexes
Log 6yr predicted BMI
3.00
2.75
2.50
3.0
3.5
4.0
Log maternal BMI
Fit
Categorical
Linear
Lowess
Quadratic
P value for quadratic effect = 8.8e-04

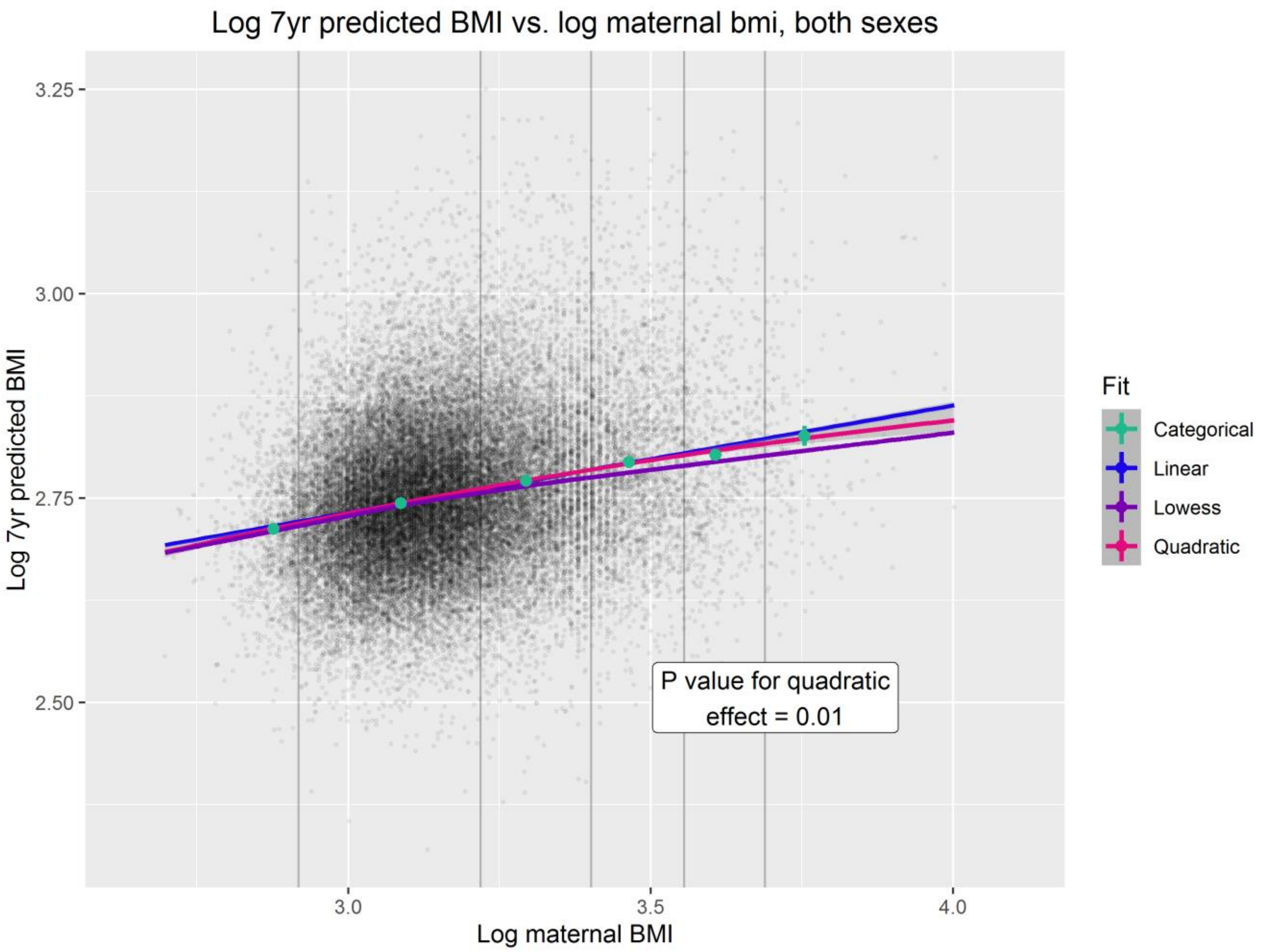
Log 7yr predicted BMI vs. log maternal bmi, both sexes
Log 7yr predicted BMI
3.25
3.00
2.75
2.50
3.0
3.5
4.0
Log maternal BMI
Fit
Categorical
Linear
Lowess
Quadratic
P value for quadratic
effect = 0.01

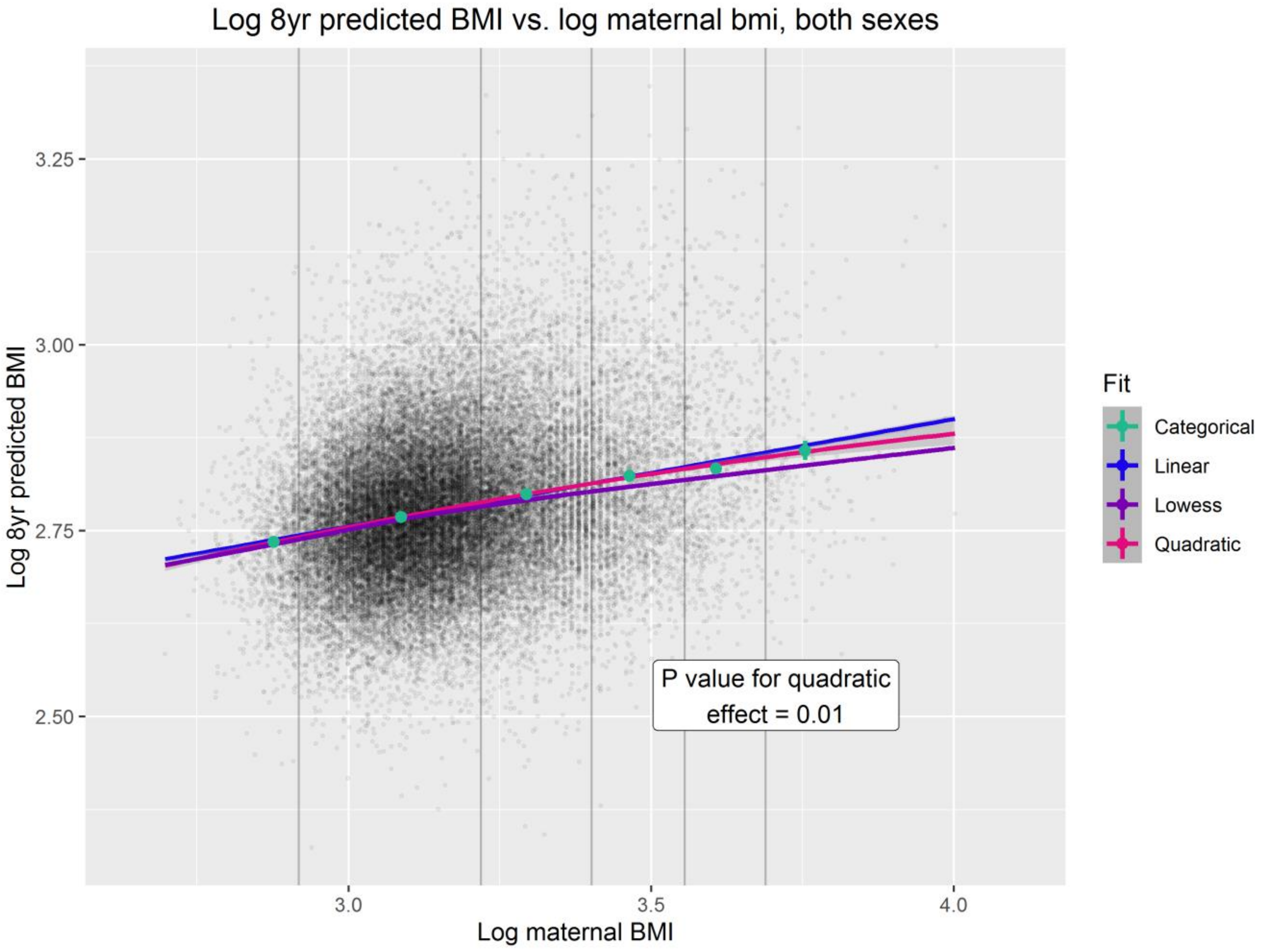
Log 8yr predicted BMI vs. log maternal bmi, both sexes
Log 8yr predicted BMI
3.25
3.00
2.75
2.50
3.0
3.5
4.0
Log maternal BMI
Fit
Categorical
Linear
Lowess
Quadratic
P value for quadratic
effect = 0.01

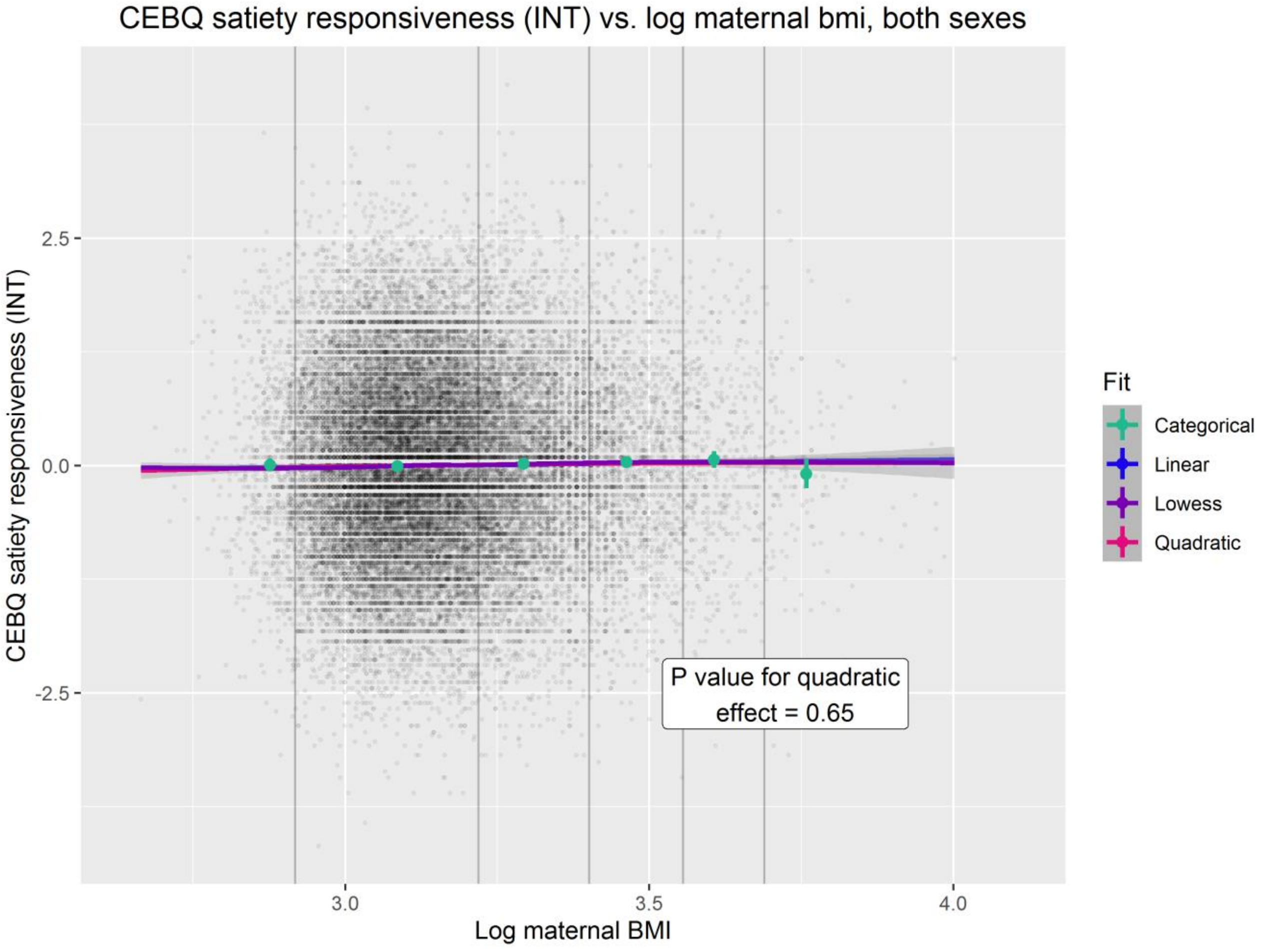
CEBQ satiety responsiveness (INT) vs. log maternal bmi, both sexes
CEBQ satiety responsiveness (INT)
2.5
0.0
-2.5
3.0
3.5
4.0
Log maternal BMI
P value for quadratic effect = 0.65
Fit
Categorical
Linear
Lowess
Quadratic

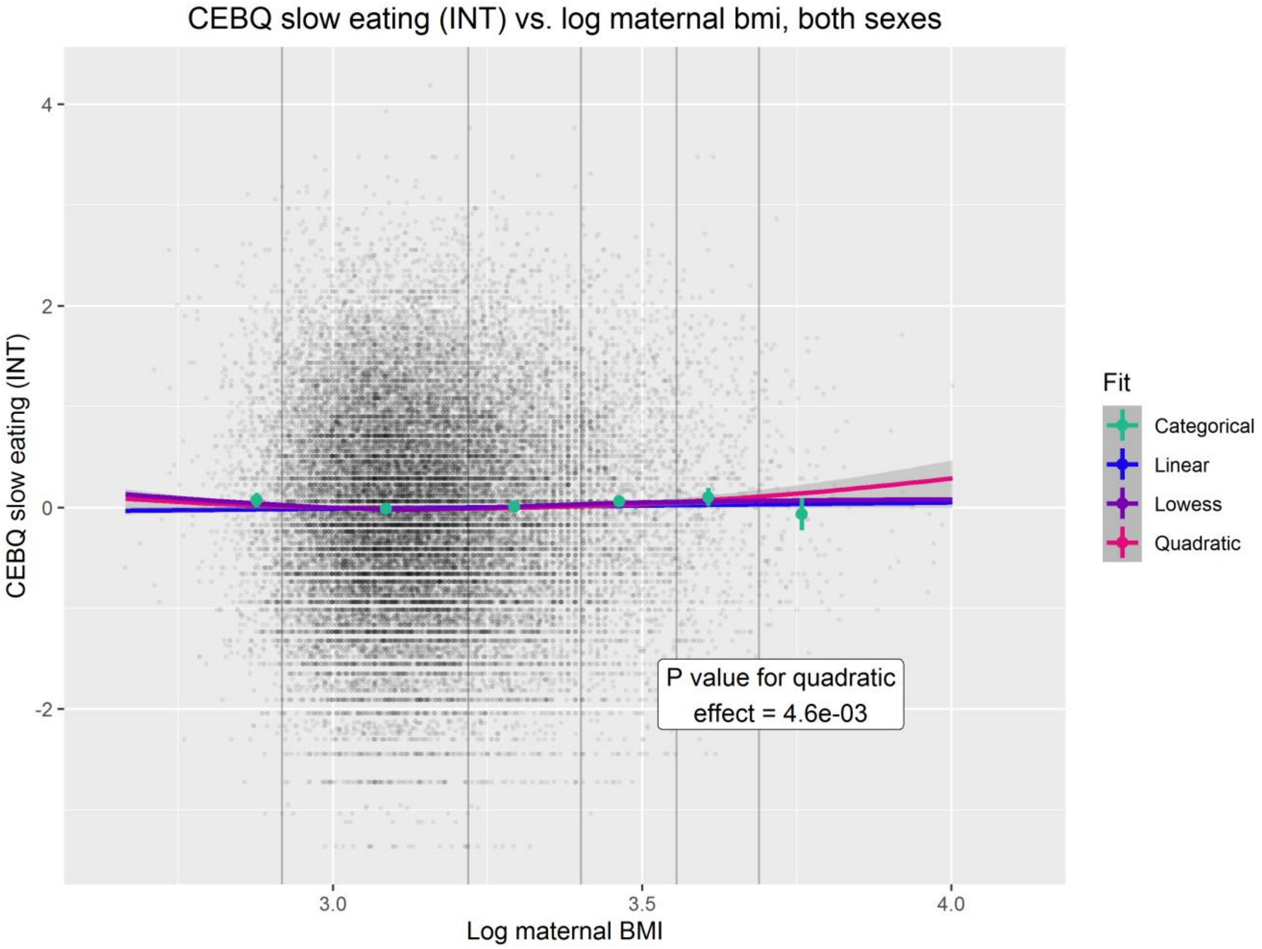
CEBQ slow eating (INT) vs. log maternal bmi, both sexes
CEBQ slow eating (INT)
4
2
0
-2
3.0
3.5
4.0
Log maternal BMI
P value for quadratic effect = 4.6e-03
Fit
Categorical
Linear
Lowess
Quadratic

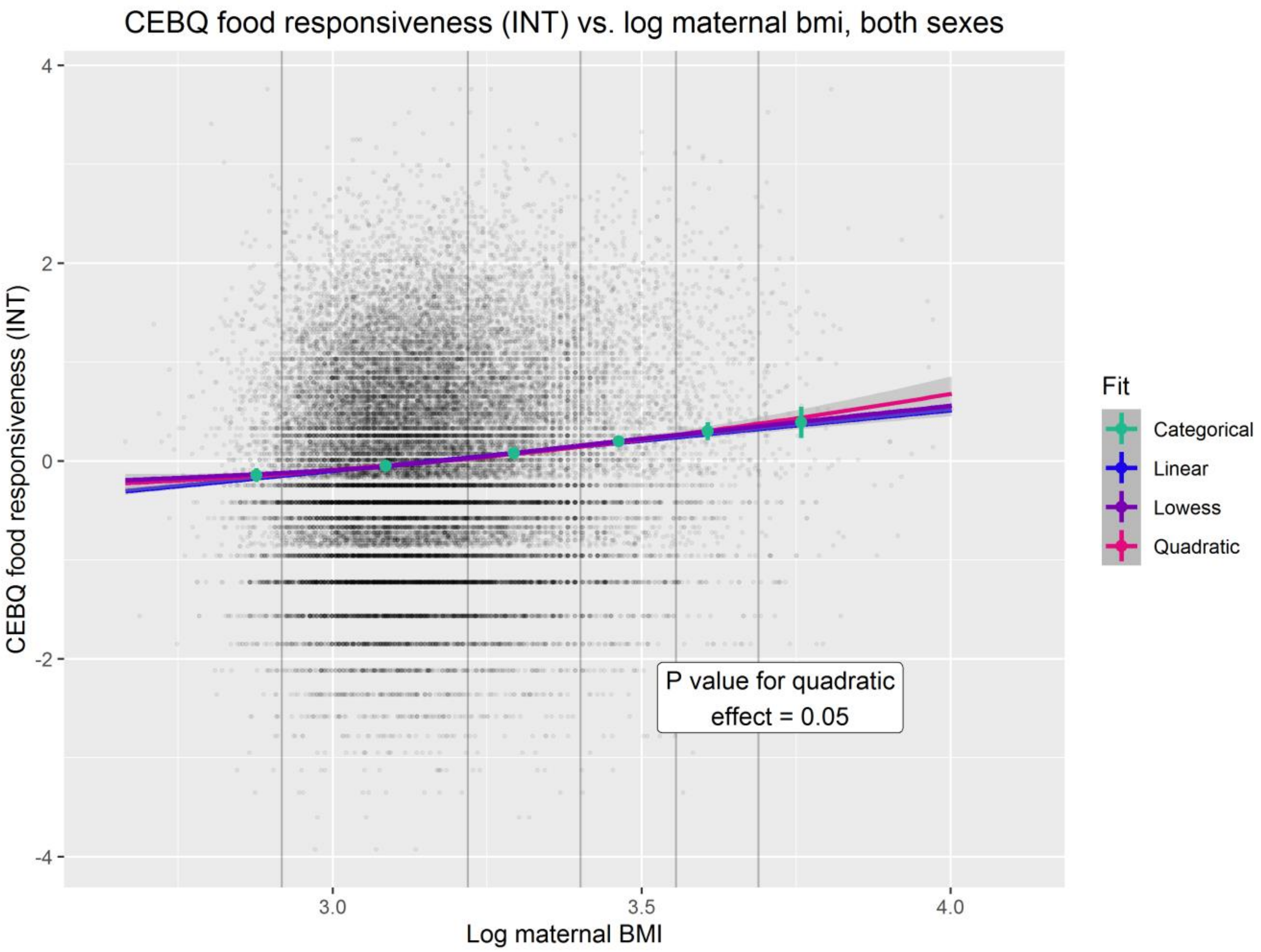
CEBQ food responsiveness (INT) vs. log maternal bmi, both sexes
CEBQ food responsiveness (INT)
4
2
0
-2
-4
3.0
3.5
4.0
Log maternal BMI
Fit
Categorical
Linear
Lowess
Quadratic
P value for quadratic effect = 0.05

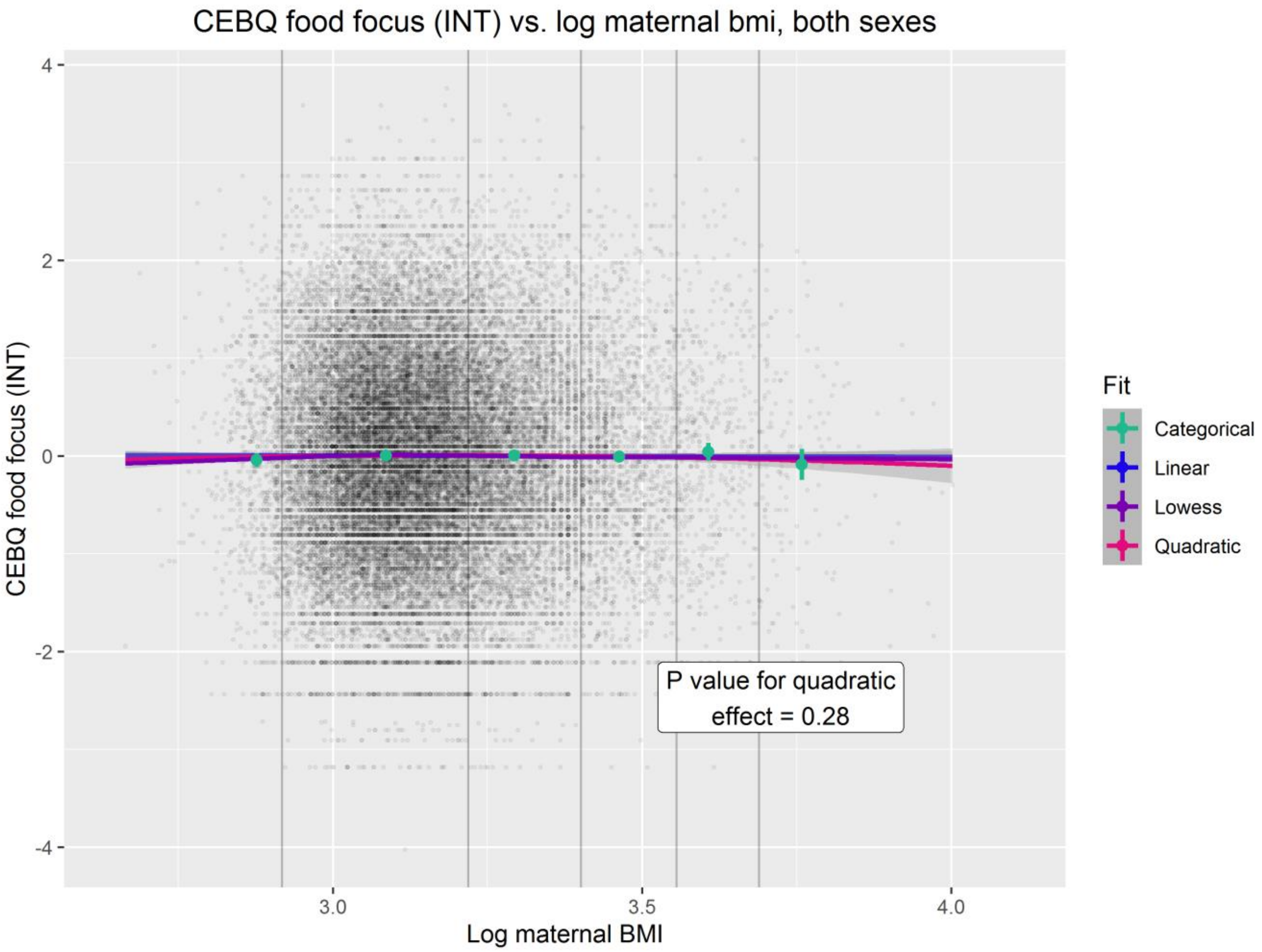
CEBQ food focus (INT) vs. log maternal bmi, both sexes
CEBQ food focus (INT)
4
2
0
-2
-4
3.0
3.5
4.0
Log maternal BMI
Fit
Categorical
Linear
Lowess
Quadratic
P value for quadratic effect = 0.28

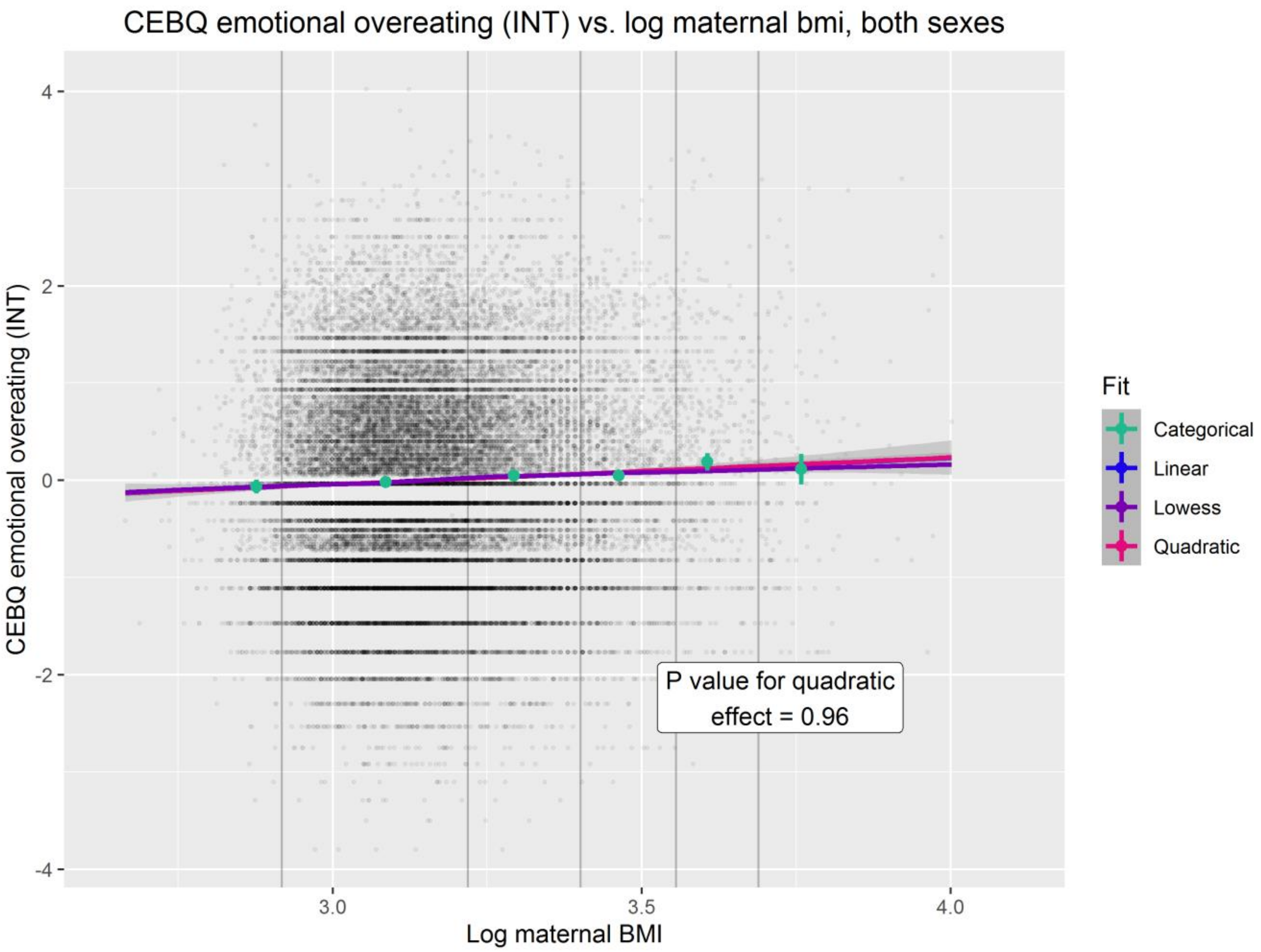
CEBQ emotional overeating (INT) vs. log maternal bmi, both sexes
CEBQ emotional overeating (INT)
Log maternal BMI
4
2
0
-2
-4
3.0
3.5
4.0
Fit
Categorical
Linear
Lowess
Quadratic
P value for quadratic effect = 0.96

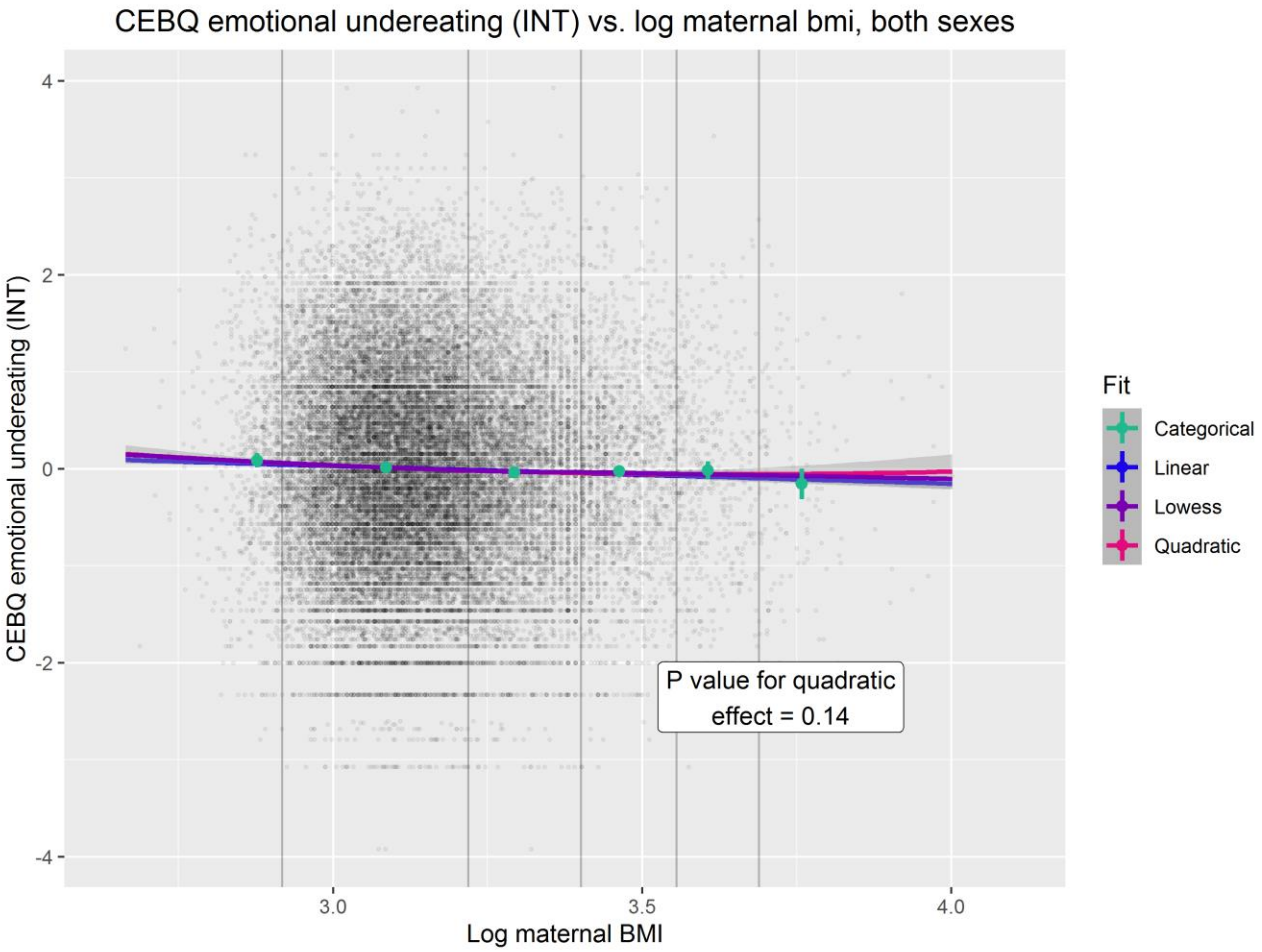
CEBQ emotional undereating (INT) vs. log maternal bmi, both sexes
CEBQ emotional undereating (INT)
Log maternal BMI
4
2
0
-2
-4
3.0
3.5
4.0
Fit
Categorical
Linear
Lowess
Quadratic
P value for quadratic effect = 0.14

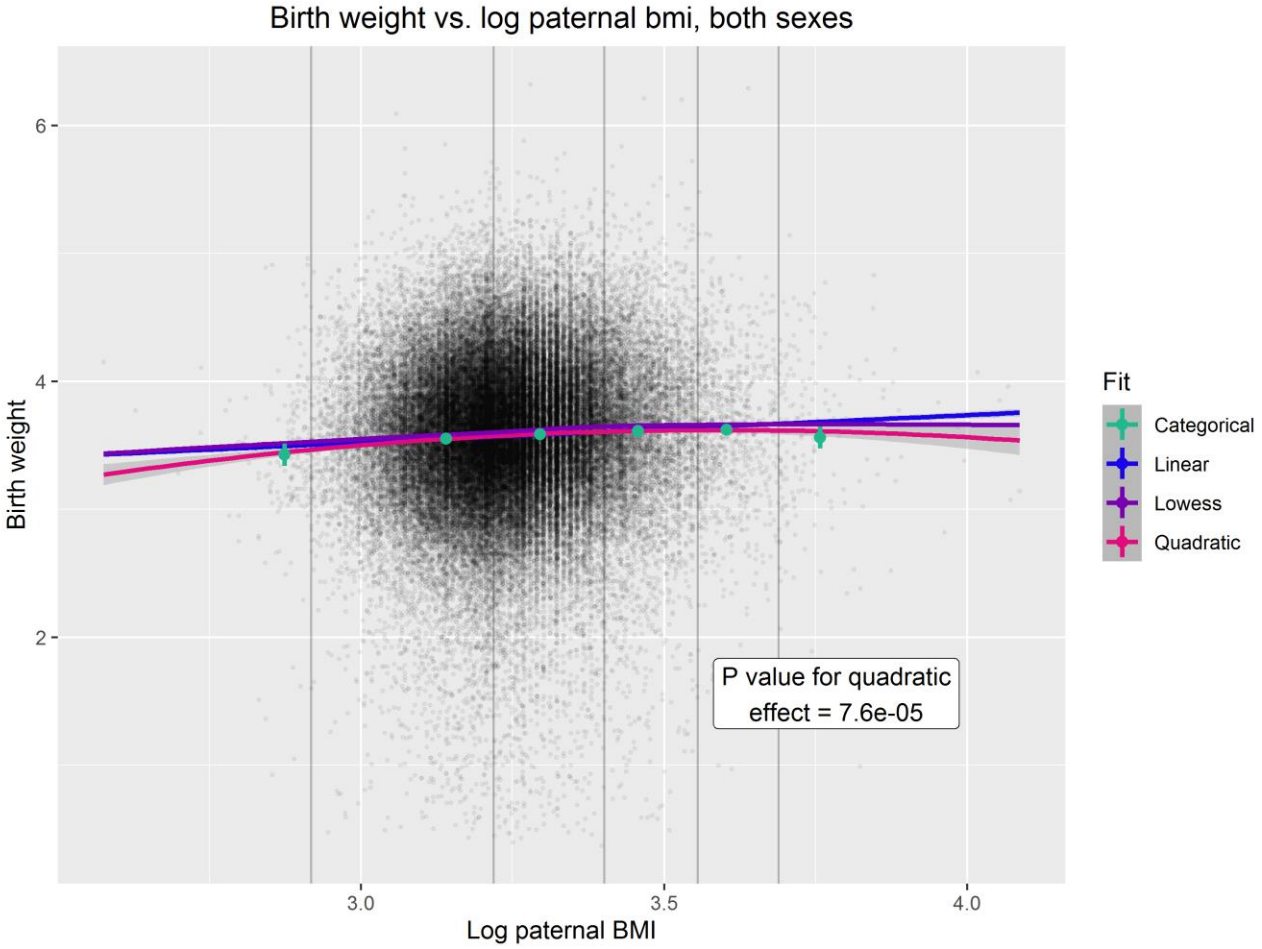
Birth weight vs. log paternal bmi, both sexes
6
4
2
Birth weight
3.0
3.5
4.0
Log paternal BMI
P value for quadratic effect = 7.6e-05
Fit
Categorical
Linear
Lowess
Quadratic

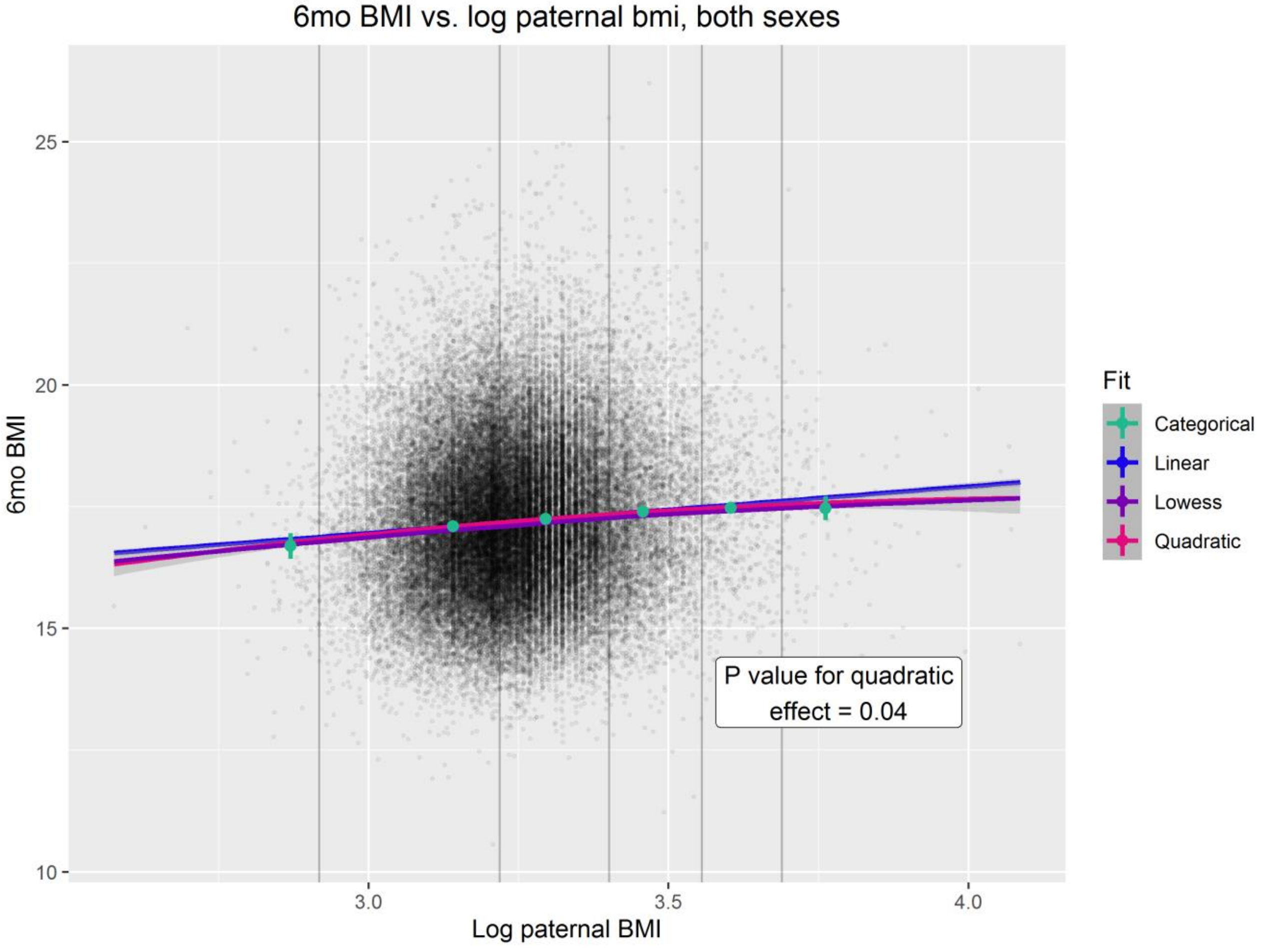
6mo BMI vs. log paternal bmi, both sexes
25
20
15
10
6mo BMI
3.0
3.5
4.0
Log paternal BMI
P value for quadratic effect = 0.04
Fit
Categorical
Linear
Lowess
Quadratic

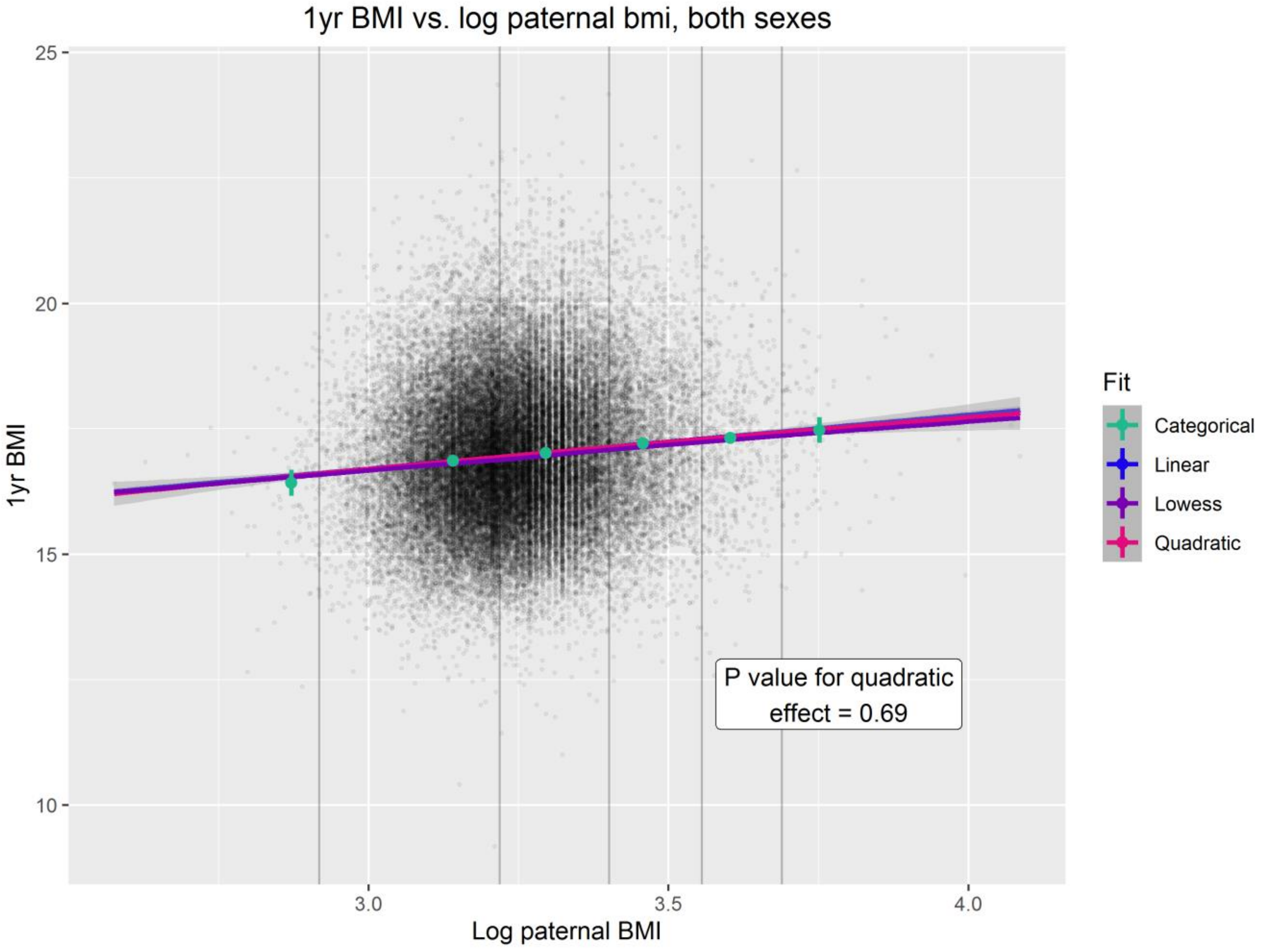
1yr BMI vs. log paternal bmi, both sexes
1yr BMI
Log paternal BMI
25
20
15
10
3.0
3.5
4.0
P value for quadratic effect = 0.69
Fit
Categorical
Linear
Lowess
Quadratic

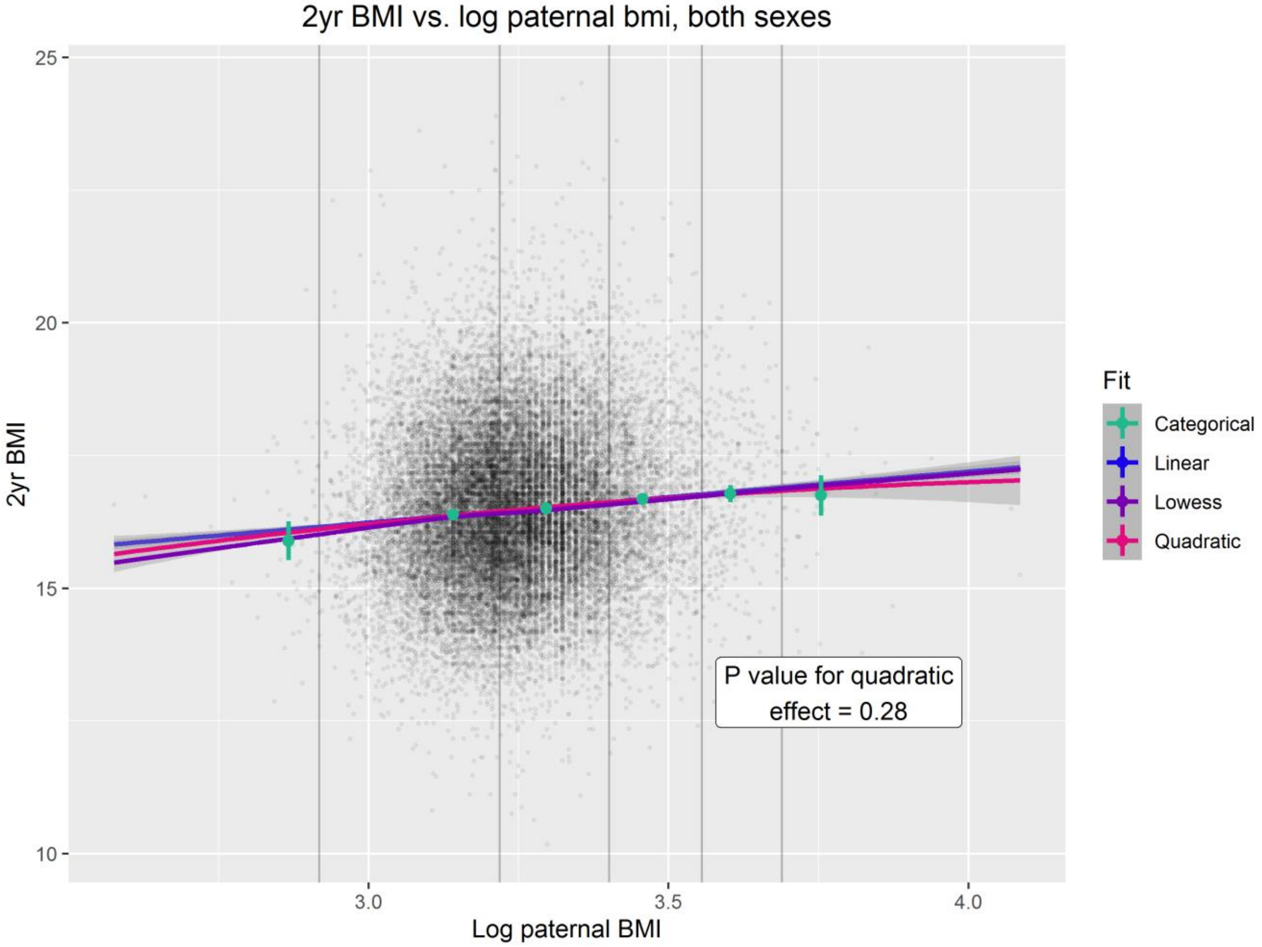
2yr BMI vs. log paternal bmi, both sexes
2yr BMI
Log paternal BMI
25
20
15
10
3.0
3.5
4.0
P value for quadratic effect = 0.28
Fit
Categorical
Linear
Lowess
Quadratic

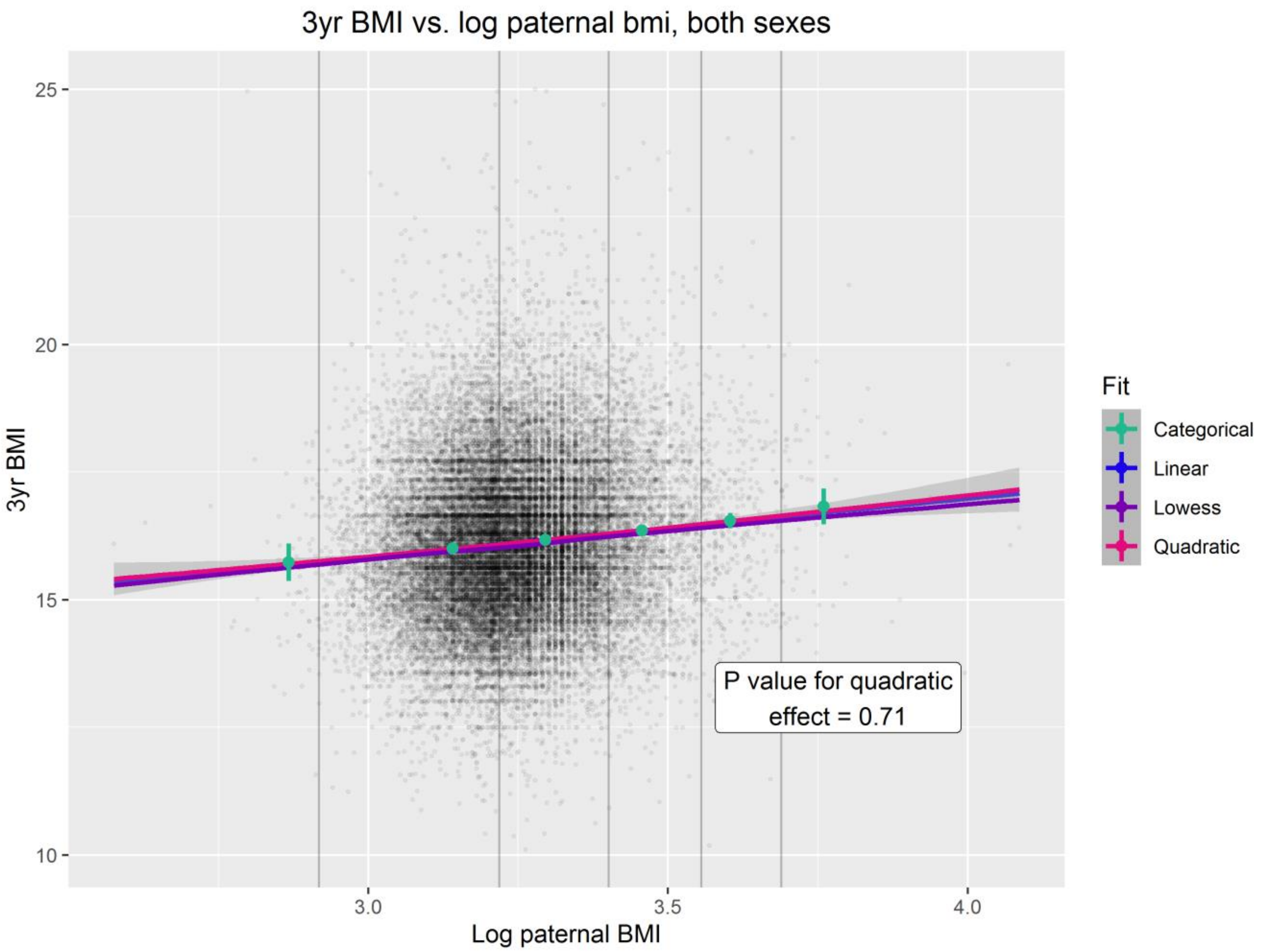
3yr BMI vs. log paternal bmi, both sexes
3yr BMI
Log paternal BMI
25
20
15
10
3.0
3.5
4.0
Fit
Categorical
Linear
Lowess
Quadratic
P value for quadratic effect = 0.71

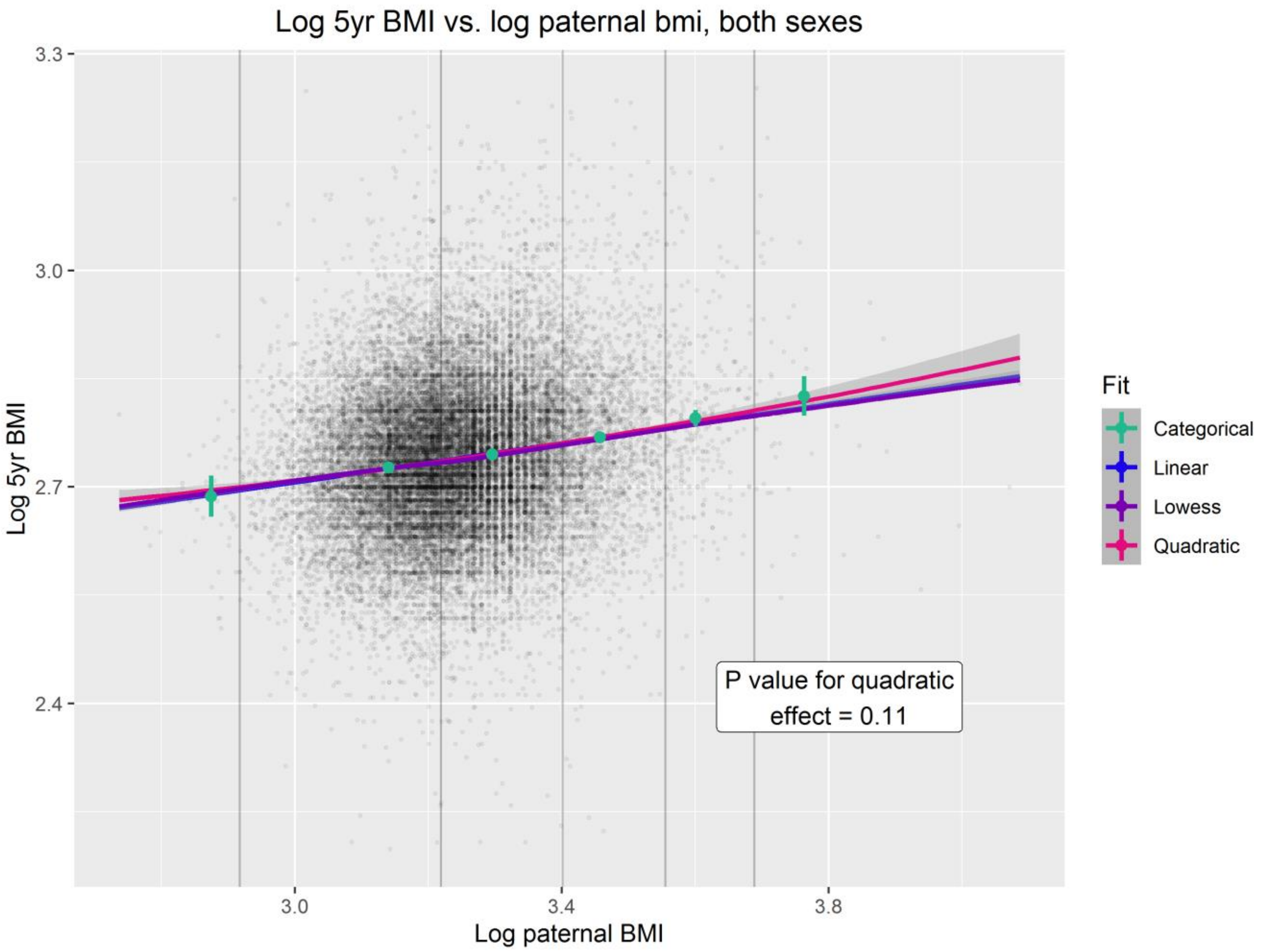
Log 5yr BMI vs. log paternal bmi, both sexes
Log 5yr BMI
Log paternal BMI
3.3
3.0
2.7
2.4
3.0
3.4
3.8
Fit
Categorical
Linear
Lowess
Quadratic
P value for quadratic effect = 0.11

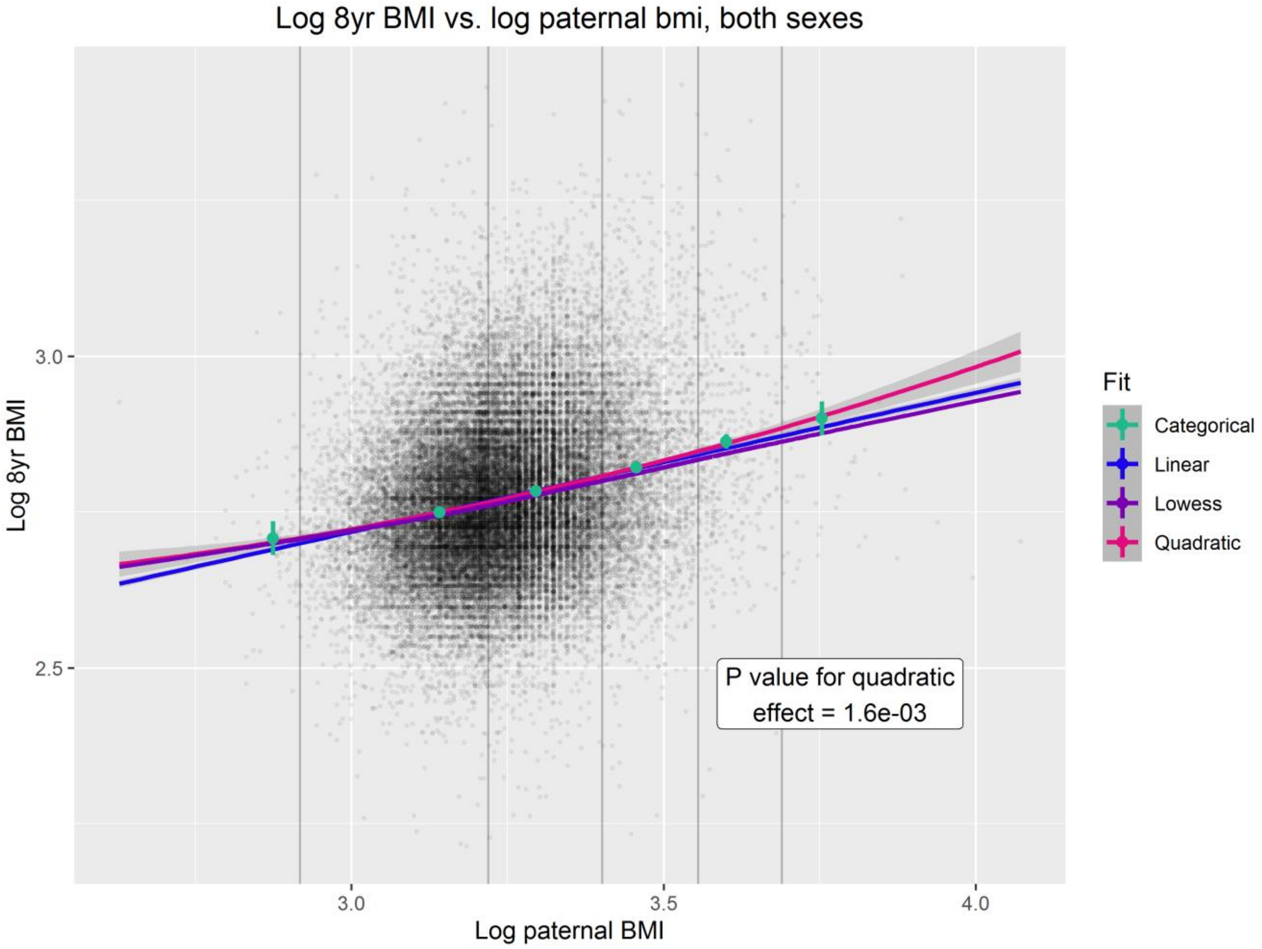
Log 8yr BMI vs. log paternal bmi, both sexes
Log 8yr BMI
3.0
2.5
Log paternal BMI
3.0
3.5
4.0
Fit
Categorical
Linear
Lowess
Quadratic
P value for quadratic effect = 1.6e-03

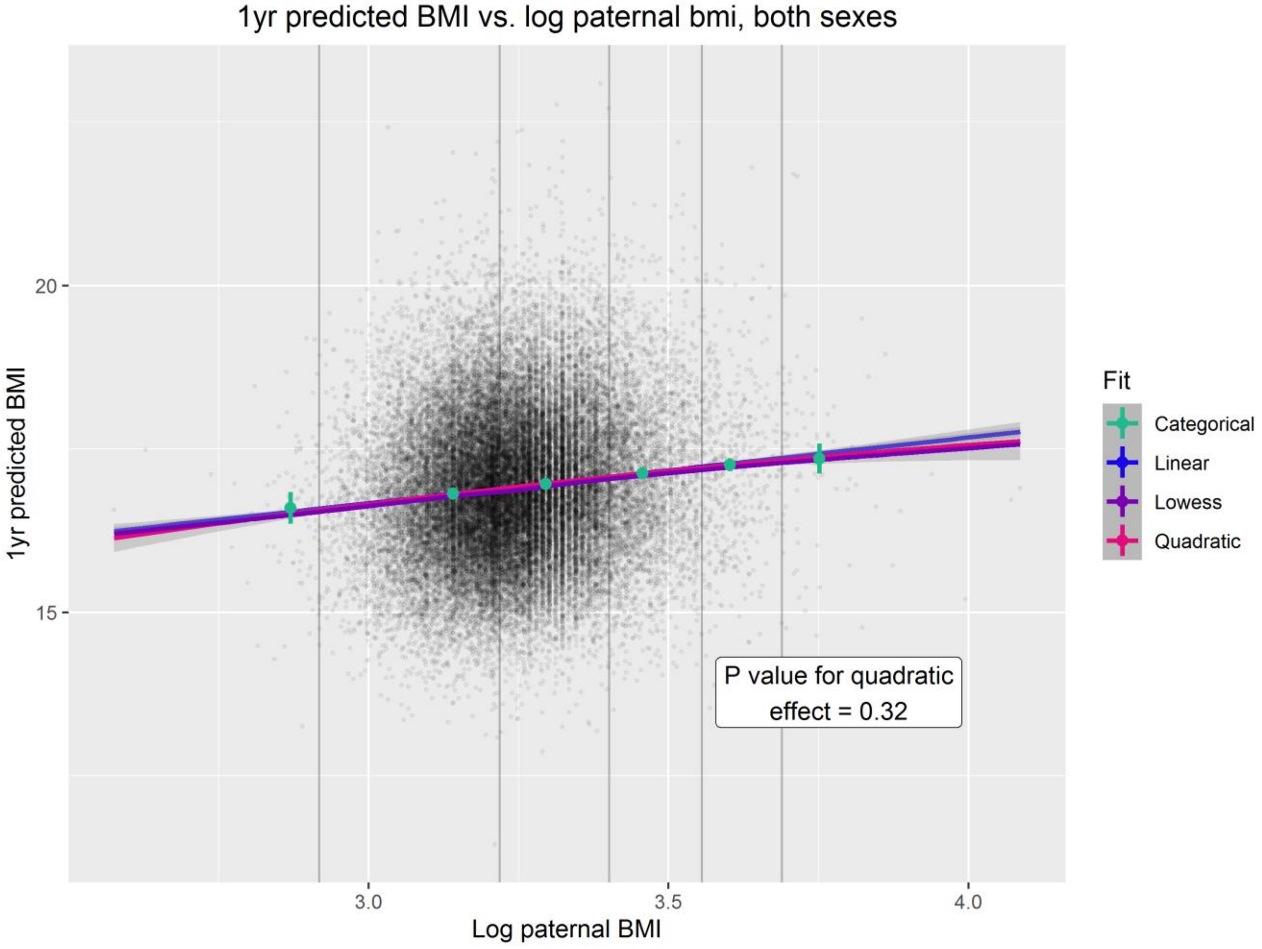
1yr predicted BMI vs. log paternal bmi, both sexes
1yr predicted BMI
20
15
Log paternal BMI
3.0
3.5
4.0
Fit
Categorical
Linear
Lowess
Quadratic
P value for quadratic effect = 0.32

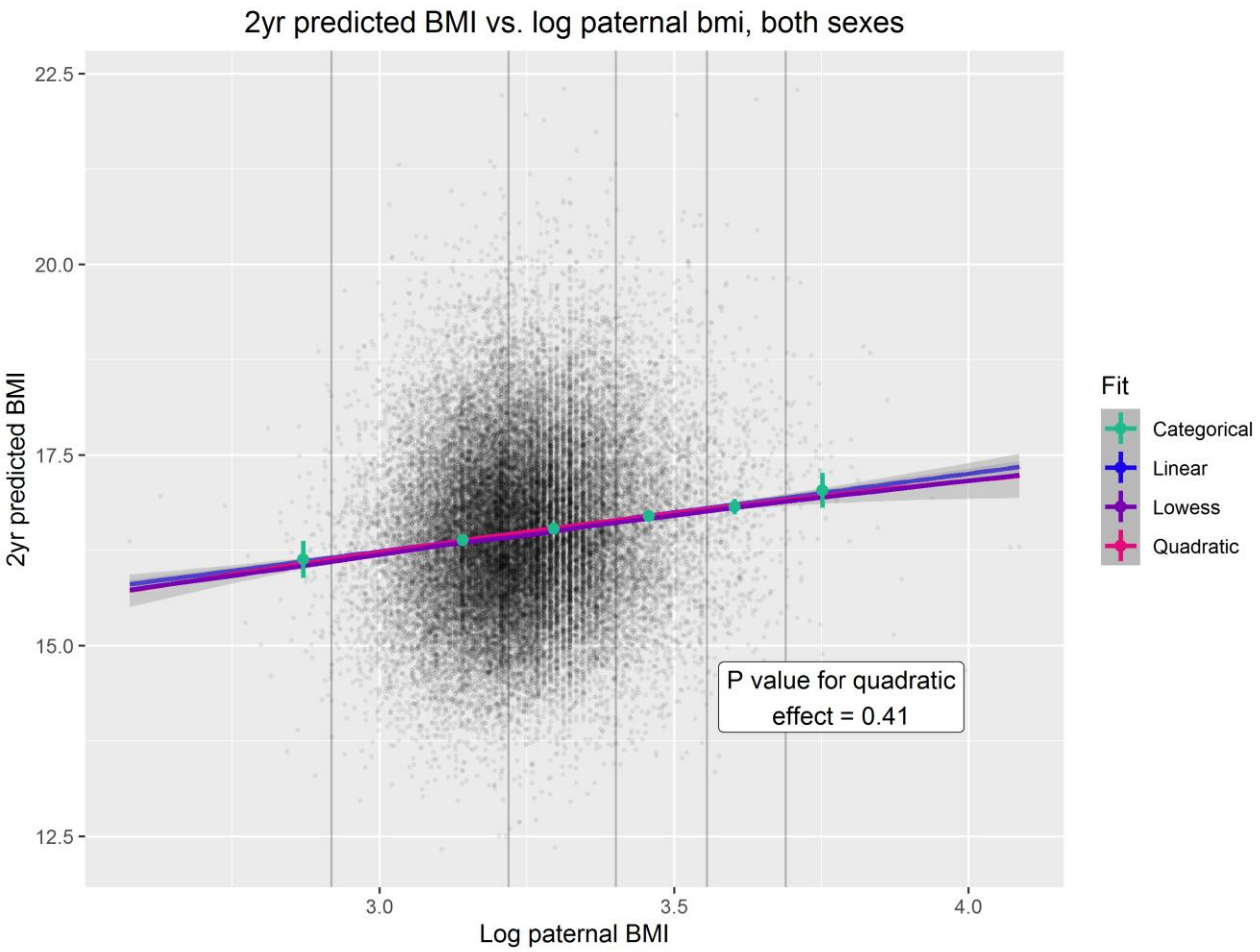
2yr predicted BMI vs. log paternal bmi, both sexes
22.5
20.0
17.5
15.0
12.5
2yr predicted BMI
3.0
3.5
4.0
Log paternal BMI
Fit
Categorical
Linear
Lowess
Quadratic
P value for quadratic effect = 0.41

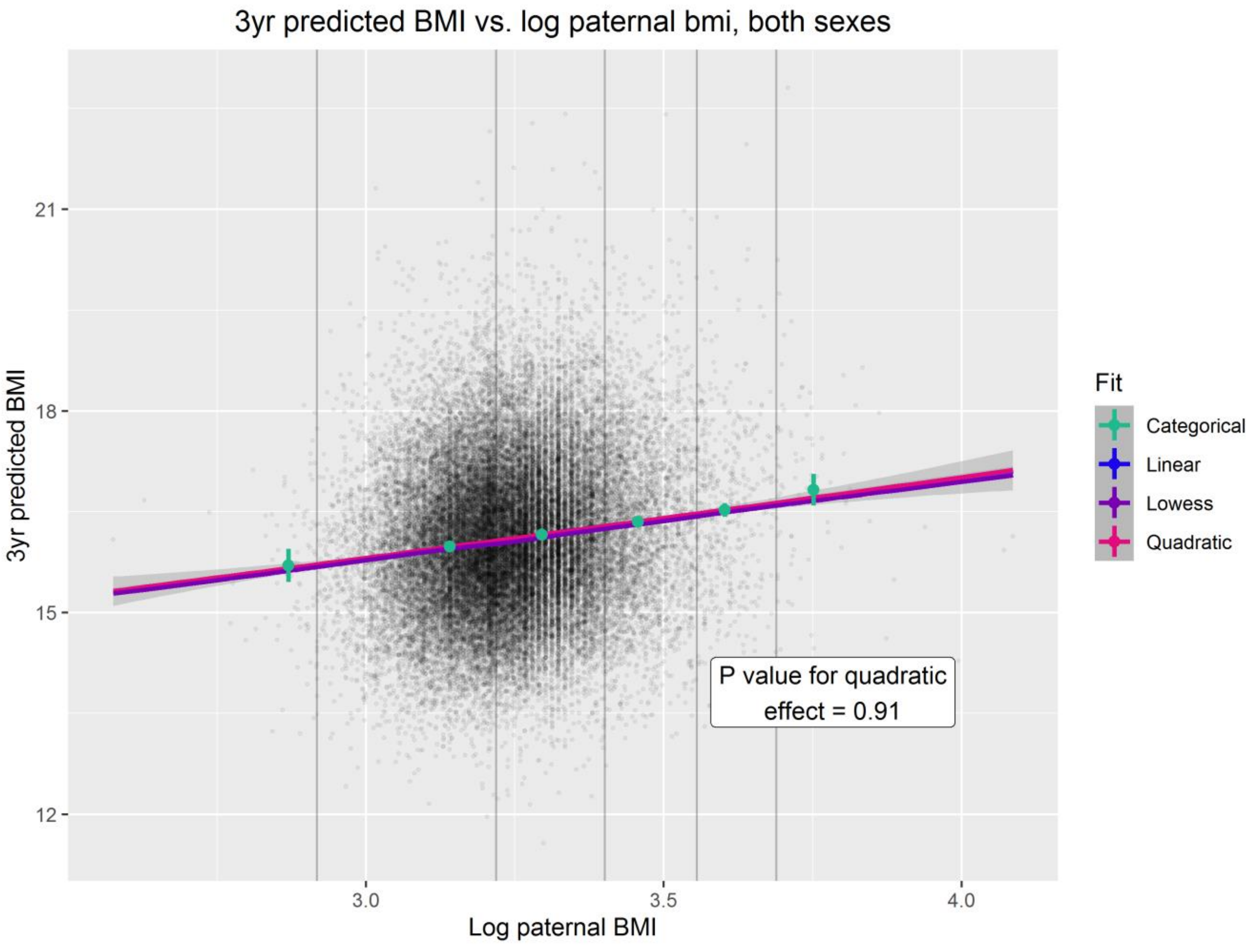
3yr predicted BMI vs. log paternal bmi, both sexes
21
18
15
12
3yr predicted BMI
3.0
3.5
4.0
Log paternal BMI
Fit
Categorical
Linear
Lowess
Quadratic
P value for quadratic effect = 0.91

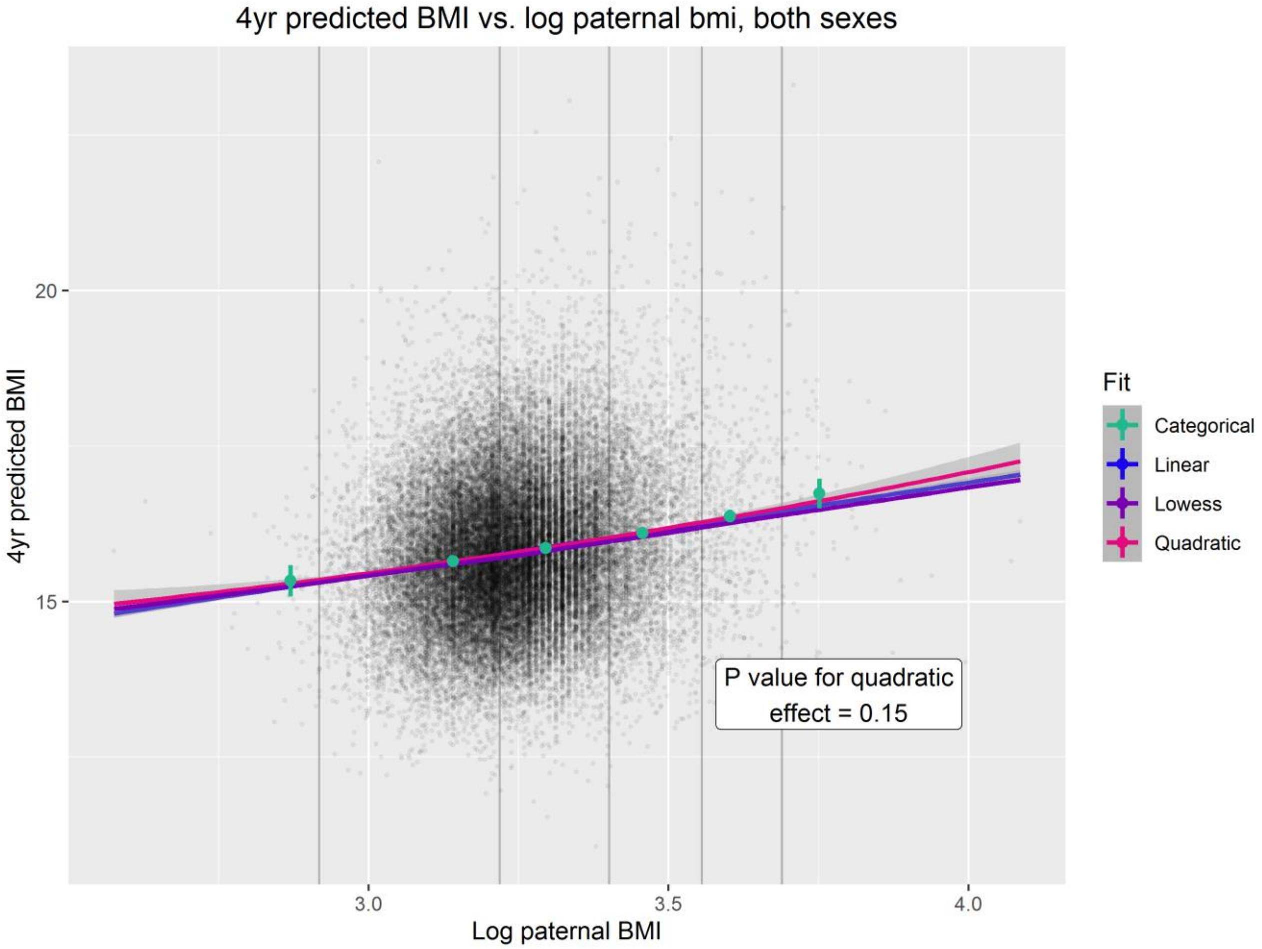
4yr predicted BMI vs. log paternal bmi, both sexes
4yr predicted BMI
20
15
3.0
3.5
4.0
Log paternal BMI
Fit
Categorical
Linear
Lowess
Quadratic
P value for quadratic effect = 0.15

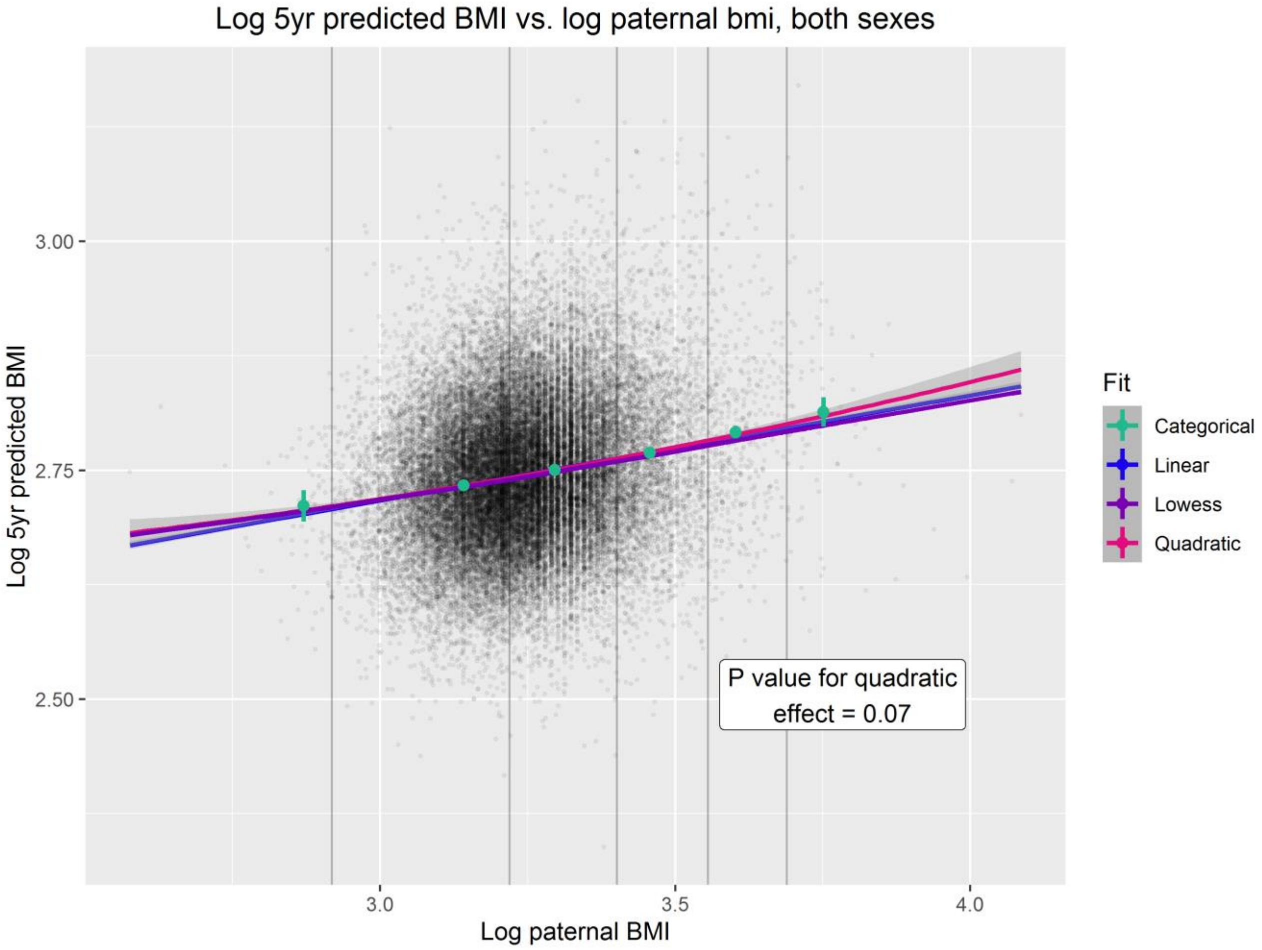
Log 5yr predicted BMI vs. log paternal bmi, both sexes
Log 5yr predicted BMI
3.00
2.75
2.50
3.0
3.5
4.0
Log paternal BMI
Fit
Categorical
Linear
Lowess
Quadratic
P value for quadratic effect = 0.07

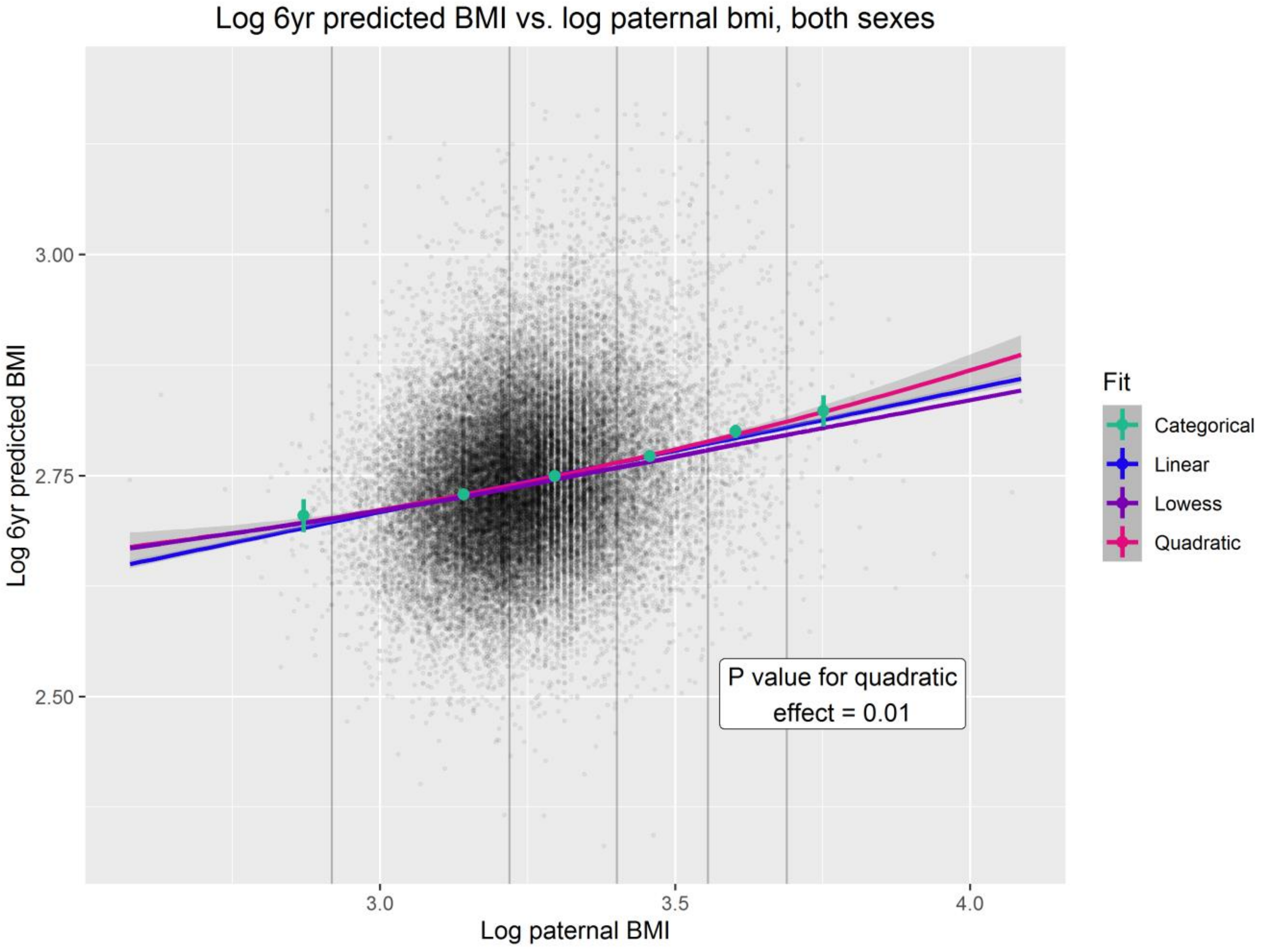
Log 6yr predicted BMI vs. log paternal bmi, both sexes
Log 6yr predicted BMI
3.00
2.75
2.50
Log paternal BMI
3.0
3.5
4.0
Fit
Categorical
Linear
Lowess
Quadratic
P value for quadratic effect = 0.01

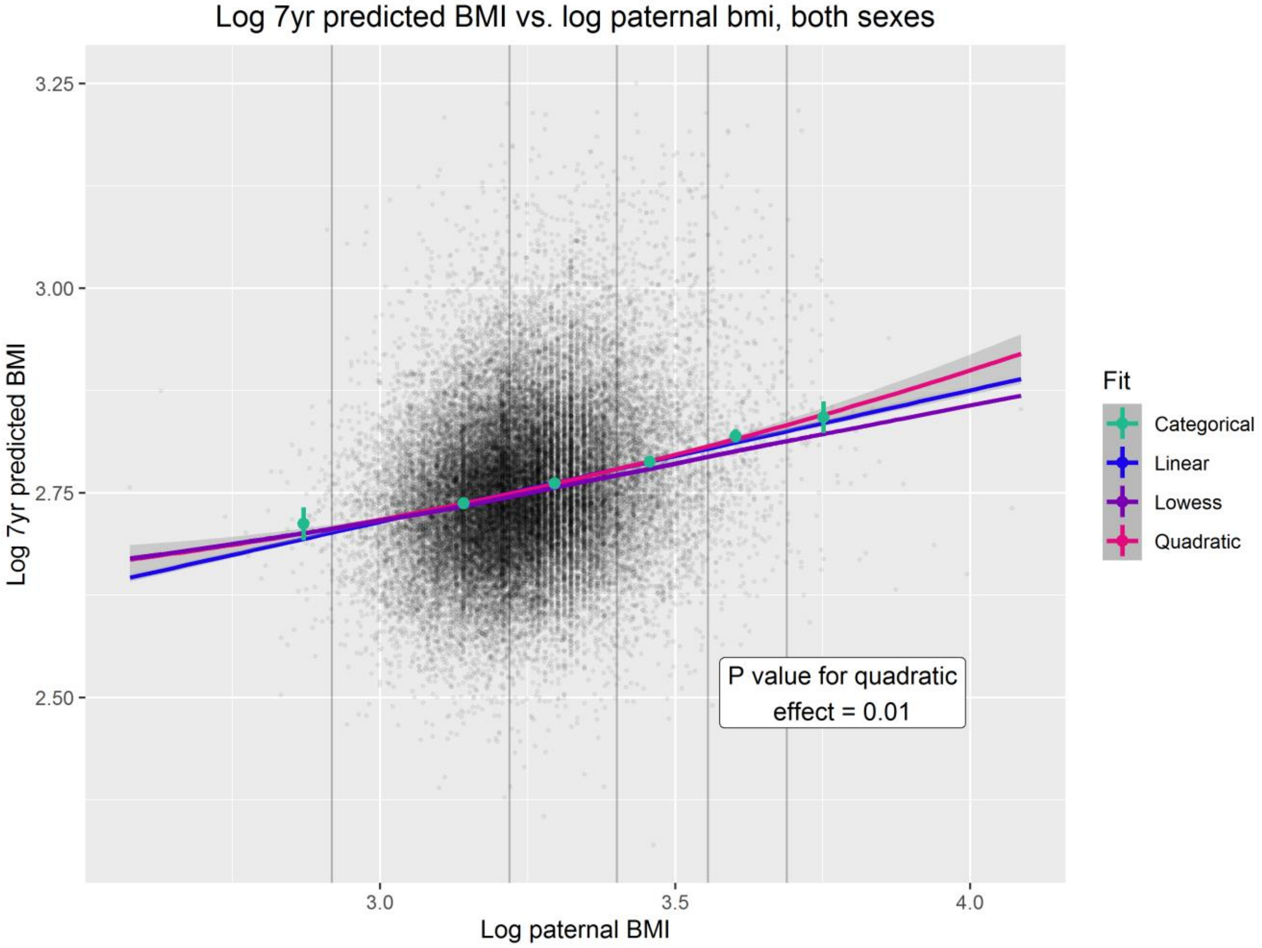
Log 7yr predicted BMI vs. log paternal bmi, both sexes
Log 7yr predicted BMI
3.25
3.00
2.75
2.50
Log paternal BMI
3.0
3.5
4.0
Fit
Categorical
Linear
Lowess
Quadratic
P value for quadratic effect = 0.01

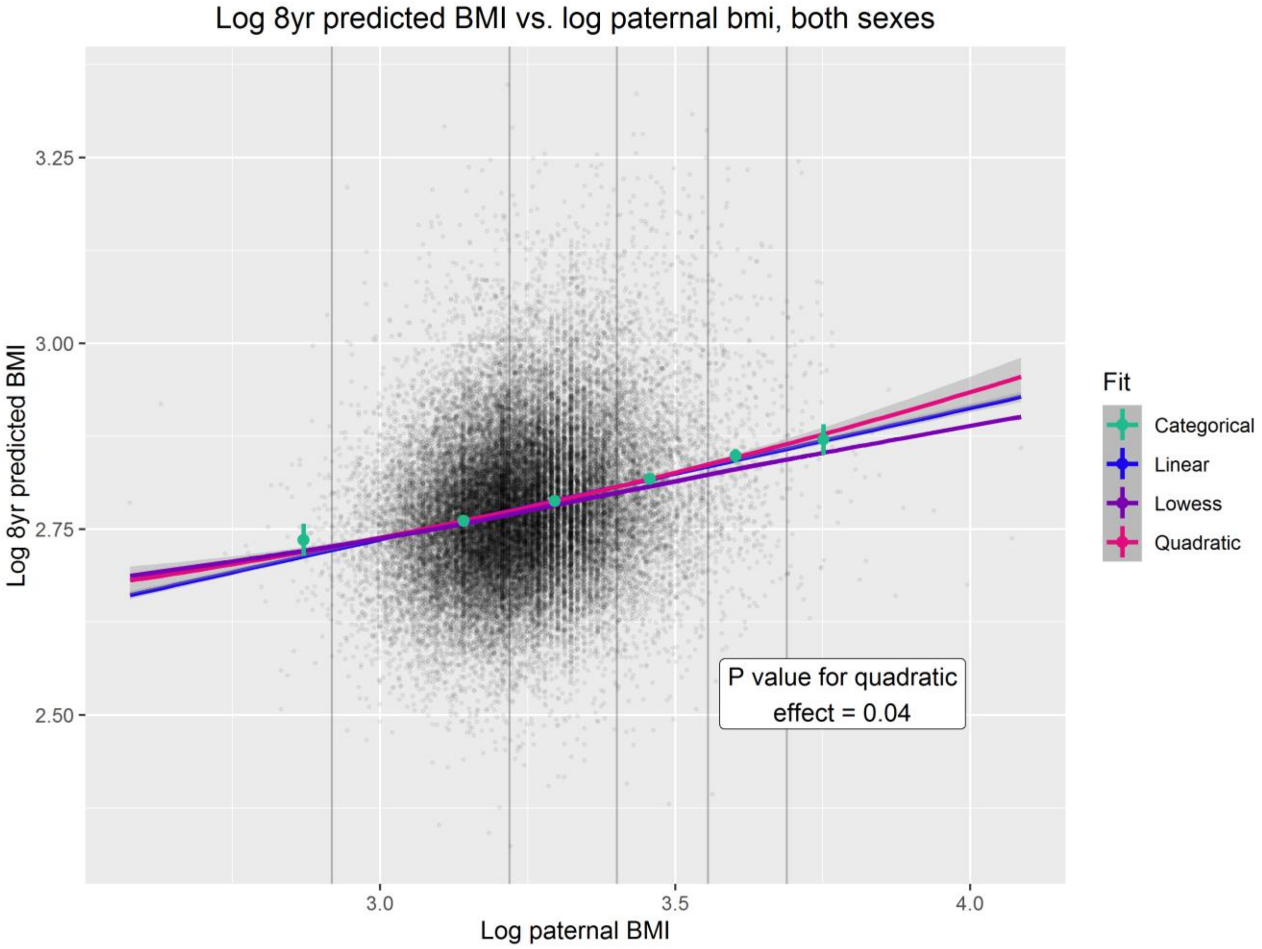
Log 8yr predicted BMI vs. log paternal bmi, both sexes
Log 8yr predicted BMI
3.25
3.00
2.75
2.50
3.0
3.5
4.0
Log paternal BMI
Fit
Categorical
Linear
Lowess
Quadratic
P value for quadratic effect = 0.04

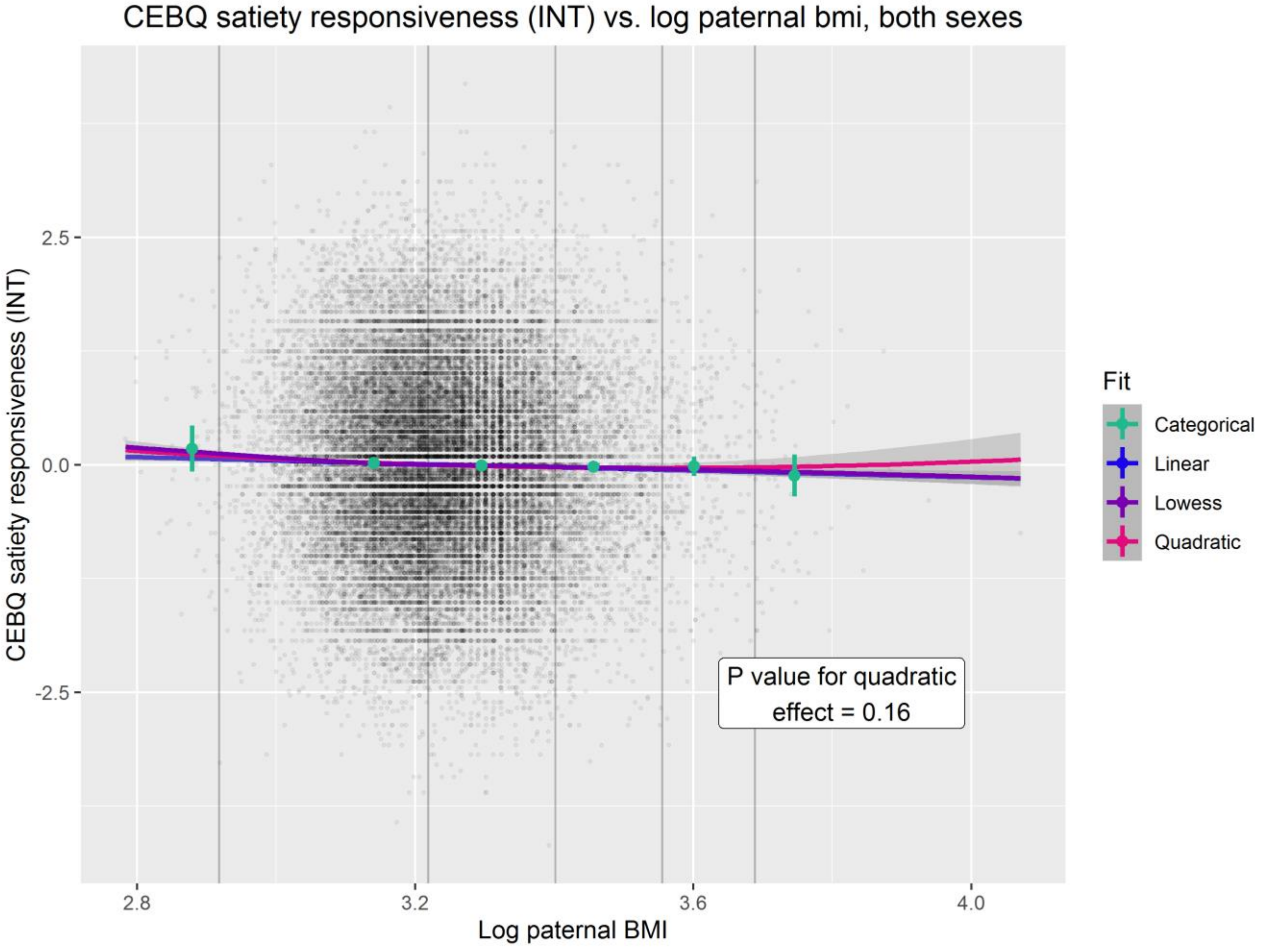
CEBQ satiety responsiveness (INT) vs. log paternal bmi, both sexes
CEBQ satiety responsiveness (INT)
2.5
0.0
-2.5
2.8
3.2
3.6
4.0
Log paternal BMI
Fit
Categorical
Linear
Lowess
Quadratic
P value for quadratic effect = 0.16

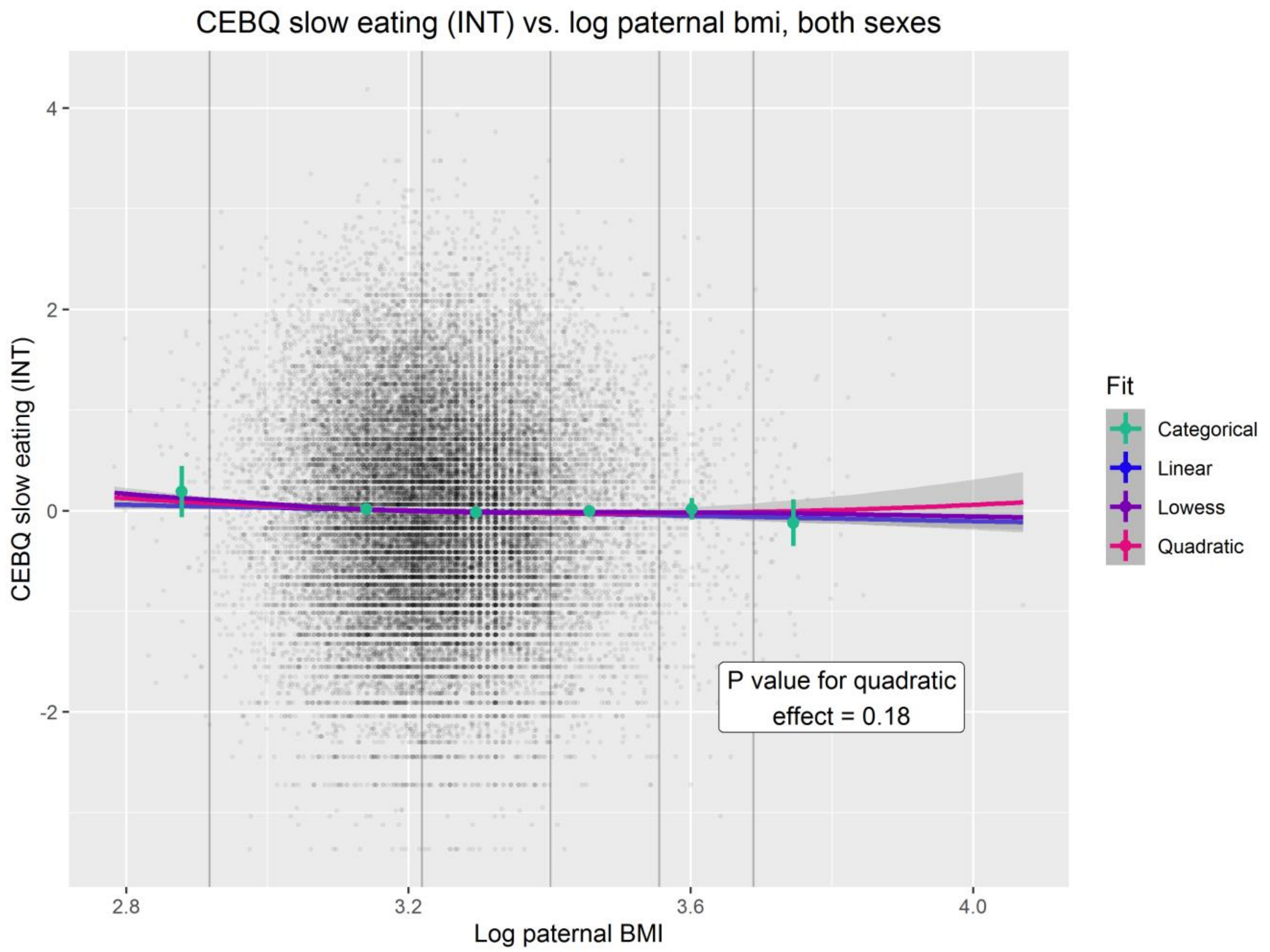
CEBQ slow eating (INT) vs. log paternal bmi, both sexes
CEBQ slow eating (INT)
Log paternal BMI
4
2
0
-2
2.8
3.2
3.6
4.0
Fit
Categorical
Linear
Lowess
Quadratic
P value for quadratic effect = 0.18

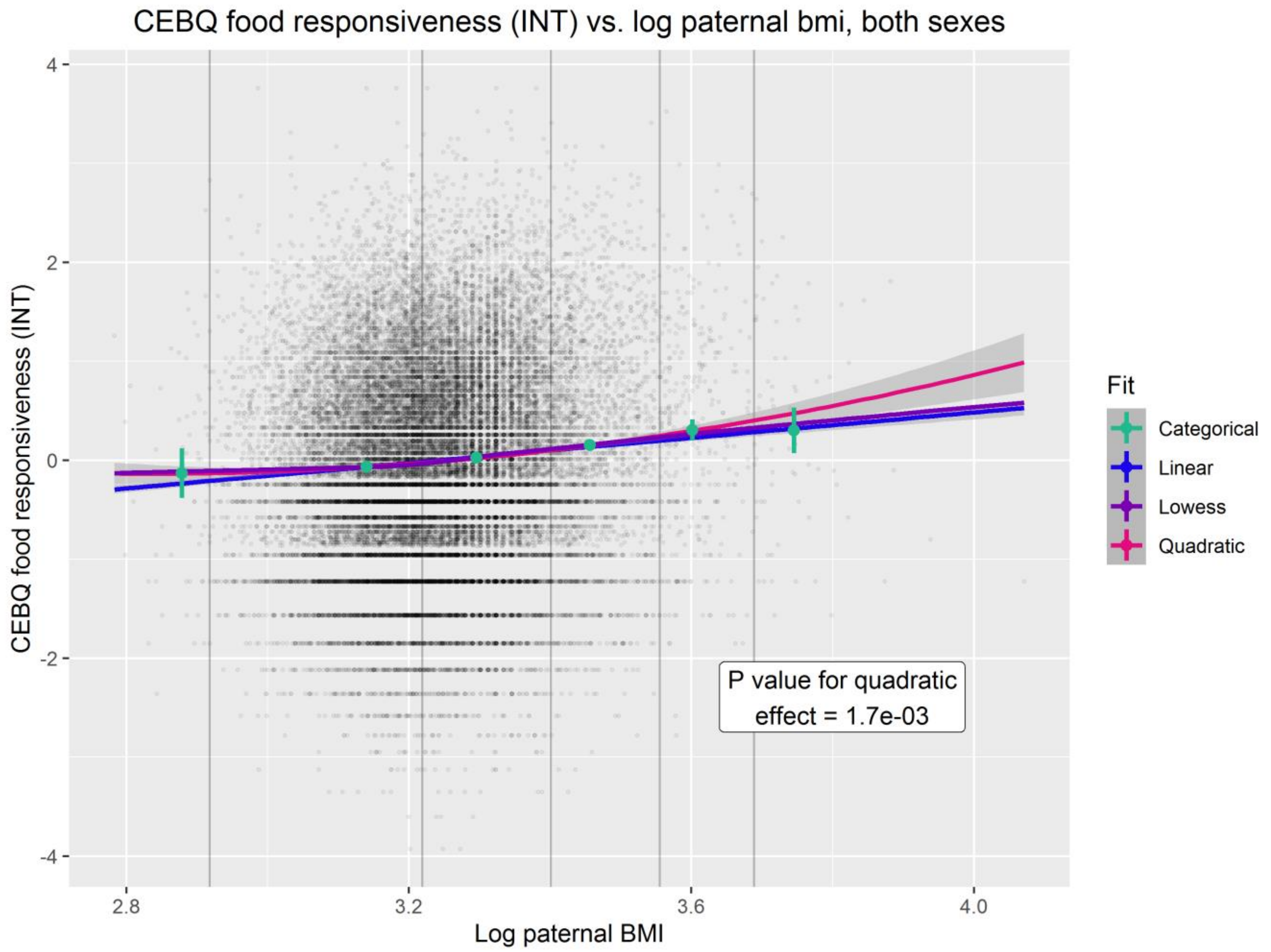
CEBQ food responsiveness (INT) vs. log paternal bmi, both sexes
CEBQ food responsiveness (INT)
Log paternal BMI
4
2
0
-2
-4
2.8
3.2
3.6
4.0
Fit
Categorical
Linear
Lowess
Quadratic
P value for quadratic effect = 1.7e-03

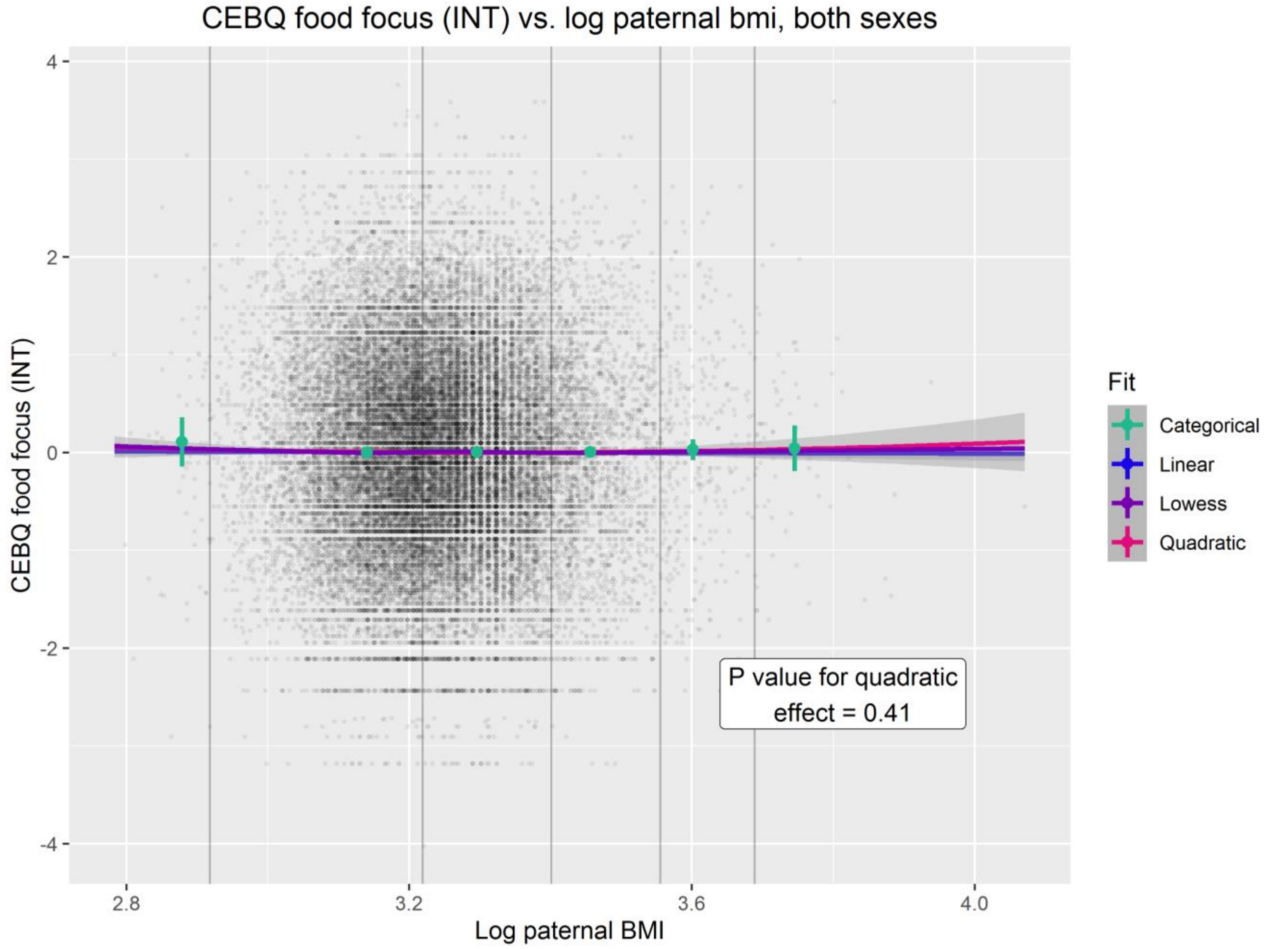
CEBQ food focus (INT) vs. log paternal bmi, both sexes
CEBQ food focus (INT)
Log paternal BMI
4
2
0
-2
-4
2.8
3.2
3.6
4.0
Fit
Categorical
Linear
Lowess
Quadratic
P value for quadratic effect = 0.41

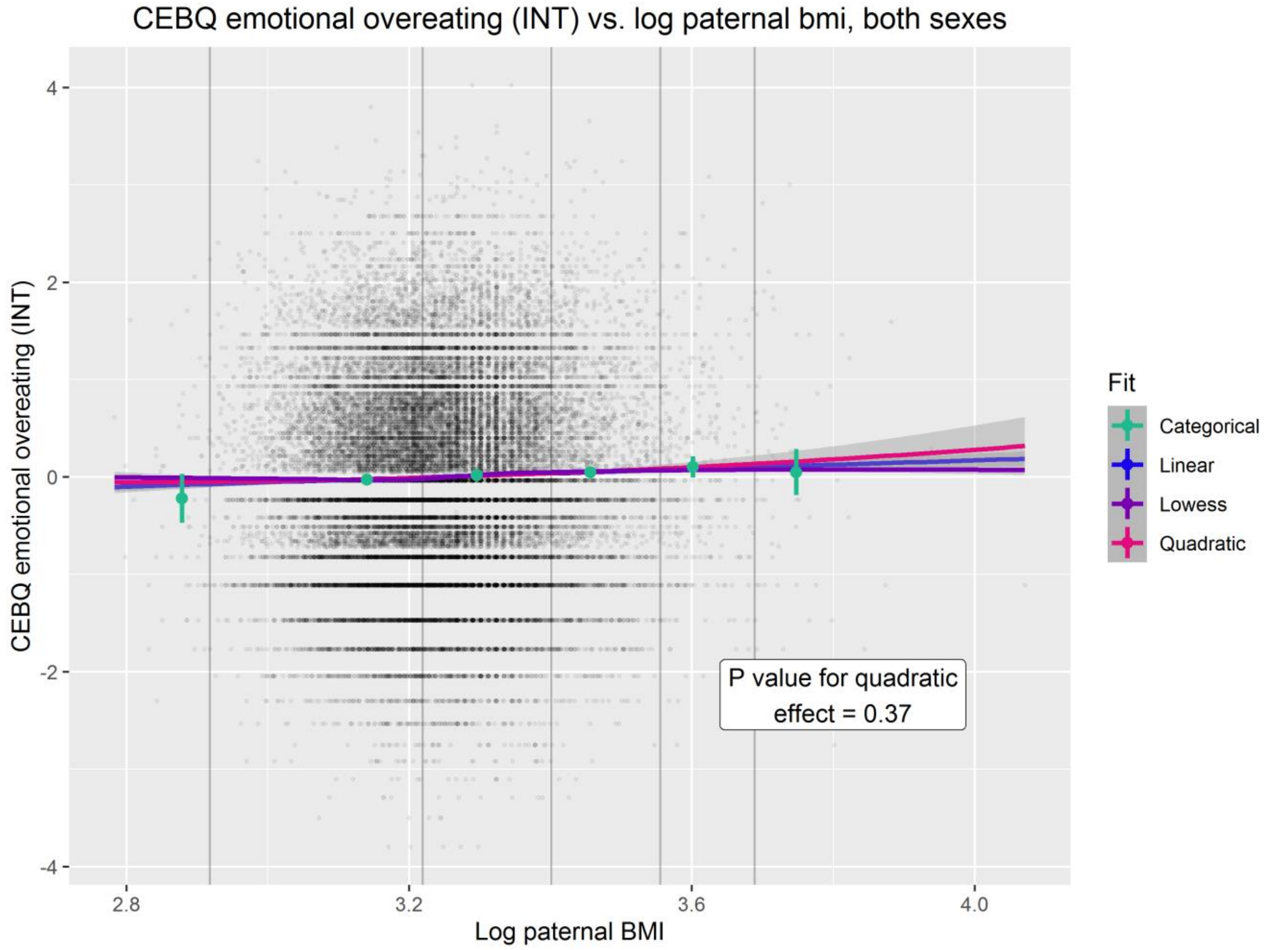
CEBQ emotional overeating (INT) vs. log paternal bmi, both sexes
CEBQ emotional overeating (INT)
Log paternal BMI
4
2
0
-2
-4
2.8
3.2
3.6
4.0
Fit
Categorical
Linear
Lowess
Quadratic
P value for quadratic effect = 0.37

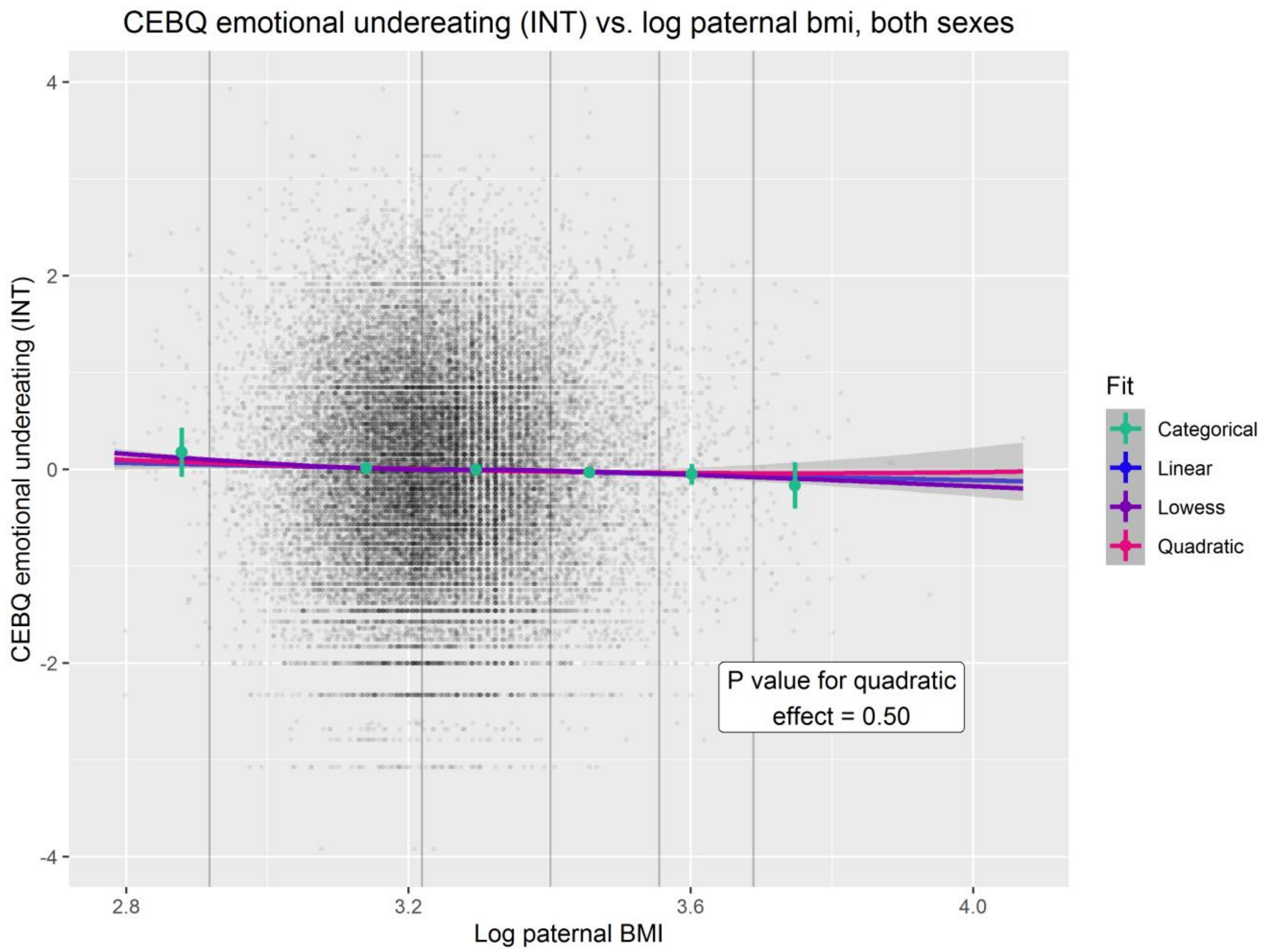
CEBQ emotional undereating (INT) vs. log paternal bmi, both sexes
CEBQ emotional undereating (INT)
4
2
0
-2
-4
2.8
3.2
3.6
4.0
Log paternal BMI
Fit
Categorical
Linear
Lowess
Quadratic
P value for quadratic effect = 0.50

### Supplementary information S8: Multiple Children of Twins and Siblings (MCoTS) model

To quantify the extent to which exposure-outcome associations were due to genetic confounding, we fitted a genetically informed structural equation model (SEM) in a subset of the MoBa sample. This SEM is an extension of the classic twin design (CTD) which has been widely used to estimate heritability (i.e. the proportion of phenotypic variance that is due to genetic effects). Because monozygotic (MZ) twins are genetically identical but dizygotic (DZ) twins are on average only 50% genetically similar, the expected phenotypic similarity of a heritable trait will be greater for MZ versus DZ twins. Building on the CTD, a Children of Twins design exploits the fact that such patterns of genetic and phenotypic similarity are mirrored in relatives of twins (5). This enables estimation of the extent to which phenotypic similarity between parents and offspring is due to genetic similarity (i.e. genetic confounding). For example, an MZ twin is 50% genetically similar to the child of their co-twin, but a DZ twin will be on average only 25% genetically similar to the child of their co-twin. If these relatedness coefficients are reflected by patterns of phenotypic similarity this provides evidence for genetic confounding. We fit an extended children of twins SEM (the Multiple Children of Twins and Siblings [MCoTS] model, described in the path diagrams below and elsewhere (5)).

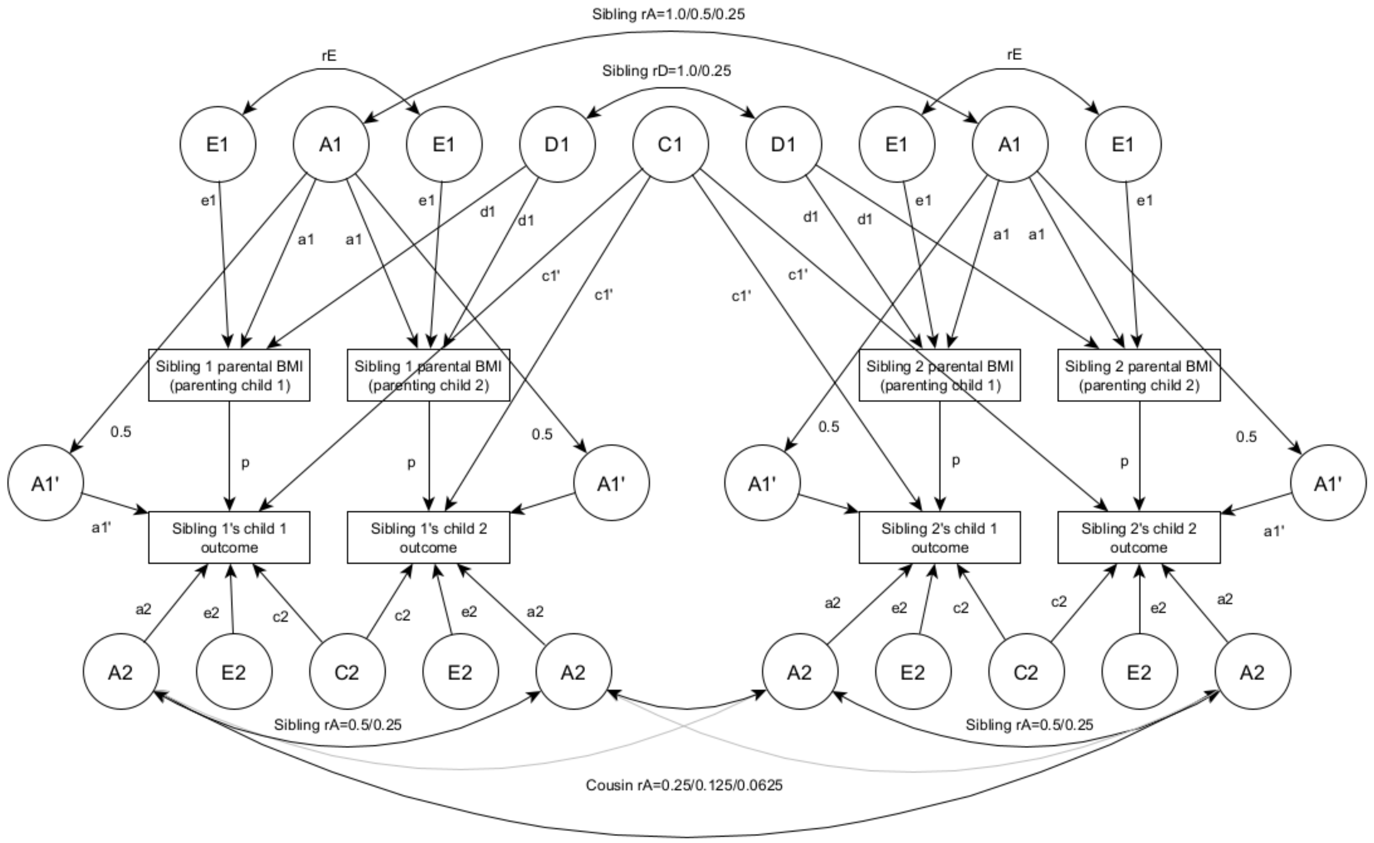


Sibling 1 and sibling 2 are full siblings, half siblings, dizygotic twins or monozygotic twins, in the parent generation. Outcomes are included for up to two children of each parental sibling, and parental BMI is allowed to vary between pregnancies via the correlation rE. Genetic correlations are set according to quantitative genetic theory (6). Additive genetic correlations between sibling pairs (Sibling rA) are set at 1.0, 0.5 or 0.25 for monozygotic twins, dizygotic twins/full siblings and half siblings respectively. Additive genetic correlations between cousin pairs (Cousin rA) are set at 0.25, 0.125 or 0.0625 for cousins related via monozygotic twins, dizygotic twins/full siblings and half siblings respectively. Dominance genetic correlations between sibling pairs (Sibling rD) are set at 1.0 or 0.25 for monozygotic twins and dizygotic twins/full siblings respectively and are zero for half siblings. Latent (unmeasured) variables are shown in circles, measured variables are shown in rectangles, single headed arrows denote causal paths, and double headed arrows denote covariances. A1: additive genetic effects on parental BMI, D1: dominance genetic effects on parental BMI, E1: nonshared environmental effects on parental BMI, A1': genetic effects common to parental BMI and offspring outcome, C1: extended family effects, A2: additive genetic effects specific to the offspring outcome, C2: shared environmental effects on offspring outcome, E2: nonshared environmental effect son offspring outcome, p: "phenotypic" effect, including any causal effect of parental BMI on the offspring outcome, as well as residual (non-genetic) confounding, rE: within-parent correlation between E1 for parenting child 1 and child 2. The path between A1 and A1' is fixed to 0.5, because parents share 50% of their genome identical by descent with their children. For simplicity, variance paths have been omitted, but variance was 1 for all latent variables. Consequently, for A1' residual variance after accounting for the path between A1' and A1 is 0.75. MCoTS SEM were fit in R version 4.0.3 (OpenMx package version 2.18.1) (7, 8). A full description of the MCoTS model is given in McAdams *et al*. (5).

**Supplementary information S9: MCoTS model with ACE partition of parental exposure**

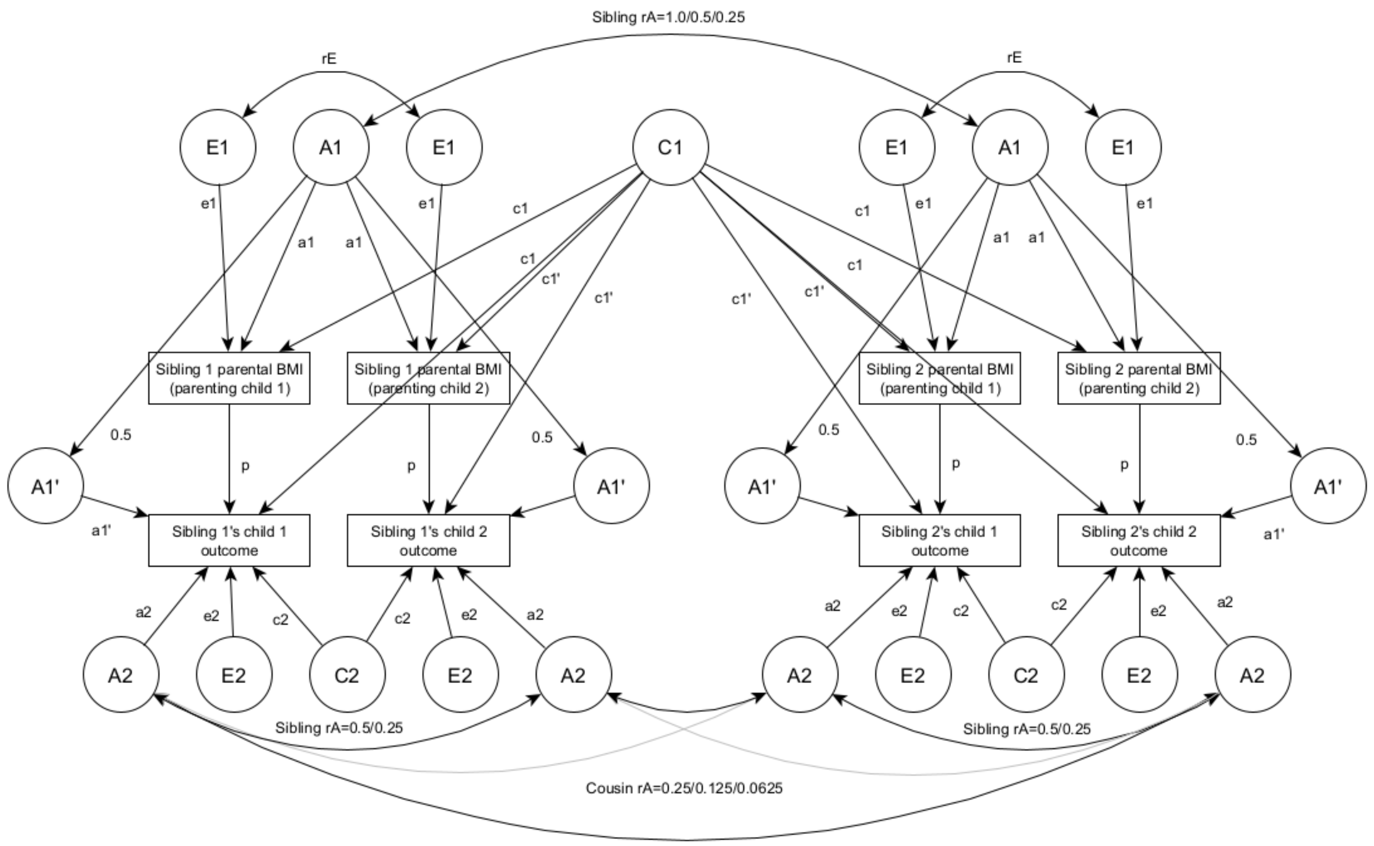


Sibling 1 and sibling 2 are full siblings, half siblings, dizygotic twins or monozygotic twins, in the parent generation. Outcomes are included for up to two children of each parental sibling, and parental BMI is allowed to vary between pregnancies via the correlation rE. Genetic correlations are set according to quantitative genetic theory (6). Additive genetic correlations between sibling pairs (Sibling rA) are set at 1.0, 0.5 or 0.25 for monozygotic twins, dizygotic twins/full siblings and half siblings respectively. Additive genetic correlations between cousin pairs (Cousin rA) are set at 0.25, 0.125 or 0.0625 for cousins related via monozygotic twins, dizygotic twins/full siblings and half siblings respectively. Latent (unmeasured) variables are shown in circles, measured variables are shown in rectangles, single headed arrows denote causal paths, and double headed arrows denote covariances. A1: additive genetic effects on parental BMI, C1: shared environmental effects on parental BMI, E1: nonshared environmental effects on parental BMI, A1': genetic effects common to parental BMI and offspring outcome, C1: extended family effects, A2: additive genetic effects specific to the offspring outcome, C2: shared environmental effects on offspring outcome, E2: nonshared environmental effect son offspring outcome, p: "phenotypic" effect, including any causal effect of parental BMI on the offspring outcome, as well as residual (non-genetic) confounding, rE: within-parent correlation between E1 for parenting child 1 and child 2. The path between A1 and A1' is fixed to 0.5, because parents share 50% of their genome identical by descent with their children. For simplicity, variance paths have been omitted, but variance was 1 for all latent variables. Consequently, for A1' residual variance after accounting for the path between A1' and A1 is 0.75. MCoTS SEM were fit in R version 4.0.3 (OpenMx package version 2.18.1) (7, 8). A full description of the MCoTS model is given in McAdams *et al*. (5).

**Supplementary information S10: MCoTS model with AE partition of parental exposure**

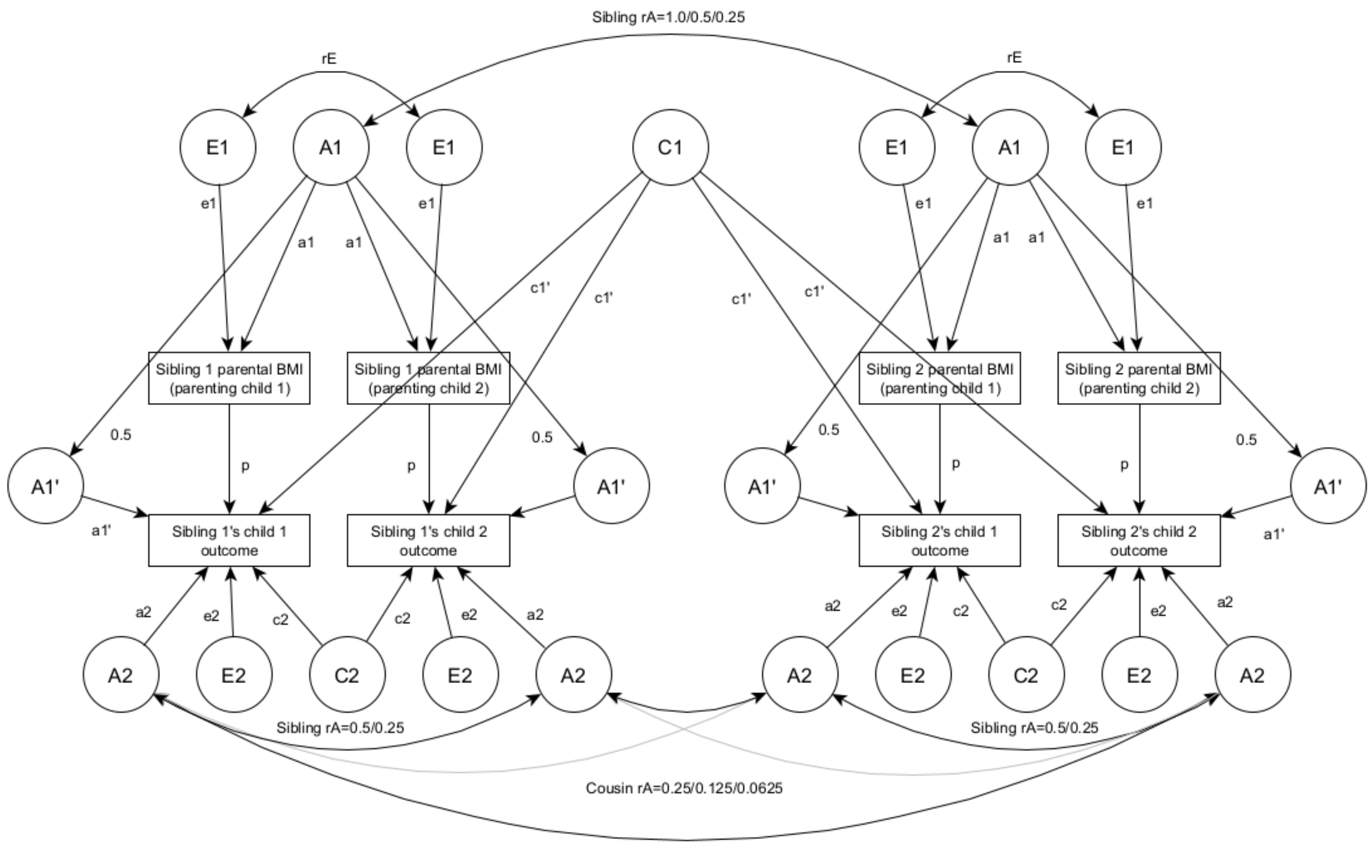


Sibling 1 and sibling 2 are full siblings, half siblings, dizygotic twins or monozygotic twins, in the parent generation. Outcomes are included for up to two children of each parental sibling, and parental BMI is allowed to vary between pregnancies via the correlation rE. Genetic correlations are set according to quantitative genetic theory (6). Additive genetic correlations between sibling pairs (Sibling rA) are set at 1.0, 0.5 or 0.25 for monozygotic twins, dizygotic twins/full siblings and half siblings respectively. Additive genetic correlations between cousin pairs (Cousin rA) are set at 0.25, 0.125 or 0.0625 for cousins related via monozygotic twins, dizygotic twins/full siblings and half siblings respectively. Latent (unmeasured) variables are shown in circles, measured variables are shown in rectangles, single headed arrows denote causal paths, and double headed arrows denote covariances. A1: additive genetic effects on parental BMI, E1: nonshared environmental effects on parental BMI, A1': genetic effects common to parental BMI and offspring outcome, C1: extended family effects, A2: additive genetic effects specific to the offspring outcome, C2: shared environmental effects on offspring outcome, E2: nonshared environmental effect son offspring outcome, p: "phenotypic" effect, including any causal effect of parental BMI on the offspring outcome, as well as residual (non-genetic) confounding, rE: within-parent correlation between E1 for parenting child 1 and child 2. The path between A1 and A1' is fixed to 0.5, because parents share 50% of their genome identical by descent with their children. For simplicity, variance paths have been omitted, but variance was 1 for all latent variables. Consequently, for A1' residual variance after accounting for the path between A1' and A1 is 0.75. MCoTS SEM were fit in R version 4.0.3 (OpenMx package version 2.18.1) (7, 8). A full description of the MCoTS model is given in McAdams *et al*. (5).

**Supplementary information S11: Liability threshold model for untransformed CEBQ outcomes**

Because several CEBQ outcomes were severely skewed, for the primary MCoTS analysis we regressed all CEBQ outcome variables on age and sex then applied a rank-based inverse normal transformation to the residuals. As a sensitivity analysis we instead applied a liability threshold MCoTS model to CEBQ outcomes that were recoded into three equally sized categories. Liability threshold models assume that a continuous, normally distributed liability captures all latent genetic and environmental determinants of the trait. The observed categorical outcome ($Y$) is related to the continuous liability ($Z$) by thresholds $\tau_1$ and $\tau_2$, such that

$$Y = \begin{cases} 2 \; if \; Z \geq \tau_2 \\ 1 \; if \; \tau_2 < Z \geq \tau_1 \\ 0 \; otherwise \end{cases}.$$

Liability threshold MCoTS SEM were fit in R version 4.0.3, similarly to the MCoTS models for continuous outcomes (OpenMx package version 2.18.1) (7, 8). The means vectors for offspring outcomes were fixed to zero and the outcome phenotypic variance was constrained to one, with $\tau_1$ and $\tau_2$ being freely estimated.

**Supplementary information S12: Differences in participant characteristics between baseline and 8 year old sample**

| Variable | | Baseline sample | | 8 year sample | | $P_{\text{difference}}$ |
|---|---|---|---|---|---|---|
| | | Mean | % | Mean | % | |
| Parental characteristics | | | | | | |
| Maternal BMI (kg/m$^2$) | | 24.1 | | 23.9 | | 8.1e-16 |
| Paternal BMI (kg/m$^2$) | | 25.9 | | 25.8 | | 4.4e-08 |
| Maternal WHO BMI category (kg/m$^2$) | <18.5 | | 3.0 | | 2.7 | 4.5e-31 |
| | 18.5–24.9 | | 65.6 | | 66.9 | |
| | 25–29.9 | | 21.8 | | 21.7 | |
| | ≥30 | | 9.6 | | 8.6 | |
| Paternal WHO BMI category (kg/m$^2$) | <18.5 | | 0.2 | | 0.2 | 5.5e-11 |
| | 18.5–24.9 | | 44.0 | | 44.7 | |
| | 25–29.9 | | 45.6 | | 45.5 | |
| | ≥30 | | 10.2 | | 9.6 | |
| Parity (number of previous births) | 0 | | 45.1 | | 45.9 | 6.5e-09 |
| | 1 | | 35.9 | | 35.6 | |
| | 2 | | 14.9 | | 14.6 | |
| | 3 | | 3.2 | | 3.0 | |
| | 4+ | | 0.9 | | 0.8 | |
| Maternal age at birth of child (years) | ≤19 | | 0.7 | | 0.5 | 4.0e-147 |
| | 20-24 | | 9.4 | | 7.4 | |
| | 25-29 | | 32.9 | | 32.4 | |
| | 30-34 | | 39.4 | | 40.9 | |
| | 35-39 | | 15.7 | | 16.7 | |
| | ≥40 | | 1.9 | | 2.1 | |
| Maternal smoking during pregnancy | No | | 92.4 | | 94.4 | 3.6e-130 |
| | Yes | | 7.6 | | 5.6 | |
| Paternal smoking during pregnancy | No | | 76.4 | | 78.4 | 8.5e-54 |
| | Yes | | 23.6 | | 21.6 | |
| Maternal educational attainment | Incomplete upper 2° school | | 0.2 | | 0.2 | <2.2e-308 |
| | Upper 2° school | | 1.8 | | 1.0 | |
| | High school/junior college | | 27.9 | | 23.5 | |
| | University/college, 4 years | | 42.3 | | 45.0 | |
| | University/college, >4 years | | 26.3 | | 28.9 | |
| | Other | | 1.6 | | 1.4 | |
| Maternal income | No income | | 2.3 | | 1.7 | 1.5e-198 |
| | <150,000 NOK | | 15.1 | | 13.0 | |
| | 150,000–199,999 NOK | | 11.1 | | 9.9 | |
| | 200,000–299,999 NOK | | 34.3 | | 34.6 | |
| | 300,000–399,999 NOK | | 25.6 | | 28.1 | |
| | 400,000–499,999 NOK | | 7.1 | | 7.7 | |
| | >500,000 NOK | | 4.5 | | 4.9 | |
| Parental language | Norwegian | | 89.1 | | 90.0 | 1.3e-17 |
| | Other | | 10.9 | | 10.0 | |
| Offspring characteristics | | | | | | |
| Gestational age (weeks) | | 39.8 | | 39.8 | | 4.7e-17 |
| Birth weight (g) | | 3563 | | 3572 | | 3.9e-07 |

Statistics are for the "Baseline" sample used for linear regression analyses of birth weight (*n* = 85,866), and an "8 year" sample similar to that used for linear regression analyses of 8 year BMI (*n* = 46,539). $\boldsymbol{P}_{\textbf{difference}}$: for continuous variables: *P*-value from a linear regression model testing the null hypothesis that variables have equal mean in both samples; for categorical variables: *P*-value from a chi squared test of the null hypothesis that variables are equally distributed in both samples, **NOK**: Norwegian Krone

**Supplementary information S13: Linear associations between parental BMI and offspring predicted BMI**

| Offspring outcome | Exposure | *n* | Estimate | 95% CI | *P* | $P_{dif}$ |
|---|---|---|---|---|---|---|
| Predicted 1 year BMI | Maternal BMI | 47524 | 0.10 | 0.09, 0.11 | 1.1e-101 | 0.05 |
| | Paternal BMI | 47524 | 0.09 | 0.08, 0.10 | 4.1e-77 | |
| Predicted 2 year BMI | Maternal BMI | 47524 | 0.10 | 0.09, 0.11 | 1.1e-95 | 0.30 |
| | Paternal BMI | 47524 | 0.09 | 0.08, 0.10 | 6.1e-83 | |
| Predicted 3 year BMI | Maternal BMI | 47524 | 0.11 | 0.10, 0.12 | 6.7e-117 | 0.51 |
| | Paternal BMI | 47524 | 0.11 | 0.10, 0.12 | 1.1e-108 | |
| Predicted 4 year BMI | Maternal BMI | 47524 | 0.13 | 0.12, 0.14 | 1.7e-166 | 0.50 |
| | Paternal BMI | 47524 | 0.13 | 0.12, 0.14 | 9.8e-157 | |
| Predicted 5 year BMI | Maternal BMI | 47524 | 0.15 | 0.14, 0.16 | 6.9e-220 | 0.36 |
| | Paternal BMI | 47524 | 0.15 | 0.14, 0.15 | 6.8e-204 | |
| Predicted 6 year BMI | Maternal BMI | 47524 | 0.17 | 0.16, 0.18 | 8.3e-270 | 0.19 |
| | Paternal BMI | 47524 | 0.16 | 0.15, 0.17 | 7e-244 | |
| Predicted 7 year BMI | Maternal BMI | 47524 | 0.18 | 0.17, 0.19 | <2.2e-308 | 0.09 |
| | Paternal BMI | 47524 | 0.17 | 0.16, 0.18 | 7.1e-275 | |
| Predicted 8 year BMI | Maternal BMI | 47524 | 0.18 | 0.17, 0.19 | <2.2e-308 | 0.05 |
| | Paternal BMI | 47524 | 0.17 | 0.16, 0.18 | 8.2e-285 | |

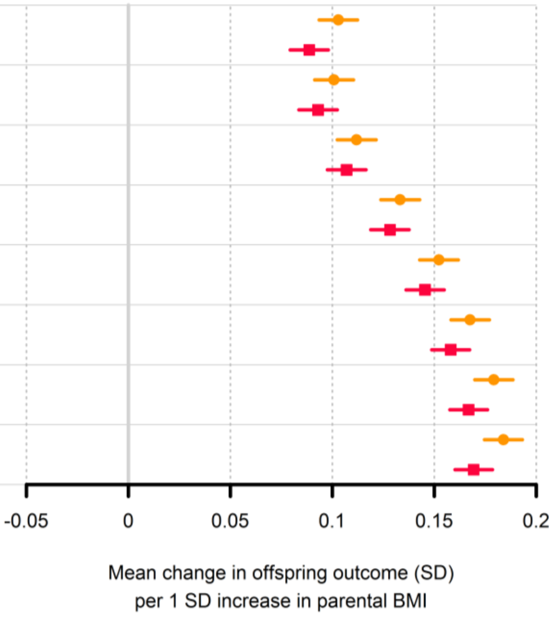


**Supplementary information S14: Linear associations between offspring 8 year BMI and CEBQ outcomes**

| CEBQ outcome | Beta[a] | Lower 95% CI | Upper 95% CI | *P* |
|---|---|---|---|---|
| CEBQ: food responsiveness | 0.37 | 0.36 | 0.38 | <2.2e-308 |
| CEBQ: fussiness | -0.09 | -0.10 | -0.08 | 8.2e-72 |
| CEBQ: satiety responsiveness | -0.26 | -0.27 | -0.25 | <2.2e-308 |
| CEBQ: slow eating | -0.14 | -0.15 | -0.13 | 6.7e-164 |
| CEBQ: emotional overeating | 0.16 | 0.15 | 0.17 | 3.1e-180 |
| CEBQ: emotional undereating | 0.00 | -0.01 | 0.01 | 0.44 |

**a**: regression coefficient from regression of offspring 8 year CEBQ outcome (regressed on offspring age and sex prior to inverse normalization of the residuals) on offspring 8 year BMI (*z*-score), adjusting for offspring age and sex, maternal parity, maternal and paternal covariates (age, smoking during pregnancy, educational attainment and income), parental language and grandparental language

### Supplementary information S15: Statistical interaction between maternal and paternal BMI

| Offspring outcome | Maternal BMI (z score)[a] | | | | Paternal BMI (z score)[b] | | | | Maternal-paternal BMI interaction[c] | | | |
|---|---|---|---|---|---|---|---|---|---|---|---|---|
| | Beta | Lower 95% CI | Upper 95% CI | *P* | Beta | Lower 95% CI | Upper 95% CI | *P* | Beta | Lower 95% CI | Upper 95% CI | *P* |
| Birth weight (z score) | 0.124 | 0.116 | 0.132 | 8.4e-220 | 0.018 | 0.010 | 0.025 | 5.5e-06 | -0.015 | -0.023 | -0.007 | 4.8e-07 |
| 6 month BMI (z score) | 0.090 | 0.081 | 0.099 | 3.1e-89 | 0.061 | 0.052 | 0.069 | 4.9e-43 | -0.011 | -0.020 | -0.003 | 1.1e-03 |
| 1 year BMI (z score) | 0.104 | 0.094 | 0.113 | 8e-101 | 0.075 | 0.065 | 0.084 | 1.4e-55 | -0.011 | -0.020 | -0.001 | 3.4e-03 |
| 2 year BMI (z score) | 0.087 | 0.074 | 0.100 | 2.5e-38 | 0.072 | 0.059 | 0.085 | 6.7e-27 | -0.007 | -0.020 | 0.006 | 0.17 |
| 3 year BMI (z score) | 0.083 | 0.071 | 0.095 | 2.1e-42 | 0.082 | 0.070 | 0.094 | 1.2e-43 | -0.005 | -0.017 | 0.007 | 0.31 |
| Log 5 year BMI (z score) | 0.142 | 0.129 | 0.155 | 5.6e-101 | 0.134 | 0.121 | 0.147 | 8.2e-98 | -0.004 | -0.017 | 0.009 | 0.47 |
| Log 8 year BMI (z score) | 0.202 | 0.191 | 0.212 | 1e-302 | 0.179 | 0.168 | 0.189 | 1.3e-252 | 0.002 | -0.008 | 0.013 | 0.60 |
| Predicted 1 year BMI (z score) | 0.107 | 0.097 | 0.118 | 6.6e-86 | 0.094 | 0.084 | 0.105 | 2.1e-70 | -0.012 | -0.023 | -0.002 | 3.9e-03 |
| Predicted 2 year BMI (z score) | 0.105 | 0.094 | 0.116 | 1e-80 | 0.099 | 0.088 | 0.109 | 5.4e-75 | -0.011 | -0.022 | 0.000 | 0.01 |
| Predicted 3 year BMI (z score) | 0.116 | 0.106 | 0.127 | 2.8e-98 | 0.113 | 0.102 | 0.123 | 1.8e-96 | -0.008 | -0.019 | 0.003 | 0.07 |
| Predicted 4 year BMI (z score) | 0.139 | 0.128 | 0.149 | 1.7e-139 | 0.134 | 0.123 | 0.145 | 8.5e-137 | -0.005 | -0.015 | 0.006 | 0.29 |
| Log predicted 5 year BMI (z score) | 0.159 | 0.148 | 0.170 | 2.2e-185 | 0.151 | 0.141 | 0.162 | 1.5e-176 | -0.005 | -0.015 | 0.006 | 0.29 |
| Log predicted 6 year BMI (z score) | 0.175 | 0.164 | 0.185 | 1e-226 | 0.164 | 0.154 | 0.175 | 5.8e-210 | -0.003 | -0.014 | 0.007 | 0.42 |
| Log predicted 7 year BMI (z score) | 0.187 | 0.176 | 0.197 | 2.7e-261 | 0.173 | 0.163 | 0.184 | 1.2e-236 | -0.004 | -0.014 | 0.007 | 0.39 |
| Log predicted 8 year BMI (z score) | 0.192 | 0.181 | 0.202 | 2.5e-275 | 0.177 | 0.166 | 0.187 | 1.9e-245 | -0.005 | -0.016 | 0.005 | 0.21 |
| CEBQ: food responsiveness (z score[d]) | 0.089 | 0.076 | 0.101 | 2.2e-44 | 0.063 | 0.050 | 0.075 | 6.2e-25 | 0.003 | -0.010 | 0.015 | 0.58 |
| CEBQ: fussiness (z score[d]) | -0.011 | -0.023 | 0.002 | 0.10 | -0.007 | -0.020 | 0.005 | 0.23 | 0.009 | -0.003 | 0.022 | 0.06 |
| CEBQ: satiety responsiveness (z score[d]) | 0.013 | 0.001 | 0.026 | 0.04 | -0.030 | -0.043 | -0.018 | 9e-07 | -0.002 | -0.014 | 0.011 | 0.70 |
| CEBQ: slow eating (z score[d]) | 0.013 | 0.000 | 0.025 | 0.04 | -0.022 | -0.034 | -0.009 | 4.1e-04 | 0.003 | -0.009 | 0.016 | 0.53 |
| CEBQ: emotional undereating (z score[d]) | -0.029 | -0.041 | -0.016 | 9.9e-06 | -0.012 | -0.025 | 0.001 | 0.05 | 0.008 | -0.005 | 0.020 | 0.14 |
| CEBQ: emotional overeating (z score[d]) | 0.039 | 0.027 | 0.052 | 1e-09 | 0.019 | 0.007 | 0.032 | 1.7e-03 | -0.001 | -0.013 | 0.012 | 0.90 |

**a**: regression coefficient from regression of offspring outcome on maternal BMI (*z*-score), adjusted for covariates as per the main linear regression analyses (Model 3), plus a maternal BMI × paternal BMI interaction term, **b**: regression coefficient from regression of offspring outcome on paternal BMI (*z*-score), adjusted for covariates as per the main linear regression analyses (Model 3), plus a maternal BMI × paternal BMI interaction term, **c**: regression coefficient for the maternal BMI (*z*-score) × paternal BMI (*z*-score) interaction term, **d**: CEBQ outcomes were regressed on offspring age and sex prior to inverse normalization of the residuals

**Supplementary information S16: MCoTS results for the association of parental BMI with offspring birth weight, adjusted for potential confounders**

| Model | Exposure | *n* | Covariance component | Estimate | 95% CI | *P* |
|---|---|---|---|---|---|---|
| 1 | Maternal BMI | 45752 | Phenotypic | 0.13 | 0.12, 0.14 | 1.7e-222 |
| | | | Genetic | -0.05 | -0.09, 0.01 | 0.07 |
| | Paternal BMI | 50999 | Phenotypic | 0.05 | 0.04, 0.05 | 1.6e-35 |
| | | | Genetic | -0.07 | -0.09, -0.04 | 3.3e-04 |
| 2 | Maternal BMI | 41211 | Phenotypic | 0.12 | 0.11, 0.13 | 1.4e-196 |
| | | | Genetic | -0.02 | -0.06, 0.03 | 0.44 |
| | Paternal BMI | 45259 | Phenotypic | 0.02 | 0.01, 0.02 | 9.9e-06 |
| | | | Genetic | -0.07 | -0.10, -0.04 | 1.1e-04 |
| 3 | Maternal BMI | 37787 | Phenotypic | 0.12 | 0.11, 0.13 | 1.7e-179 |
| | | | Genetic | 0.00 | -0.04, 0.05 | 0.91 |
| | Paternal BMI | 41216 | Phenotypic | 0.01 | 0.01, 0.02 | 3.9e-04 |
| | | | Genetic | -0.04 | -0.08, -0.01 | 0.07 |

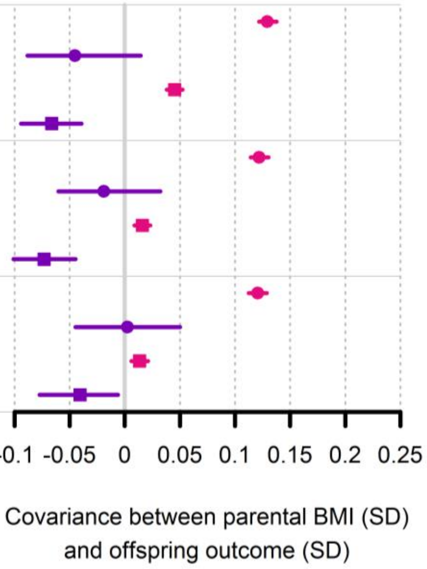


**Model 1**: not adjusted for potential confounders (as per main MCoTS analyses), **Model 2**: exposure and outcome adjusted (via linear regression) for the other parent's BMI, **Model 3**: exposure and outcome adjusted (via linear regression) for the other parent's BMI and the index parent's age and income

**Supplementary information S17: MCoTS results for the association of parental BMI with offspring birth weight and BMI**

| Offspring outcome | Exposure | *n* | Covariance component | Estimate | 95% CI | *P* |
|---|---|---|---|---|---|---|
| Birth weight | Maternal BMI | 45752 | Phenotypic | 0.13 | 0.12, 0.14 | 1.7e-222 |
| | | | Genetic | -0.05 | -0.09, 0.01 | 0.07 |
| | Paternal BMI | 50999 | Phenotypic | 0.05 | 0.04, 0.05 | 1.6e-35 |
| | | | Genetic | -0.07 | -0.09, -0.04 | 3.3e-04 |
| 6 Month BMI | Maternal BMI | 35913 | Phenotypic | 0.10 | 0.09, 0.11 | 9.3e-107 |
| | | | Genetic | 0.06 | 0.02, 0.10 | 0.02 |
| | Paternal BMI | 39529 | Phenotypic | 0.08 | 0.07, 0.08 | 2.3e-70 |
| | | | Genetic | 0.05 | 0.01, 0.09 | 0.03 |
| 1yr BMI | Maternal BMI | 31100 | Phenotypic | 0.13 | 0.12, 0.14 | 4.8e-154 |
| | | | Genetic | 0.13 | 0.09, 0.17 | 1.4e-06 |
| | Paternal BMI | 34210 | Phenotypic | 0.09 | 0.08, 0.10 | 4e-85 |
| | | | Genetic | 0.08 | 0.03, 0.11 | 1.7e-03 |
| 2yr BMI | Maternal BMI | 17001 | Phenotypic | 0.10 | 0.09, 0.11 | 3.4e-56 |
| | | | Genetic | 0.14 | 0.08, 0.19 | 3.9e-04 |
| | Paternal BMI | 18653 | Phenotypic | 0.08 | 0.07, 0.09 | 2.2e-43 |
| | | | Genetic | 0.13 | 0.07, 0.17 | 6.5e-05 |
| 3yr BMI | Maternal BMI | 21512 | Phenotypic | 0.10 | 0.09, 0.11 | 1.8e-62 |
| | | | Genetic | 0.18 | 0.14, 0.22 | 1.8e-10 |
| | Paternal BMI | 23319 | Phenotypic | 0.09 | 0.08, 0.10 | 1.8e-56 |
| | | | Genetic | 0.18 | 0.12, 0.22 | 1.4e-09 |
| 5yr BMI | Maternal BMI | 18194 | Phenotypic | 0.16 | 0.15, 0.18 | 2.4e-140 |
| | | | Genetic | 0.14 | 0.06, 0.19 | 1.1e-03 |
| | Paternal BMI | 19393 | Phenotypic | 0.15 | 0.14, 0.16 | 5.8e-146 |
| | | | Genetic | 0.12 | 0.04, 0.19 | 2.7e-03 |
| 8yr BMI | Maternal BMI | 24954 | Phenotypic | 0.24 | 0.23, 0.25 | <2.2e-308 |
| | | | Genetic | 0.19 | 0.15, 0.22 | 1.9e-12 |
| | Paternal BMI | 27051 | Phenotypic | 0.21 | 0.20, 0.22 | <2.2e-308 |
| | | | Genetic | 0.20 | 0.15, 0.24 | 2.7e-14 |

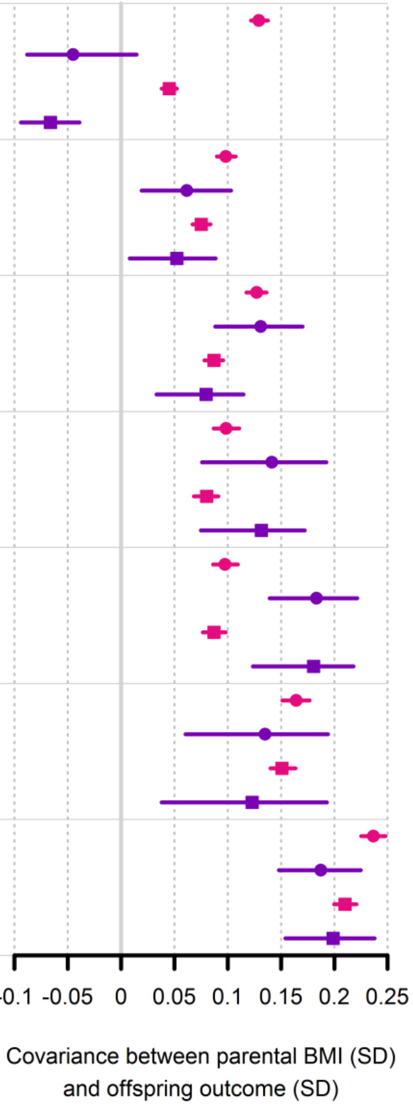

**Supplementary information S18: MCoTS results for the association of parental BMI with offspring predicted BMI**

| Offspring outcome | Exposure | *n* | Covariance component | Estimate | 95% CI | *P* |
|---|---|---|---|---|---|---|
| Predicted 1yr BMI | Maternal BMI | 25347 | Phenotypic | 0.13 | 0.12, 0.15 | 8.5e-130 |
| | | | Genetic | 0.13 | 0.08, 0.17 | 5.5e-05 |
| | Paternal BMI | 27530 | Phenotypic | 0.11 | 0.10, 0.12 | 3.5e-100 |
| | | | Genetic | 0.10 | 0.05, 0.13 | 5.3e-04 |
| Predicted 2yr BMI | Maternal BMI | 25347 | Phenotypic | 0.13 | 0.12, 0.14 | 1.9e-113 |
| | | | Genetic | 0.12 | 0.07, 0.16 | 6e-05 |
| | Paternal BMI | 27530 | Phenotypic | 0.11 | 0.10, 0.12 | 8e-103 |
| | | | Genetic | 0.12 | 0.07, 0.15 | 1.9e-05 |
| Predicted 3yr BMI | Maternal BMI | 25347 | Phenotypic | 0.14 | 0.13, 0.15 | 3.6e-134 |
| | | | Genetic | 0.13 | 0.08, 0.17 | 3.1e-06 |
| | Paternal BMI | 27530 | Phenotypic | 0.13 | 0.12, 0.14 | 5.1e-131 |
| | | | Genetic | 0.14 | 0.09, 0.17 | 2.6e-07 |
| Predicted 4yr BMI | Maternal BMI | 25347 | Phenotypic | 0.17 | 0.16, 0.18 | 7.5e-187 |
| | | | Genetic | 0.16 | 0.11, 0.20 | 8.4e-08 |
| | Paternal BMI | 27530 | Phenotypic | 0.15 | 0.14, 0.16 | 4.9e-185 |
| | | | Genetic | 0.15 | 0.10, 0.18 | 7.5e-09 |
| Predicted 5yr BMI | Maternal BMI | 25347 | Phenotypic | 0.20 | 0.19, 0.21 | 3.5e-249 |
| | | | Genetic | 0.18 | 0.13, 0.23 | 2.2e-09 |
| | Paternal BMI | 27530 | Phenotypic | 0.18 | 0.17, 0.19 | 2.2e-246 |
| | | | Genetic | 0.16 | 0.11, 0.19 | 8.6e-10 |
| Predicted 6yr BMI | Maternal BMI | 25347 | Phenotypic | 0.22 | 0.21, 0.23 | 9.7e-301 |
| | | | Genetic | 0.20 | 0.15, 0.24 | 6.5e-11 |
| | Paternal BMI | 27530 | Phenotypic | 0.20 | 0.19, 0.21 | 1.4e-296 |
| | | | Genetic | 0.17 | 0.12, 0.21 | 7.1e-11 |
| Predicted 7yr BMI | Maternal BMI | 25347 | Phenotypic | 0.24 | 0.23, 0.25 | <2.2e-308 |
| | | | Genetic | 0.20 | 0.15, 0.25 | 8.4e-12 |
| | Paternal BMI | 27530 | Phenotypic | 0.21 | 0.20, 0.22 | <2.2e-308 |
| | | | Genetic | 0.18 | 0.13, 0.22 | 1.1e-11 |
| Predicted 8yr BMI | Maternal BMI | 25347 | Phenotypic | 0.24 | 0.23, 0.26 | <2.2e-308 |
| | | | Genetic | 0.20 | 0.15, 0.24 | 1.1e-12 |
| | Paternal BMI | 27530 | Phenotypic | 0.22 | 0.21, 0.23 | <2.2e-308 |
| | | | Genetic | 0.19 | 0.13, 0.23 | 8.6e-13 |

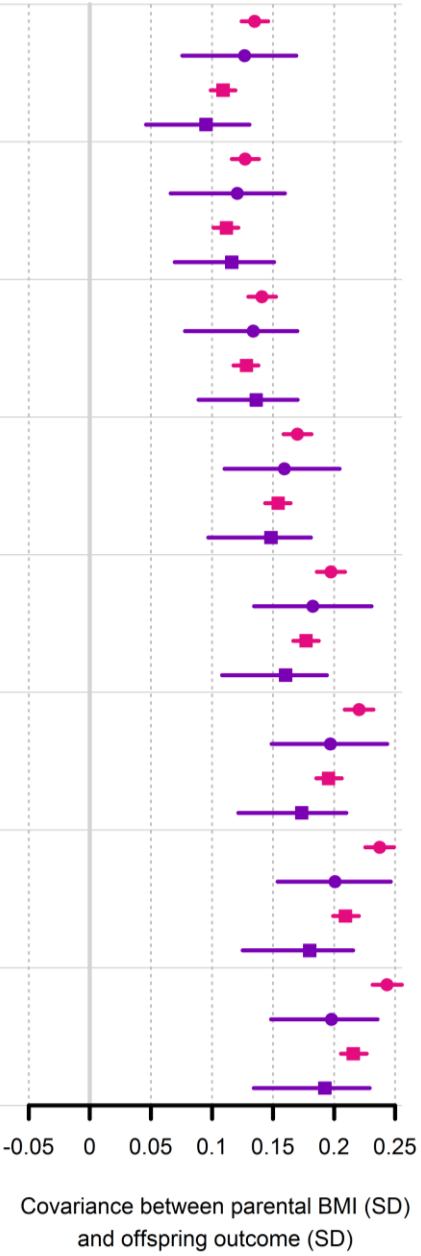

**Supplementary information S19: MCoTS results for the association of parental BMI with offspring weight, BMI and ponderal index at birth**

| Offspring outcome | Exposure | *n* | Covariance component | Estimate | 95% CI | *P* |
|---|---|---|---|---|---|---|
| Birth weight | Maternal BMI | 45752 | Phenotypic | 0.13 | 0.12, 0.14 | 1.7e-222 |
| | | | Genetic | -0.05 | -0.09, 0.01 | 0.07 |
| | Paternal BMI | 50999 | Phenotypic | 0.05 | 0.04, 0.05 | 1.6e-35 |
| | | | Genetic | -0.07 | -0.09, -0.04 | 3.3e-04 |
| Birth BMI | Maternal BMI | 44906 | Phenotypic | 0.12 | 0.11, 0.13 | 2.1e-193 |
| | | | Genetic | -0.03 | -0.08, 0.03 | 0.22 |
| | Paternal BMI | 50029 | Phenotypic | 0.05 | 0.04, 0.06 | 1.3e-39 |
| | | | Genetic | -0.08 | -0.11, -0.04 | 2.7e-04 |
| Birth PI | Maternal BMI | 44906 | Phenotypic | 0.09 | 0.08, 0.09 | 1.2e-104 |
| | | | Genetic | 0.00 | -0.05, 0.05 | 0.89 |
| | Paternal BMI | 50029 | Phenotypic | 0.04 | 0.03, 0.05 | 6.3e-28 |
| | | | Genetic | -0.05 | -0.09, -0.01 | 0.02 |

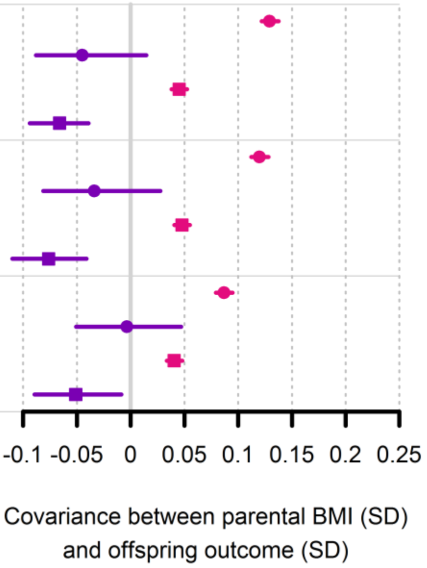

**Supplementary information S20: MCoTS results for the association of parental BMI with offspring eating behaviour (CEBQ) traits**

| Offspring outcome | Exposure | *n* | Covariance component | Estimate | 95% CI | *P* |
|---|---|---|---|---|---|---|
| Food responsiveness | Maternal BMI | 20074 | Phenotypic | 0.10 | 0.08, 0.11 | 9.7e-56 |
| | | | Genetic | 0.07 | -0.15, 0.17 | 0.28 |
| | Paternal BMI | 21573 | Phenotypic | 0.08 | 0.07, 0.09 | 7.6e-45 |
| | | | Genetic | 0.00 | -0.09, 0.10 | 0.94 |
| Fussiness | Maternal BMI | 20089 | Phenotypic | -0.01 | -0.02, 0.01 | 0.28 |
| | | | Genetic | 0.19 | -0.16, 0.24 | 1.1e-04 |
| | Paternal BMI | 21587 | Phenotypic | 0.00 | -0.01, 0.01 | 0.63 |
| | | | Genetic | -0.13 | -0.22, 0.03 | 0.02 |
| Satiety responsiveness | Maternal BMI | 20077 | Phenotypic | 0.02 | 0.01, 0.03 | 4.1e-03 |
| | | | Genetic | 0.08 | -0.15, 0.19 | 0.37 |
| | Paternal BMI | 21576 | Phenotypic | 0.00 | -0.01, 0.01 | 0.67 |
| | | | Genetic | 0.00 | -0.14, 0.12 | 0.97 |
| Slow eating | Maternal BMI | 20071 | Phenotypic | 0.01 | 0.00, 0.03 | 0.02 |
| | | | Genetic | -0.12 | -0.19, 0.05 | 0.03 |
| | Paternal BMI | 21570 | Phenotypic | -0.02 | -0.03, -0.01 | 1.2e-03 |
| | | | Genetic | -0.05 | -0.16, 0.07 | 0.28 |
| Emotional undereating | Maternal BMI | 19713 | Phenotypic | -0.02 | -0.03, -0.01 | 1.8e-03 |
| | | | Genetic | 0.09 | -0.01, 0.15 | 0.04 |
| | Paternal BMI | 21175 | Phenotypic | -0.02 | -0.04, -0.01 | 5.1e-05 |
| | | | Genetic | 0.09 | 0.02, 0.14 | 0.02 |
| Emotional overeating | Maternal BMI | 19807 | Phenotypic | 0.04 | 0.03, 0.05 | 4.3e-12 |
| | | | Genetic | 0.12 | -0.11, 0.18 | 0.06 |
| | Paternal BMI | 21269 | Phenotypic | 0.03 | 0.01, 0.04 | 9.6e-06 |
| | | | Genetic | 0.02 | -0.07, 0.12 | 0.61 |

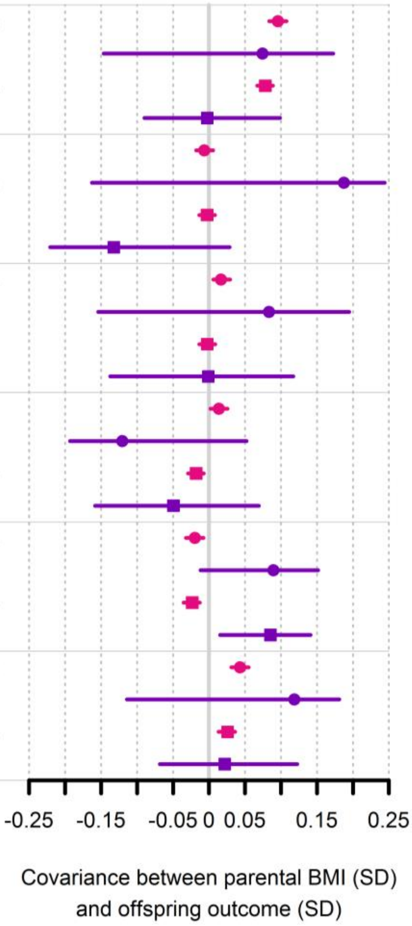


**Supplementary table S1**:

Full linear regression results for associations between exposures and outcomes (see separate excel file sup_tables.xlsx).

**Supplementary table S2:**

Full MCoTS results including model fit statistics and estimated variance components for parental and offspring phenotypes (see separate excel file sup_tables.xlsx).
